# Adiposity and sex steroid hormones: evidence on direct measures and changes over time -- a systematic review and meta-analysis

**DOI:** 10.64898/2026.09.21.26363542

**Authors:** Mihiretu M Kebede, Hannah Spahits, Srimanti Dutta, Renée Turzanski Fortner, Charlotte Le Cornet

## Abstract

**Introduction:** The evidence synthesis on literature characterizing changes in body fatness and adiposity distribution with sex steroid measurements in different matrices (blood, tissue, urine and other excreted matrices) is lacking.

**Methods:** PubMed and Web of Science were used to identify eligible studies published until June 2026. Random-effects meta-analyses were conducted, and the strength of evidence was determined based on the Continuous Update Project criteria.

**Results:** Of 16,875 screened references, 140 studies deemed relevant: 96 on circulating blood biomarkers from 39 weight loss intervention (surgical and lifestyle) and 60 observational studies reporting direct measure of adiposity distribution (e.g., DEXA, MRI), 20 studies on biomarkers measured in urine or other excreted matrices (i.e., saliva, urine, breast ductal lavage), and 25 tissue-based studies (e.g., adipose, breast, endometrial, skin).

Weight loss was associated with a reduction of total and free testosterone, and an increase in sex hormone binding globulin (SHBG). While no significant associations were found between weight loss and premenopausal estradiol, dehydroepiandrosterone sulfate, and androstenedione. Decrease in total and free estradiol and estrone were observed in postmenopausal women. Findings in observational studies generally supported those results, as well as tissue-based studies on estrogen-related markers despite the heterogeneous tissue types. Studies on biomarker measured in excreted matrices were largely inconclusive.

**Conclusion:** Strong evidence of a probable association was found between body fatness and SHBG, androgens, and postmenopausal estrogens. Limited evidence remained for other sex steroids.

## Introduction

The proportion of overweight and obese adults worldwide has more than doubled since 1990, reaching 43% and 16%, respectively, in 2022 ^1^, which means 1 in 8 people in the world is living with obesity. Current trend projections suggest that, by 2030, half of the adult population will have a high body mass index (BMI>25kg/m2)^2^. The World Cancer Research Fund (WCRF) Third Expert Report identified twelve different cancer types for which overweight and obesity are at least “probable” risk-increasing factors ^3^. In 2022, 3.4% of cancers in the world were attributable to high BMI ^4^. This proportion was higher in women compared to men (5.4% vs. 1.9%), with 31% of endometrial cancers, 6.9% of breast cancers, and 3.9% of ovarian cancers attributable to excess body weight ^5^. Sex steroids are an established mechanistic pathway underlying the association between excess body fatness and increased risk of breast, ovarian and endometrial cancer risk, notably for predominantly hormone-dependent sub-types (ER+ post-menopausal breast cancer, type I endometrial cancer, and endometrioid ovarian cancer). Adipose tissue is the predominant source of estrogens in obese women ^6^ which makes adiposity to be strongly associated with higher estrogen and androgen concentrations in both pre- ^7^ and post-menopausal ^8–11^ women. Although pooled analyses have reported associations between circulating sex steroids and indirect measures of body fatness such as BMI ^7,8^, no large systematic review has comprehensively summarized the evidence on the association between changes in direct measures of body fatness and adipose depot (i.e., distribution of adipose tissue) and sex steroid measurements in different matrices (blood, tissue, urine and other excreted matrices) so far.

This systematic review and meta-analysis aims to synthesize and appraise the evidence for an effect of body fatness on sex steroid hormones levels in women in 3 dimensions, evaluating the associations between i) Changes in body fatness (e.g., as induced by bariatric surgery or lifestyle intervention) and circulating sex steroid hormones ii) Adiposity distribution (e.g., direct measures of body fatness such as body fat percentage i.e., measured by DXA, and measures of fat distribution compartments e.g., visceral (VAAT) or subcutaneous (SAAT) abdominal adipose tissue) and levels of circulating sex steroid hormones iii) Adiposity measures (e.g., BMI, body fat distribution compartments) and sex steroid hormones levels measured in biospecimens outside of blood (e.g., breast tissue, urine).

## Materials & methods

The review was registered in PROSPERO database (CRD42020201611) and conducted in accordance with the Preferred Reporting Items for Systematic Reviews and Meta-Analyses (PRISMA) statement ^12^, and with the framework to conduct systematic reviews on mechanistic pathways developed by the WCRF/University of Bristol’s ^13^.

### Literature search

Comprehensive search algorithms were developed in PubMed and Web of Science (WoS) to identify studies reporting the relevant exposure (changes in body fatness, adiposity distribution, adiposity measure), and outcomes of interest (sex steroid hormones). The full description of the search strategy can be accessed in Table S1 and Table S2. Overall, 24,737 references (9,955 in PubMed and 14,792 in WoS) were identified relevant for our review as previously described^14^. The searches were completed on 19 March 2020 in Pubmed, and on 23 and 25 March 2020 in WoS. After excluding 7,862 duplicates between the two databases, 16,875 titles/abstracts were screened independently by two reviewers in Covidence. An updated search was conducted in June 2026.

### Title/abstract screening

#### Study inclusion

Original articles published in English, including women 18 years old or older, which examined the effect of body fatness on sex steroid hormones were included eligible following the following criteria; Intervention studies including lifestyle (dietary, physical activity, behavioural) or surgical (biliopancreatic diversion, gastric bypass, gastric banding, sleeve gastrectomy, vertical banded gastrectomy) interventions which reported significant body fatness change (any of direct or indirect body fatness type) or change of more than 10% ^15^ over a period of at least 3 months follow-up. Longitudinal follow-up studies with changes of sex steroids and body fatness between 2 time points were also included and included in meta-analysis of lifestyle intervention studies. Other observational studies were also included. Given already existing comprehensive meta-analysis on associations between BMI and sex steroids ^8^, we focused on observational studies having direct measures of body fatness only (measured by dual-energy X-ray absorptiometry, magnetic resonance imaging, or adipocyte size).

Additionally, all studies with association reported between body fatness (any of direct or indirect body fatness type) and sex steroids measured in tissue localized in the breast, ovary, endometrium, or around gynecological organs (VAAT, SAAT), or in urine and other excreted matrices (saliva, ductal, urine) were included when women were not using exogenous hormones or it remains unknown.

#### Study exclusion

Articles with no sex-stratified associations reported, or study samples restricted to men were excluded. Intervention studies without values reported on hormone concentration before/after the intervention were excluded. We also excluded observational studies reporting indirect measures of body fatness (e.g., BMI, WHR) only, or observational studies without assessment of the association between body fatness and sex steroid concentrations. For circulating estrogens and progesterone, we excluded studies that did not report values stratified by menopausal status, menstrual cycle phase in premenopausal women, and studies where women used exogenous hormones (menopausal hormone therapy or oral contraceptives) at biospecimen (blood, tissue, urine and other excreted matrices) collection. No exclusion based on menopausal status and menstrual cycle phase was applied for studies measuring estrogens in tissue, urine or other excreted matrices.

Additionally, studies on women with conditions or treatments inclined to alter body fatness and/or sex steroid levels (cancer, polycystic ovary syndrome, anorexia, infertility) were excluded, as well as experimental studies (animal and cell line). If multiple studies were published on the same population and association, the most comprehensive one was included and the other(s) excluded.

### Full text review and data extraction

Full texts were reviewed and relevant studies were extracted and critically appraised by two reviewers in a database created in Epiinfo. The main characteristics of each study were extracted including study and population characteristics, measures of adiposity, measures of sex steroids, measure of association/between group differences, and adjustment variables.

#### Evidence appraisal

The overall strength and level of evidence was adapted from the tool used in the continuous update report program ^16^ with five categories: strong evidence (convincingly causal, probably causal, substantial effect on risk unlikely), or limited evidence (suggestive of a possible causal relationship, no conclusion of a causal relationship possible). An upgrade in level was given when the meta-analysis remains significant even after removing the outliers. An upgrade in level was given if the meta-analysis remains statistically significant after excluding the outliers.

### Statistical analysis

Units of circulating sex steroids concentration at baseline were harmonized (Table S3). Medians were converted to means, and similarly, confidence intervals (CIs), interquartile ranges (IQRs), or standard errors (SEs) were converted to standard deviations (SDs)^17^. Random effects meta-analyses were conducted for i) intervention studies and ii) the correlation between circulating biomarkers and direct measures of body fatness. Due to the substantial heterogeneity of reported statistical methods (e.g., regression, ANOVA) and adjustment factors, meta-analyses other than those based on correlations were not feasible for observational studies. Similarly, due to the heterogeneity of biospecimens and the paucity of studies, no meta-analysis could be conducted on studies with biomarkers measured in biospecimens outside of blood. Studies not included in the meta-analysis were summarized narratively.

Meta-analysis was performed where exposures, outcomes, and statistical analysis were consistent in at least three separate study samples. All meta-analyses for estrogens were stratified by menopausal status and menstrual cycle phase.

All analyses were conducted in R. Standardized mean difference (SMD) was computed for intervention studies using *meta* ^18^ and *dmetar* ^19^ packages. SMD was calculated when the intervention study reported mean value of circulating sex steroid concentrations at baseline, and at follow-up, and at least one standard deviation (i.e., either at baseline or follow-up time) was reported. When a study reported more than one follow-up, we used the data from the last reported follow-up. In case of different arms reported in one study, individual arms were included as independent samples (e.g., Kraemer et al., 2003 has 2 sub-groups of women randomized to periodized and non-periodized resistance training ^20^). Meta-analysis of correlation was conducted for observational studies using *metacor* package. Meta package performs the Fisher’s z-transformation before pooling, and pooled correlation coefficients up to 0.19 were considered very weak, 0.2-0.39 as weak, 0.40-0.59 moderate, 0.6-0.79 strong and 0.8-1 very strong ^21^. Meta-analyses were conducted separately for unadjusted and adjusted correlation and only adjusted correlation was reported in the descriptive results when available. Moreover, meta-analysis of standardized regression coefficients was possible only for two biomarkers. No other meta-analysis was possible for observational studies due to the substantial heterogeneity of statistical methods, adjustment factors, and inconsistency in measurements of adiposity. The between-study heterogeneity was estimated using tau-squared (τ²). The I² statistic was used to quantify the proportion of variation across studies in the meta-analysis that is attributable to heterogeneity rather than chance interpreted as not important heterogeneity when I^2^<40%, moderate up to 60% and substantial beyond ^22^. Outlier analyses were performed using *find.outliers* function of the *meta* package ^19^, to identify and exclude studies with extreme effect sizes. A sensitivity analysis stratified by menopausal status was conducted for all biomarkers when 3 or more studies were available for both pre- and post-menopause. Publication bias was assessed through visual inspection of funnel plot symmetry when at least 10 comparison groups were included in the meta-analysis. The overall conclusion of evidence is provided for each sex steroids individually and summarised in Fig. S1.

Studies with biospecimens outside of blood were summarized visually with parallel categories (ParCat) plots using *plotly.express library’s parallel_categories* function in Python.

## Results

### Studies characteristics

Of 16,875 title/abstract screened, 963 studies were selected for full text review. After the exclusion of intervention studies without significant change in body fatness, observational studies on indirect measure of body fatness such as BMI (main reason n=365), as well as studies with women not eligible due to health precondition or medications (e.g. pregnant, using exogenous hormones), or studies having no relevant association, 140 studies were deemed relevant for inclusion (Figure 1 and supplementary results). Study characteristics are presented in Table S4). A total of 96 studies were found for circulating sex steroids; 38 intervention studies (surgical and lifestyle) ^20,23–59^, one longitudinal study reporting on 2 time points ^60^, and 60 observational studies on direct measure of body fatness ^11,37,41,61–118^ among with studies reporting association based on ANOVA ^62,72^; p-values ^63^; p-trends ^113^; or regression ^11,79,86,88,98–100,103,105,112,115–117^ had a high degree of heterogeneity and therefore not meta-analysed.

**Figure 1:**
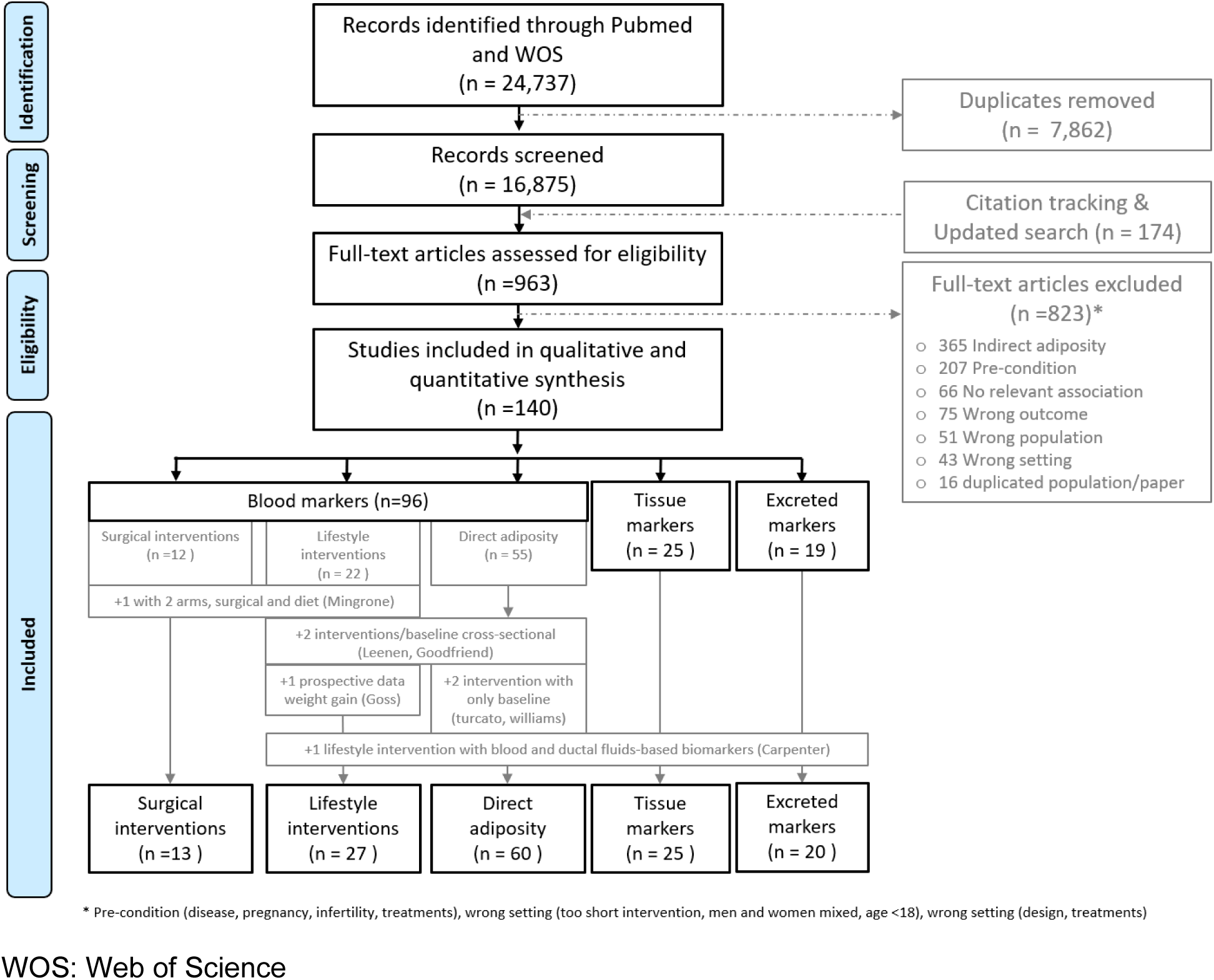
PRISMA diagram on the systematic literature review on association between body fatness and sex steroid hormones.

Additionally, 20 studies on body fluid-based markers ^56,119–136^, and 25 studies on tissue-based markers ^137–161^ were included. Seven studies had more than one study design or biospecimen and were therefore included in different groups of study as shown in Figure 1 ^34,37,41,56,60,110,111^. A total of 64 studies were exclusively on premenopausal women ^20,23–28,34,37–43,59,63–66,68,71,73,74,76,78,82,86,87,89–91,94,95,97,102–104,108,109,111,113,114,118–130,137–142,162–164^, 45 studies on postmenopausal women ^11,35,36,44–56,60–62,67,69,70,72,81,83,85,88,93,98,100,101,106,131–136,143–147,165,166^, eight studies reported findings stratified by menopausal status ^29,57,96,105,110,148–150^, 16 studies included pre and postmenopausal women mixed ^32,75,80,99,112,115,151–157,159–161^, and seven studies did not report menopausal status ^30,31,58,84,116,117,167^.

The mean baseline BMI of subjects receiving surgical interventions was between 39.2 (Menezes, 2021) and 56.7 (27) kg/m^2^ and weight between 103.8 (Menezes, 2021) and 148.0 (27) kg, while lifestyle interventions had mean BMI values ranging from 25.0 (48) to 36.1 (39) kg/m2 and weight from 78.0 (50) to 121.6 (36) kg.

In observational studies reporting direct measure of body fatness, the mean body fat percentage ranged from 20.1 ^91^ to 48.1 ^67^ %, while mean fat mass ranged from 14.4 ^65^ to 52 ^64^ kg. The mean of abdominal adipose tissue ranged from 184.5 ^76^ to 585.5 ^110^ cm^2^ when SAAT, from 40.2 ^71^ to 184.3 ^110^ cm^2^ when VAAT, from 81 ^66,80^ to 195.9 ^84^ cm^2^ for intra-abdominal adipose tissue (IAAT), from 260.3 ^71^ to 653.4 ^109^ cm^2^ for total abdominal adipose tissue (TAAT). Total breast fat volume ranged from 277.5 ^68^ to 502.8 ^87^ cm^3^.

#### EVIDENCE FROM TISSUE-BASED MARKERS

Due to the large heterogeneity of tissue types, adiposity measures, and analysis, the tissue-based studies could not be meta-analysed, but Sankey plots show a qualitative visualization of the associations (supplementary figures S2, S3, S4).

### Aromatase

A total of eight studies reporting aromatase expression were identified ^147,148,152,153,157^ ^140,147,150^.

Three studies were from the same scientific group but on different study samples of women undergoing mastectomy for breast cancer risk reduction ^148,152,153^. One of the studies had stratified analyses by BRCA1 and BRCA2 status and menopausal status ^148^, while the two others did not ^152,153^. Five studies reported aromatase measured in breast tissue ^148,150,152,153,157^, which reported positive associations between adiposity and aromatase expression. Further studies evaluated aromatase expression in endometrial tissue ^140^, in VAAT or in SAAT ^147^.

In premenopausal women, a weak non-significant correlation was reported between BMI and aromatase mRNA expression measured in endometrial tissue (r=0.28, p=0.06) ^140^. Two studies investigated aromatase mRNA expression measured in breast tissue from premenopausal women and found divergent results: a study on BRCA1 and BRCA2 mutation carriers found a significant positive trend of aromatase expression with increasing BMI categories (p<0.001) ^148^, whereas a study on non-carriers observed no significant association ^150^. Similar results in breast tissue were reported for postmenopausal women ^148,150^. A weak significant positive correlation was found between BMI and aromatase mRNA expression measured in VAAT (r=0.23, p<0.05), but not significant when measured in SAAT ^147^.

Four additional studies found positive associations between BMI and breast aromatase mRNA levels in pre- and postmenopausal women ^150,152,153,157^. Moreover, two studies reported the association stratified by menopausal status, but no difference was observed per menopausal status ^148,150^.

### ERα (ER, ESR1 and ERα)

A total of 12 studies investigated adiposity with ERα receptor expression levels measured in breast ^150,156^, endometrial ^142,144,151^, adipose tissue (AT) ^137,138,143,147,154,155^, and skin tissue ^149^.

In premenopausal women, five studies observed no significant associations between BMI and ERα mRNA expression in the breast ^150^ or SAAT ^138^, as well as with Erα expression levels in endometrium ^142^, gluteal subcutaneous AT ^137^, and skin ^149^.

In postmenopausal women, one lifestyle intervention showed that weight loss and reduction in body fat percentage was associated with higher ERα mRNA expression in SAAT, although this association was not significant after adjusting for multiple comparisons ^143^. No significant association was also reported between BMI and ERα mRNA expression in breast ^150^. One study reported that lower BMI was associated with higher ERα protein levels in endometrial glandular tissue ^144^, while this association was positive when ERα levels were measured in skin ^149^.

Out of the four studies including both pre- and post-menopausal women, one reported an inverse association between BMI and Erα DNA methylation in breast tissue ^156^, and three observed no significant associations between total body fat percentage and Erα mRNA expression in femoral AT or in SAAT ^155^, as well as between BMI and Erα levels in SAAT or in VAAT ^154^. Furthermore, a bariatric surgery intervention did not find significant differences in ERα expression levels measured in endometrial tissues three months post-intervention ^151^.

### ERβ

A total of four studies investigated the association between adiposity and ERβ. Four studies measured ERβ in AT ^137,154,155^, and one in skin ^149^.

In premenopausal women, ERβ protein measured during the follicular phase was weakly inversely correlated with WHR (r=−0.315, p=0.03) (ERβ in gluteal subcutaneous AT ^137^) but not with BMI (ERβ in skin tissue ^149^). In postmenopausal women, only one study was found and reported a positive association between BMI and ERβ levels in skin tissue (p<0.02) ^149^. The two studies included both pre- and post-menopausal women and reported a significant association neither between BMI and ERβ levels in SAAT or in VAAT ^154^ nor between body fat percentage and ERβ mRNA expression in femoral AT or in SAAT ^155^.

### ERα/ERβ ratio

Three studies reported the association between the ERα/ERβ ratio measured in AT and anthropometric measures ^137,154,155^. One study included premenopausal women in the follicular phase of the menstrual cycle and reported a positive association between WHR and the ERα/Erβ protein ratio measured in gluteal subcutaneous AT (r= 0.406, p=0.01) ^137^. No studies were found on postmenopausal women. Two studies in both pre- and post-menopausal women reported a weak negative correlation between body fat percentage and ERα/ERβ mRNA expression ratio measured in SAAT (r=−0.359, p<0.05), and not significant when measured in femoral AT ^155^, while the other reported no significant correlation between BMI and ERα/Erβ protein ratio when measured in either SAAT or VAAT ^154^.

### Estradiol

Three studies were identified on estradiol measured in breast ^146,150^ and in adipose ^145^ tissues. In premenopausal women, one study without information on menstrual cycle, reported no significant association between BMI and estradiol measured in breast tissue ^150^.

In postmenopausal women, overweight women were shown to have higher estradiol levels in breast tissue compared to normal-weight women (P< 0.05) ^150^, and a moderate positive correlation was observed between BMI and breast tissue estradiol (r=0.52, p=0.03) ^146^. However, one study found no significant correlation between BMI and estradiol measured in SAAT or in VAAT (p≥0.05) ^145^.

### Estrone

Two studies were identified in postmenopausal women and showed that estrone measured in breast tissue (r=0.48, p=0.05) ^146^ and VAAT (r=0.46, p<0.05) ^145^ was moderately correlated to BMI. However, no significant correlation was found between BMI and estrone measured in SAAT (r= 0.32, p≥0.05) ^145^.

### Progesterone receptors (PR)

Five studies reported PR expression on endometrial and skin tissues ^141,142,144,149,151^. In premenopausal women, BMI was inversely associated with PR protein in endometrial ^141^ and skin tissues^149^ among women who were in the follicular phase of the menstrual cycle. However, no significant correlation was observed with PR expression levels in the endometrium (e.g., PR stromal epithelium, r=0.20, p>0.05) ^142^ of women during this phase. In postmenopausal women, PR protein expression in skin tissue was reported to be higher in obese than in normal BMI women (p<0.006) ^149^, while no significant association was found between BMI and PR protein in endometrial tissue ^144^. Additionally, in a study that included both pre- and post-menopausal women without stratified analysis, a weight loss intervention measured PR in endometrial tissue before and after 12 months of follow-up and found a significant decrease in PR protein H-score^151^.

### 17βHSD

The enzyme in sex steroid metabolism, 17βHSD, was evaluated in two studies ^139,143^. In premenopausal women, one study reported a moderate correlation between BMI and 17βHSD5 mRNA expression measured in SAAT adipocyte (r=0.50, p=0.03), while no correlation was shown when measured in SAAT or VAAT preadipocyte, or VAAT adipocyte (r<0.10, p>0.05)^139^. In postmenopausal women, no significant changes in 17βHSD measured in SAAT were observed after 6 months of weight loss lifestyle intervention, after adjustment for multiple testing ^143^.In contrast, 17βHSD1:2 measured in VAAT was strongly negatively correlated with BMI (r=−0.75) in another study ^147^.

#### EVIDENCE FROM CIRCULATING MARKERS

##### Estradiol (E2)

A total of 12 intervention studies ^28,33,35,36,41,42,51,53–56,60^ were included, as well as 19 observational studies on direct measures of body fatness ^11,41,68,72–74,81,87,88,91,93,94,96,97,103,104,109,110,164^, six studies investigated urine-based estradiol 120,125,127,129,134,136, one study analysed estradiol in saliva specimens ^130^, and one on ductal fluid estradiol ^56^.

##### Premenopausal women

***Intervention***: One surgical intervention study showed significant decrease in circulating luteal estradiol level after significant weight loss ^28^, while the meta-analysis of three lifestyle intervention studies with significant adiposity loss showed a non-significant change in circulating follicular estradiol (SMD= −0.24 [−0.66; 0.17], I^2^=82%, Fig. S5) ^33,41,42^.

***Observational:*** *A* meta-analysis on three studies for the association of circulating levels of follicular estradiol with VAAT showed a very weak significant inverse correlation (pooled r= −0.18 [−0.28; −0.07], I^2^=59%, Fig. S6) ^104,109,110^. However, the significant pooled correlation was no longer observed after multivariable adjustment ^41,109,110^. No meta-analysis was possible for the remaining studies ^68,74,87,97,103,164^.

***Urine and other excreted matrices:*** During the follicular phase, no significant changes in urinary estradiol or its metabolites (2-OHE2, 4-OHE2, 2-MeOE2, and 4-MeOE2) were observed following a physical activity intervention with significant weight loss ^120^. In observational studies, no significant associations were observed between urine-based estradiol in the follicular ^125^ or luteal phase ^125,127,129^ and BMI. One study measuring estradiol levels in saliva found a non-linear association with body fat percentage ^130^.

***Conclusion:*** Strong evidence of substantial causal association unlikely given consistent null results of the meta-analysis, supported by a study on breast tissue.

##### Postmenopausal women

***Intervention:*** No surgical intervention studies were identified. A meta-analysis of lifestyle intervention studies showed a significant decrease in circulating estradiol with loss of adiposity (SMD= −0.88 [−1.47;-0.29], I^2^=98%, seven studies and eleven comparison groups, Fig. S7) ^35,36,51,53–56^. The exclusion of the outlier ^53^ does not change the estimates meaningfully (SMD= −0.58 [−0.90;-0.26], I^2^=84%), Additionally, two other intervention studies and one longitudinal study recorded an increase in circulating estradiol after weight gain ^35,53^ and significant IAAT increase^60^.

***Observational:*** Meta-analysis was not feasible ^11,72,81,88,93,96^.

***Urine and other excreted matrices:*** One dietary intervention study did not find a significant correlation between weight loss and changes in urine-based estradiol (r=0.15, p=0.17) ^134^. A cross-sectional study found a weak correlation between BMI and estradiol and estradiol glucuronide levels in urine (r=0.20, p<0.05 and r=0.25, p<0.01, respectively) ^136^. After a 12-week lifestyle intervention in obese women, 24% reduction of estradiol measured in breast ductal fluid was observed following a fat mass loss of 14% ^56^.

***Conclusion:*** Strong evidence of probable causal association given consistent evidence from lifestyle intervention studies which show significant association in meta-analysis, although heterogeneity remained unexplained (lifestyle intervention, I^2^=98%). The strong evidence of probable association is consistent with the findings from urine and tissue-based studies.

##### Free estradiol

A total of seven intervention studies ^33,35,41,51,53–55^ were found, nine observational studies ^11,41,62,68,79,87,88,103,164^, and one study with free estradiol index measured in urine ^131^.

##### Premenopausal women

***Intervention:*** There was no surgical intervention studies identified. Two lifestyle intervention studies reported contradictory findings ^33,41^. One reported a significant decrease in estradiol with BMI, weight, fat mass, VAAT, WHR and abdominal fat loss ^41^, while the other showed an increase in estradiol concentration following the weight loss intervention ^33^.

***Observational:*** Due to large methodological heterogeneity, meta-analysis was not feasible to quantitatively synthesize the results of the observational studies ^41,68,87,103,164^.

***Conclusion:*** Limited evidence – no conclusion

##### Postmenopausal women

***Intervention:*** No surgical intervention studies were identified. The meta-analysis of lifestyle intervention studies showed a non-significant change in free estradiol after significant loss of body fatness (SMD= −0.84 [−1.64;-0.04], I^2^=98%, six studies and nine comparison groups, Fig. S8) ^35,50,51,53–55^. After the exclusion of the outlier ^53^, the between-study heterogeneity decreased (SMD=−0.41 [−0.61;-0.21], I^2^=84%). Additionally, two other intervention studies recorded an increase in circulating free estradiol with weight gain ^35,53^, although significant in the study with the highest weight changes only ^35^.

***Observational:*** No meta-analysis was possible for the remaining studies ^11,62,79,88^.

***Urine and other excreted matrices:*** The free estradiol index measured in urine of postmenopausal women was significantly correlated with BMI (r=0.50, p<0.01) and truncal fat (r=0.35, p<0.01) but not with body fat percentage (r=0.24, p=0.07) ^131^.

***Conclusion:*** Strong evidence of probable causal association because meta-analysis on lifestyle intervention showed significance, but heterogeneity remained unexplained (I^2^=79%). This probable causal association is also in line with studies on urine-based markers.

##### Estrone (E1)

A total of nine intervention studies ^35,41,42,45,51,53–55,60^ were included, nine observational studies ^41,74,81,83,88,94,96,100,104^, six studies on urine-based estrone ^120,125,127,129,134^, Mole, 1989 #597 or urine-based estrone production rate ^124^.

Regarding estrone conjugate, two intervention studies ^33,56^, two observational studies ^11,88^ investigated estrone sulfate. Urine levels of estrone glucuronide were also assessed in four observational studies ^119,123,126,136^, and overall conjugated estrone was assessed in one study ^128^. Moreover, 5 observational studies on estrone metabolites investigated hydroxyestrone (OHE) in urine d ^120–122,129,131–133^.

##### Premenopausal women

***Intervention:*** No surgical intervention studies were identified but two lifestyle intervention studies reported no significant change in estrone concentrations after significant reductions in adiposity ^41,42^.Similar results were reported for estrone sulfate ^33^.

***Observational:*** No meta-analysis was possible for the remaining studies ^41,74,94,104^.

**Urine and other excreted matrices:** A physical activity intervention did not show significant association between weight loss and follicular estrone measured in urine ^120^. Similarly, no significant correlations were reported between BMI and follicular ^125^ or luteal urinary estrone levels ^125,127,129^. One study reported a higher estrone production rate in women with obesity located in the lower part of the body, relative to those with upper-body obesity ^124^.

One intervention study found a lower concentration of estrone glucuronide in urine (all phases of menstrual cycle) following bariatric surgery leading to >25% weight loss ^119^. In contrast, three observational studies found no significant difference in estrone glucuronide concentrations ^123,126^ or overall conjugated estrone levels ^128^ across BMI groups, whether measured in the follicular phase ^123^ or across different phases of menstrual cycle ^126,128^.

Out of the four studies that investigated the associations between adiposity and levels of urinary estrone metabolites ^120–122,129^, two lifestyle intervention studies did not observe significant change in 2-OHE1 following a 4-months intervention leading to significant loss of body fat mass ^120,121^. However, conflicting results were observed for 16a-OHE1 and 2:16a-OHE1, particularly in the follicular phase. Furthermore, no significant changes were reported for 4-OHE1, 2:4-OHE1, 2-MeOE1, and 4-MeOE1 ^120^. Observational studies tended toward a very weak inverse association between adiposity and urine concentration of 2-OHE1 ^122,129^. Two intervention studies ^120,121^ as well as two observational studies ^122,129^ reported contradictory associations for urinary 16α-OHE1 levels. Similarly, examining the 2:16*a*-OHE1 ratio, inconsistent findings were found in intervention studies ^120,121^ and one observational study ^122^.

***Conclusion:*** Limited – no conclusion

##### Postmenopausal women

***Intervention:*** No surgical intervention studies were identified. The meta-analysis of lifestyle intervention studies did not suggest significant change in circulating estrone levels after significant loss of adiposity (SMD=−0.30 [−0.73;0.13], I^2^=97%, six studies and nine comparison groups, Fig. S9) ^35,45,51,53–55^. After the exclusion of one outlier ^53^, the SMD was statistically significant and the between-study heterogeneity was drastically reduced (SMD=−0.15 [−0.22;-0.08], I^2^=20%). One longitudinal study observed a positive association between increase in IAAT and circulating estrone over two years follow-up ^60^.Two intervention studies with weight gain showed conflicting results ^35,53^. One lifestyle intervention study also reported no significant change in circulating estrone sulfate following significant adiposity loss ^56^.

***Observational:*** Meta-analysis of unadjusted correlations suggested a weak significant positive correlation between estradiol and fat mass (r=0.22 [0.12;0.31], I^2^=20%, two studies and four comparison groups, Fig. S10) ^81,83^, but no adjusted correlations was available. Linear regression beta coefficients reported for the association of circulating estrone levels and body fat percentage was reported in two studies ^88,100^, showing contradictory results. However, two studies on concentrations of estrone sulfate showed a significant positive association with body fat composition ^11,88^.

***Urine and other excreted matrices:*** One dietary intervention study concluded on a non-significant correlation between BMI and urinary estrone at baseline, but found a modest positive correlation with weight loss (r=0.33, p=0.017) ^134^. A cross-sectional study found weak correlations between BMI and estrone and estrone glucuronide (r=0.25, p<0.01 and r=0.20, p<0.05, respectively) ^136^. Regarding hydroxyestrones, two studies reported very weak to weak inverse correlations between urinary 2-OHE1 and abdominal fat (r =−0.17, p=0.03) ^132^ at baseline of an intervention study, and BMI (r=−0.36, p<0.01) in a cross-sectional study ^131^. A weak inverse correlation was also observed between abdominal fat and 16α-OHE1 (r =−0.20, p=0.01) ^132^, but the correlation was weaker and not significant with BMI (r=−0.12, p=0.25) ^131^. The 2:16*a*OHE1 ratio was not correlated with abdominal fat (r =−0.02, p=0.75) in one study ^132^, while a weak inverse correlation was reported with BMI in two studies (r=−0.29, p<0.01)^131^, and (r=−0.22, p=0.20) ^133^, respectively.

***Conclusion:*** Strong evidence of a probable causal association between adiposity and postmenopausal estrone levels as the meta-analysis on intervention studies is significant after outlier removal and the heterogeneity is low (lifestyle intervention, I^2^=0%; observational I^2^=20%). These results are consistent with findings from studies on tissue and urinary measurements. However, the evidence on estrone conjugates and their metabolites in postmenopausal women is insufficient to draw reliable conclusions

##### Testosterone

A total of 23 intervention studies ^23,24,26–31,33,36,38–43,47,51–55,60^, and 29 observational studies on direct measures of body fatness ^11,41,63,64,68–71,73,74,82,84,85,87–89,92,94–96,98,99,102,104,108–110,112,113^ were identified. No body-fluids-based studies were identified.

***Intervention:*** The meta-analysis on surgical weight loss interventions showed a significant reduction in circulating testosterone levels (SMD=−0.47 [−0.65;-0.29], I^2^=65%, ten studies and eleven comparison groups, Fig. S11) ^23,24,26–31,58,59^. The association became slightly stronger after the exclusion of one study ^24^ reporting an outlier value (SMD=−0.52 [−0.67;-0.37], I^2^=52%).

The meta-analysis on lifestyle weight loss intervention studies showed a statistically significant reduction in circulating testosterone levels (SMD=−0.27 [−0.43;-0.11], I2=90%, 14 studies and 20 comparison groups, Fig. S12) ^33,36,38–43,47,51–55^. Although the effect size was smaller, the reduction remained significant after excluding outlying values ^36,38,53^ (SMD=−0.12 [−0.19;-0.05], I2=45%). However, the funnel plot showed asymmetry (Fig. S13) suggesting small-study effects. No significant change in circulating testosterone level was reported in two intervention studies reporting weight gain ^43,53^ as well as in a longitudinal study reporting a significant increase in IAAT in 2-years follow-up ^60^. Stratified analysis did not show substantial change in the interpretation (supplementary results).

***Observational:*** The meta-analysis indicated a weak significant positive correlation between circulating testosterone levels and fat mass (pooled r=0.30 [0.11; 0.47], I^2^=0%) ^71,96^, as well as with SAAT (r=0.25 [0.05; 0.44], I^2^=0) ^71,96^, each with two studies and three comparison groups (Fig. S14). In contrast, the pooled results for correlation of circulating testosterone and VAAT were not significant (r=0.05 [−0.21; 0.30], I^2^=74%, six studies and 8 comparison groups) ^41,64,71,82,96,110^. The meta-analysis of standardized beta coefficients from adjusted linear regression models for the association between testosterone and breast fat volume indicate a non-significant association, and high between-study heterogeneity (pooled beta=4.42 [−5.63;14.48], I^2^=100%, Fig. S15) ^68,74,87^. No meta-analysis was feasible for the remaining studies ^11,88,98,99,109,112^ and study on weight gain showed significant positive trend in testosterone mean across categories of body fat ^113^.

***Conclusion:*** Strong evidence of probable causal association as consistent evidence was observed across meta-analysis, although some unexplained heterogeneity remained (surgery intervention, I^2^=51%; lifestyle intervention, I^2^=40%; observational, I^2^=0-100%).

##### Free testosterone (free, and bio testosterone)

A total of 14 intervention studies ^23,28,29,33,36,38,41,42,50,51,53–55,60^, and 19 observational studies on direct measures of body fatness ^41,64,67–71,73,79,82,85,87,94,96,98,104,108,110,112^ were identified. No studies on urine or other excreted matrices were identified.

***Intervention:*** The meta-analysis on surgical intervention indicated a significant decrease in free circulating testosterone following loss of adiposity (SMD=−0.77 [−1.03;-0.51], I^2^=37%, four studies and five comparison groups, Fig. S16) ^23,28,29,58^ and no outlier was detected. Similarly, our pooled analysis of lifestyle interventions showed a significant reduction in circulating free testosterone levels following significant loss of adiposity (SMD=−0.59 [−1.01;-0.18], I^2^=97%, ten studies and 14 comparison groups, Fig. S17) ^33,36,38,41,42,50,51,53–55^. After excluding studies with outlier values ^33,38,53^, the decrease in free testosterone remained statistically significant and the heterogeneity decreased (SMD=−0.33 [−0.49;-0.17], I^2^=69%) despite the effect size was reduced. The funnel plot showed asymmetry (Fig. S18), suggesting small-study effects. Weight and IAAT gain were associated with a significant increase in circulating free testosterone in an intervention study ^53^ but showed no significant change in a longitudinal study ^60^. Stratifying our meta-analysis by menopause status indicates a borderline significant association in postmenopausal women (SMD=−0.50 (−0.99;0.00), I2=97%) (supplementary results).

***Observational:*** Meta-analysis of adjusted coefficients indicated a weak significant positive correlation between circulating free testosterone and VAAT (pooled r=0.31 [0.14;0.46], I²=29%) ^41,71,82,96,110^. Similarly, pooled analysis for the correlation of circulating free testosterone and fat mass (pooled r=0.30 [0.11; 0.47], I²=0%,) ^82,96^, as well as SAAT (pooled r=0.33 [0.13; 0.5], I²=0%) ^71,96^ suggested significant positive correlation (Fig. S19). No meta-analysis was possible for the remaining studies. ^68,79,87,98,112^.

***Conclusion:*** Strong evidence of probable positive causal association as consistent results were observed in meta-analysis. The strength of evidence is stronger in postmenopausal women relative to premenopausal women.

##### Androstenedione

A total of 12 intervention studies ^24,26,28,31,39,41,52,54,55,60^, and 14 observational studies evaluating direct measures of body fatness ^11,41,63,70,73,74,78,82,85,88,89,94,99,110^ and circulating androstenedione, and androstenediol ^88^, androsterone glucuronide ^70^ and androsterone ^89^ were identified. No studies on urine or other excreted matrices were identified.

***Intervention***: Meta-analysis of surgical intervention showed a significant reduction in circulating androstenedione levels after adiposity loss (SMD=−0.32 [−0.52;-0.11], I^2^=59%, Fig. S20) ^24,26,28,31,58,59^ and no outlier was detected. Similarly, the meta-analyse of the lifestyle intervention studies suggested changes in androstenedione levels but this was not significant (SMD=−0.11 [−0.25;0.03], I2=68%, five studies and ten comparison groups, Fig. S21) ^39,41,52,54,55^. Excluding one study reporting an outlier ^41^ did not meaningfully alter the pooled estimate but no between-studies heterogeneity remained (SMD=−0.05 [−0.12;0.01], I2=0%). The funnel plot showed asymmetry (Fig. S22) suggesting small-study effects. Studies reporting gain in weight and IAAT showed significant decrease in circulating androstenedione in a longitudinal study ^60^ but not significantly changed in an intervention study ^53^. Stratifying our pooled analysis by menopause status yielded comparable results, with borderline effect observed in postmenopausal women (SMD=−0.07 [−0.14;-0.00], I2=0%) (supplementary results).

***Observational:*** The meta-analysis indicated no significant correlation between circulating levels of androstenedione and VAAT (pooled r=−0.16 [−0.51; 0.24], I^2^=73%, three studies and four comparison groups, Fig. S23) ^41,82,110^. None of the analyses on the association of androstenedione with direct adiposity measures using linear regression revealed any significant associations ^11,74,88,99^, except one study reporting a significantly positive association with total breast fat volume ^74^. Concentrations of circulating androstenediol ^88^, androsterone glucuronide ^70^, and androsterone ^89^ levels in relation to adiposity also showed no significant associations.

***Conclusion:*** There is no strong evidence for a substantial effect on androstenedione, unless a very high body fat loss, which may also be accompanied or triggered by other related biomarkers. The effect might be stronger in postmenopausal women. Limited evidence – no conclusion on androstenediol, androsterone glucuronide, androsterone due to the lack of studies.

##### SHBG

A total of 32 intervention studies ^20,23,26–36,38,39,41,43–55,57,60^, and 28 observational studies evaluating direct measures of body fatness ^11,41,65–70,73,74,79,80,82,85,87,88,94–96,99,101,102,105,106,110,112,114,163^ were included. No studies on urine or other excreted matrices were identified.

***Intervention:*** The meta-analysis on surgical intervention showed a statistically significant increase in SHBG following adiposity loss (SMD=2.62 [1.72;3.52], I^2^=97%, ten studies and ten comparison groups, Fig. S24) ^23,26–32,34,59^. Excluding five studies reporting outlying values ^23,30–32,59^ from the meta-analysis did not alter the interpretation, although the heterogeneity was moderately reduced (SMD=3.45 [2.91;4.00], I^2^=51%). The funnel plot showed a tendency toward a symmetrical pattern (Fig. S25) suggesting small-study effects.

The meta-analysis of lifestyle intervention studies also found a significant increase in circulating SHBG levels following adiposity loss (SMD=0.71 [0.34;1.07], I^2^=96%, 22 studies and 32 comparison groups, Fig. S26) ^20,33–36,38,39,41,43–55,57^. After the exclusion of outliers ^33,38,46–48,51,53,55^, the association remained statistically significant (SMD=0.46 [0.35;0.57], I^2^=74%). The funnel plot showed non-symmetrical patterns (Fig. S27) suggesting small-study effects. Two intervention studies on postmenopausal women reporting weight gain in the maintenance phase showed a significant decrease in SHBG levels ^35,53^. On the contrary, one intervention in premenopausal women did not find a significant change in SHBG after weight gain ^43^. A longitudinal study reported less than 10% increase in SHBG levels after an increase in IAAT in two years follow-up, but the study was weakly powered (n=18) ^60^. Stratified analysis tended to show higher significant inverse association in postmenopausal compared to premenopausal women, and with longer follow-up only (supplementary results).

***Observational***: Results in Fig. S28 indicated an inverse correlation between circulating SHBG and VAAT (pooled r=−0.47 [−0.58; −0.34], I2=0%, four studies and six comparison groups) ^41,82,96,110^. No significant correlations were observed with body fat percentage and fat mass (e.g., fat mass, pooled r= −0.09 [−0.26; 0.09], I2=32%, three studies four comparison groups) ^66,82,96^) while significant when pooling studies unadjusted correlation coefficients (e.g., fat mass, pooled r =−0.36 [−0.45; −0.27], I2=42%, ten studies and 12 comparison groups) ^52,65,70,79,80,90,95,102,106,110^). Ten studies performed linear regression to investigate the association between SHBG and adiposity but no meta-analysis was feasible ^11,68,74,79,87,88,99,105,112,114^, except for breast fat volume were the meta-analysis of adjusted standardized beta coefficients showed non-significant association with circulating SHBG (pooled beta=3.53 [−8.33;15.38], I2=100 %, Fig. S29) ^68,74,87^.

***Conclusion***: Strong evidence of probable inverse association as consistent results were observed in meta-analysis and the heterogeneity was rather low (surgery intervention, I2=51%; lifestyle intervention, I2=77%; observational, I2=0%). Relatively stronger associations were observed with high loss of body fatness, VAAT and in postmenopausal women.

##### DHEA(S)

A total of 13 intervention studies ^24–26,28–31,37,39–41,53,60^, and 19 observational studies evaluating direct measures of body fatness with DHEAS ^11,37,41,61,63,69,70,73,75,78,82,84,85,88,89,94,96,99,111^, as well as with DHEA ^70,74,82,84,88,89^ were included. No body-fluids-based studies were identified.

***Intervention:*** The meta-analysis on surgical interventions showed non-significant changes in circulating DHEAS concentration following a significant loss of adiposity (SMD=0.18 [−0.37;0.74], I^2^=90%, nine studies and ten comparison groups, Fig. S30) ^24–26,28–31,58,59^, and the exclusion of an outlier value ^25^ did not change the estimate (SMD=−0.10 [−0.34;0.15], I^2^=81%).

Meta-analysis of the effect estimates of the lifestyle intervention showed a non-significant change in circulating DHEAS after significant adiposity loss (SMD=0.05 [−0.34;0.44], I^2^=90%, five studies and six comparison groups, Fig. S31) ^37,39–41,53^ and no outlier detected. Non-significant change in DHEAS was also recorded in the maintenance phase (weight gain) of an intervention study ^53^ and a longitudinal study ^60^. Similarly, another study reported no significant change in circulating DHEAS levels after significant weight gain ^53^.

***Observational:*** Non-significant pooled correlation between DHEAS and fat mass (pooled r= −0.13 [−0.32; 0.06], I^2^=0%, two studies and three comparison groups) ^82,96^, body fat percentage (pooled r=−0.03 [−0.23; 0.17], I^2^=59%) ^75,82,111^, or VAAT (pooled r=−0.11 [−0.28; 0.06], I^2^=0%, three studies and four comparison groups) ^41,82,96^ was observed (Fig. S32). Adjusted beta coefficients reported in three studies ^11,88,99^ indicated no significant association between circulating DHEAS and six different adiposity measures (body fat percentage, central fat, fat mass, peripheral fat, SAAT, and VAAT) but no meta-analysis was feasible. Regarding DHEA, only one meta-analysis on unadjusted coefficients was possible and indicated a non-significant correlation between DHEA and SAAT (pooled r= −0.01 [−0.28; 0.26], I^2^=59%) ^70,84,89^. Three studies reported a non-significant adjusted beta coefficient value for the associations between DHEA and measures of adiposity ^74,84,88^.

***Conclusion:*** There is strong evidence of substantial effect unlikely on DHEAS. No conclusion can be drawn for circulating levels of DHEA.

### Other sex steroid markers

Sex steroid markers including sum of estrogen, estriol, dihydrotestosterone, free androgen index, and progesterone (including pregnanediol glucuronide), were identified in few studies, but the paucity of evidence and the high heterogeneity in the design of the studies hampered the conduct of meta-analysis. We narratively summarized the results of those studies in the supplementary results. Therefore, no conclusion can be drawn for these biomarkers.

## Discussion

This systematic review and meta-analysis of the effects of body fatness on sex steroid hormones included biomarkers measured in blood, urine and other excreted matrices, as well as in tissue, and investigated not only cross-sectional associations, but also longitudinal changes in body fat composition and its impact on hormone concentrations integrating evidence from weight loss interventions. Strong evidence of convincing positive association was found between body fat composition and postmenopausal circulating estrone. Strong evidence of probable positive association was found between body fat composition and level of testosterone (total and free), postmenopausal estradiol (total and free) and inverse association with SHBG. Strong evidence of a null association was observed for the association between body fat composition and premenopausal estradiol and estrone, as well as DHEA(S) and androstenedione. Limited evidence remained for estrogen metabolites, progesterone and its metabolites, PR, as well as for aromatase, and premenopausal free estradiol in their association with body fat composition. After stratification by menopausal status, the association between testosterone and body fatness, though significant in both groups, was found to be significantly higher in premenopausal compared to postmenopausal women. For SHBG, the inverse significant association was only found in postmenopausal women. This stratification was not possible for DHEA(S), free testosterone, and progesterone.

The association between circulating sex steroid hormones and BMI has already been investigated in previous studies. In premenopausal women, a pooled analysis of seven prospective studies found significant trends between BMI levels and sex steroids and showed that obese women (BMI>=30kg/m2) had a lower mean of estradiol after adjusting for menstrual cycle phase, luteal progesterone, and SHBG compared to leaner women (BMI<22.5 kg/m2). In contrast, higher concentrations were found for calculated free estradiol, estrone, DHEAS and free testosterone ^7^. Although a significant positive trend was reported for the association between testosterone levels and BMI, the heterogeneity between BMI subgroups was not significant. No association was reported for androstenedione levels and BMI. Our results partly based on longitudinal data from intervention studies, could not confirm those results due to the limited number of studies, except for the positive association observed between BMI and testosterone levels, the inverse association with SHBG, and the null association with androstenedione. In postmenopausal women, a report of 18 pooled prospective studies comparing hormone concentrations in three different assay types (extraction, direct and mass spectrometry assays) found a strong and linear positive association between concentration of estradiol and estrone with BMI, and a rather moderate positive association between testosterone and BMI, in each assay type ^8^. Additionally, a review on cross-sectional studies concluded an inverse association between plasma levels of DHEA and BMI based on four studies (three on pre- and one on postmenopausal women), whereas the association with DHEAS based on 11 studies was not significant (6 pre- and 5 postmenopausal women) ^168^.

A recent mendelian randomisation was in line with our results indicating that SHBG was rather protective against general adiposity, while hyperandrogenism (elevated Bio testosterone and FAI) was linked to increased central/visceral obesity and lower total body adiposity ^115^. In addition to confirming the results observed in previous pooled analysis, the current meta-analysis extends the evidence by incorporating tissue-based markers and weight loss interventions. These findings are relevant for prevention, as they suggest that sex steroid levels can be modified with changes in body fatness through preventive clinical or behavioural interventions.

Several reports have also shown the association of sex steroid hormone concentrations with the risk of developing cancers. The pooled analysis of seven prospective studies on premenopausal women, which we mentioned earlier, demonstrated higher levels of testosterone to be positively associated with breast cancer risk, while progesterone, and SHBG were not ^7^. In postmenopausal women, a strong positive association between estradiol, estrone and testosterone and breast cancer risk was observed ^8^, and inverse with SHBG, specifically in ER+ breast cancer, while in ER− subtype, a rather positive association was observed ^169^. Estrogen is also shown to have a critical role in endometrial cancer etiology ^170^.

Compared to breast cancer risk, a stronger magnitude of association has been observed between circulating estradiol levels and a risk of developing endometrial cancer at menopause. Higher levels of estrone, and testosterone (total and free), and lower levels of SHBG, were associated with increased risk of endometrial cancer. Associations were not significant with androstenedione and DHEAS ^171^, and in premenopausal women. Sex steroids were also found to be associated with ovarian cancer showing a 25% higher risk when comparing highest to lowest tertiles of circulating testosterone ^172^. This association tended to mainly occur in type I ovarian cancer (endometrioid and mucinous tumours subtypes). No associations were reported for other sex steroids except when stratified by menopausal status, a positive association was observed with androstenedione and SHBG in premenopausal women. Results from the UK Biobank show that in postmenopausal women, higher level of bioavailable testosterone was inversely associated with colorectal cancer ^173^ although this association was rather borderline significant in another study ^174^. Similarly, higher levels of SHBG were positively associated with colorectal cancer risk ^174^. Estradiol concentrations could not be investigated due to high proportion below the limit of detection ^175^.

Aside from cancer outcomes, sex steroids have also been found associated with obesity-related diseases. In a prospective study on postmenopausal women (Multi-Ethnic Study of Atherosclerosis), higher levels of testosterone were found to be associated with increased risk of cardiovascular disease and coronary heart disease, while higher levels of estradiol were associated with a lower risk of coronary heart disease ^176^. A significant inverse association was also reported in a meta-analysis of 13 cohorts which showed a lower level of SHBG with increased risk of type 2 diabetes, whereas higher levels of estradiol were associated with an increased risk of type 2 diabetes in postmenopausal women ^177^. No associations were found for other hormones (testosterone total and free).

The strengths of the current systematic literature review, on the one hand, was the comprehensive search with strictly defined inclusion and exclusion criteria ensuring a high-quality body of evidence. Only studies exclusively on healthy women, and with blood/body fluid collected during the same menstrual cycle phase (or corrected for menopausal status) were included. For the intervention studies, a significant weight loss after a minimum of 3-months follow-up was required for inclusion. The inclusion of surgical weight loss intervention studies allowed investigation of an abrupt and substantial reduction in body fatness, relative to the more gradual and less pronounced reductions observed in behavioural lifestyle interventions.

The conduct of meta-analysis by menopausal status was possible, although the strict inclusion criteria have led to a smaller number of included studies for certain markers and hampered more sub-group analysis. Another asset of this study was to integrate, in addition to the circulating markers, all types of study investigating biomarkers measured in biomaterial other than blood, though it should be noted that these studies were conducted across a wide range of tissue types (e.g., adipose, breast, endometrial, skin) or urine and other excreted matrices (e.g., saliva, breast ductal lavage) which hampered meta-analysis due to the heterogeneity.

Limitations were also noted in this systematic review, notably the small number of studies included for some biomarkers (progesterone, estrogen metabolite, but also in premenopausal estradiol, free estradiol and estrone), the small number of participants in some studies (<30), and the heterogeneity in method and adjustment used across studies to measure associations which limited the capacity to quantitatively synthesize them through meta-analysis. However, we conducted numerous meta-analyses for several of the biomarkers and comprehensively synthesized a large body of evidence linking adiposity with sex steroids.

Additionally, for intervention studies, it can be noted that the change in weight loss was heterogeneous between studies, as duration of follow-up widely varies (3 to 48 months for surgical, and 3 to 13.5 months for lifestyle interventions). The high heterogeneity was also noticeable in the meta-analysis, which remained high despite excluding outliers. The large heterogeneity of studies is also due to the difference in laboratory assay methods, and specimen (e.g., serum or plasma). Therefore, the final judgment of evidence according to the CUP criteria was mainly guided by the consistency of evidence generated from the meta-analysis of intervention studies without accounting for heterogeneity. Meta-analysis on observational studies, and support from studies on urine and other excreted matrices and tissue were considered to upgrade or downgrade the level of evidence.

## Conclusion

To our knowledge, this is the first comprehensive systematic review and meta-analysis that integrated evidence from both interventional and observational studies, synthesizing data on a wide range of sex steroid hormones measured across multiple biological matrices including blood, tissue, and urine and other excreted samples. In addition to adjusted cross-sectional associations it also examined longitudinal changes in body fat composition and their impact on hormone concentrations. The findings indicate that higher body fatness is associated with lower levels of SHBG and higher levels of testosterone, and free testosterone, as well as increased concentrations of estrone, estradiol and free estradiol in postmenopausal. Evidence for estrogen metabolites, progesterone and its metabolites, PR, and premenopausal free estradiol were inconclusive. In contrast, strong evidence suggests no substantial effect of body fatness on DHEA, androstenedione, and premenopausal estrone and estradiol. Since sex steroid hormones are associated with various diseases, our findings reveal that targeted weight loss interventions significantly alter sex steroid hormone concentrations, not just in blood, but also in body fluids and tissues, offering a direct strategy to prevent sex-steroid-related cancers and endocrine diseases.

## Data Availability

Data accessibility: Data was extracted from published scientific papers which are available and accessible.

## Authors contribution

RTF conceived and designed the study. CLC coordinated data collection and drafted the manuscript. HS, CLC and MK collected data and conducted the analysis. All authors critically revised and approved the manuscript for submission.

## Ethics Statement

The authors have nothing to report.

## Data accessibility

Data was extracted from published scientific papers which are available and accessible.

## Acknowledgments

We would like to acknowledge Lieke Lanjouw, Sofia Schüssler, Norah A. Burchardt, Jasmin Ostermann, Eva Dichiser, Nina Decker, Alyssa Vaziri, Florian Karpa, and Jane Mary Luis for their support during the title abstract screening, full text screening and data extraction phases of the review.

