## Supplementary files for "Adiposity and sex steroid hormones: evidence on direct measures and changes over time -- a systematic review and meta-analysis"

### Contents of the Supplementary File

### Supplementary Results

#### Studies characteristics

Some of the included studies had different characteristics; One intervention study has two arms, one surgical and the other dietary intervention (1), two non-randomized dietary intervention studies are included in both meta-analysis of intervention and observational studies since they also reported cross-sectional associations between direct measure of body fatness and sex steroids, as well as prospective data on weight change and changes on sex steroids (2, 3). Two non-randomized intervention studies without relevant data on pre- and post-intervention but for which baseline correlations were available (4, 5) were analyzed with observational studies on direct measure of asiposity. One lifestyle intervention study had estrogens measured 3 months apart in blood and ductal fluid specimens (6).

Additionally, three interventions (7-9), one cohort (10) and one cross-sectional (11) studies reported weight gain, or regain after intervention, and therefore were only summarized narratively.

Studies not amendable to meta-analysis included those that reported only p-value of correlation coefficient (12), as well as having heterogeneity on methodological, adjustments factors, biomarker measurements, and/or fat type (13-25).

#### Other sex steroid markers

Circulating sex steroid markers for which studies identified had high between-study heterogeneity to conduct a meta-analysis or less than 3 studies, are presented below:

**Total estrogen**

In postmenopausal women, the study from De Waard et al. showed significant weak correlation between weight changes and total estrogen (sum of E1+E2+E3) measured in urine (r=0.34, p=0.01) (26).

**Estriol (E3)**

A total of five studies investigated estriol levels in urine with body fat type (26-30).

In premenopausal women, one physical activity intervention (27), and one cross-sectional (28) studies found non-significant association between body fatness and urinary estriol measured during follicular phase. In luteal phase, no significant association was also observed in two cross-sectional studies (29, 30).

In postmenopausal women, one dietary intervention study found a modest correlation between BMI and urinary estriol (r=0.27, p=0.05) and weight loss and a decrease in urinary estriol (r=0.32, p=0.018) at baseline, but not at follow-up (26).

**Dihydrotestosterone (DHT)**

A total of four observational studies on circulating DHT (12, 31-33) were identified. Only studies evaluating unadjusted correlations were identified. The meta-analysis of unadjusted correlation coefficients was significant (r= -0.35 [-0.52; -0.14], I2=0%, 3 studies) (31-33).

**Free androgen index (FAI)**

A total of seven studies (9, 12, 23, 34-36) were identified with circulating FAI in relation to adiposity. Four intervention and three observational studies.

One surgical intervention study (34) with two arms (Pre- and postmenopausal women) measured FAI. A significant association between decrease in BMI, waist and hip circumference and decrease in FAI was found in both study arms.

In lifestyle intervention studies, a significant association was found between decrease in FAI levels and decreased in fat mass, percent body fat; and weight (35). And another found a significant association between decrease in FAI and loss in BMI, weight, percent body fat and VAT (9). Significant increase in FAI was also observed after weight regain (9).

Three studies reported the association of free androgen index with four different direct adiposity measures (12, 23, 36). Only one study investigated the associations by performing multiple adjusted linear regression analysis and showed a significant positive association between FAI and body fat percentage (23). The other studies reported unadjusted correlations between FAI and four different type of adiposity.

**Progesterone**

A total of two intervention studies (37, 38), five observational studies (17, 25, 39-42) were included. Four studies reported urine-based pregnanediol glucuronide (PdG), the main metabolite of progesterone (43-46).

*Premenopausal women*

Intervention: One surgical intervention study found not significant change in luteal progesterone concentration 12 months after vertical banded gastroplasty (37), while one 4-months physical activity intervention leading to significant decrease in fat mass and body fat percentage showed a significant decrease in follicular progesterone (38).

Observational: One study on women in the ovulation phase reported non-significant difference in progesterone between extreme tertiles of trunk/leg fat ratio when adjusted for age, race, energy intake and physical activity (-6.1%, ptrend=0.09) (39). In follicular phase, a significant inverse association between VAAT and circulating progesterone was observed in a linear regression model adjusted for age and BMI, while the association with TAAT and SAAT were not significant (40). Three other studies investigating the relationship with breast fat volume and showed non-significant associations with follicular (41, 42) and luteal circulating progesterone (25, 42).

Urine and other excreted matrices: One surgical intervention study showed an increase of luteal PdG after more than 25% weight loss over a 6-months follow-up (43). Similarly, a cohort reported inverse associations between adiposity and PdG levels in under- and overweight women all the menstrual cycle phases (follicular, P peak, luteal) (46). Two cross-sectional studies reported that luteal PdG levels were higher in obese than normal weight women (44, 45).

*Postmenopausal women*

Observational: One cross-sectional study found no significant association between body fat percentage and progesterone using linear regression adjusted for reproductive factors and BMI (17).

#### Stratified analysis

**Testosterone**

When stratified by menopausal status, the meta-analysis of lifestyle intervention studies showed significant SMDs in both pre- and postmenopausal women (SMD=-0.33 [-0.58;-0.09] seven studies and eight comparison groups (3, 9, 35, 38, 47-49), and SMD=-0.22 [-0.40;-0.03] 6 studies and eleven comparison groups (7, 50-54), respectively). When stratified by length of follow-up, the average weight loss was 8.8% after six months vs. 8.7% after 12 months. The corresponding changes of testosterone levels were significant (SMD=-0.49 [-0.93;-0.05]; I2=94%, three studies with four comparison groups (3, 7, 35)) after six months, while not after 12 months (SMD=-0.09 [-0.20;0.01], I2=0%, one study with three comparison groups (51)). However, only one study with three arms was included in the analysis for the 12-month follow-up (51).

**Free testosterone**

Lifestyle intervention studies showed non-significant SMDs in premenopausal women (SMD=-0.72 (-1.79;0.34), I2=95%, four studies (3, 38, 47, 49)) while a borderline significant association was observed in postmenopausal women (SMD=-0.50 (-0.99;0.00), I2=97%, four studies and nine comparison groups (7, 50, 51, 53)).

**Androstenedione**

Meta-analysis of lifestyle intervention studies showed a non-significant association in premenopausal women (SMD=-0.28 [-1.04; 0.47], I2=92%, two studies and three comparison groups (3, 35)) and a borderline significant in postmenopausal women (SMD=-0.07 [-0.14;-0.00], I2=0%, three studies and seven comparison groups (51-53)) following a statistically significant weight reduction at 6 and 12 months. The associations between change in body fatness measure and androstenedione were not statistically significant in both follow-up periods (6-months, SMD=-0.28 [-1.04; 0.47], I2=92%, two studies with three comparison groups (3, 35); 12-months, SMD=-0.03 [-0.13;0.08], I2=0%, one study with three comparison groups (51)).

**SHBG**

Meta-analysis of lifestyle intervention studies showed a non-statistically significant association in premenopausal women (SMD=0.43 [-0.02; 0.88], I2=86%, seven studies with nine comparison groups (1, 3, 9, 35, 38, 47, 55)) and a significant association in postmenopausal women (SMD=0.75 [0.20;1.31], I2=0%, thirteen studies with nineteen comparison groups (7, 8, 50-54, 56-61)) following a significant weight loss at 6 and 12 months. Lifestyle intervention studies showed non-significant associations in the 6-months follow-up (SMD=1.01 [-0.30; 2.32], I2=98%, five studies with seven comparison groups (3, 7, 35, 56, 60)), while a significant SHBG increase was observed in the 12-months follow-up (SMD=0.40 [0.24;0.57], I2=86%, five studies with nine comparison groups (1, 8, 51, 58, 62)).

### Supplementary tables

#### Supplementary table S1: Search Body fatness to sex steroids in PubMed

| S.No | Search terms |
| --- | --- |
| #1 | (body fatness[Title/Abstract] OR adiposity[Title/Abstract] OR obesity[Title/Abstract] OR obese [Title/Abstract] OR abdominal obesity[Title/Abstract] OR morbid obesity[Title/Abstract] OR metabolic obesity[Title/Abstract] OR overweight[Title/Abstract] OR over-weight [Title/Abstract] OR over weight [Title/Abstract] OR BMI[Title/Abstract] OR body mass index[Title/Abstract] OR waist-Hip ratio[Title/Abstract] OR waist to hip ratio[Title/Abstract] OR waist-height ratio[Title/Abstract] OR waist to height ratio[Title/Abstract] OR waist hip ratio*[Title/Abstract] OR intra-abdominal fat[Title/Abstract] OR adipocytes[Title/Abstract] OR adipose tissue[Title/Abstract] OR weight loss[Title/Abstract] OR weight gain[Title/Abstract] OR anthropometry[Title/Abstract] OR body composition [Title/Abstract] OR body mass [Title/Abstract] OR skinfold measurement* [Title/Abstract] OR skinfold thickness[Title/Abstract] OR bio-impedence[Title/Abstract] OR waist circumference[Title/Abstract] OR hip circumference[Title/Abstract] OR body composition[Title/Abstract] OR body constitution[Title/Abstract]) |
| #2 | Estrogens[Title/Abstract] OR Estrogen[Title/Abstract] OR Oestrogens[Title/Abstract] OR Oestrogen*[Title/Abstract] OR Oestrogen*[Title/Abstract] OR Estrone[Title/Abstract] OR Oestrone[Title/Abstract] OR Estradiol[Title/Abstract] OR Oestradiol[Title/Abstract] OR Estrone[Title/Abstract] OR Oestrone[Title/Abstract] OR sex hormone*[Title/Abstract] OR sex steroid*[Title/Abstract] OR estriol[Title/Abstract] OR Oestriol [Title/Abstract] OR estrogen metabolites[Title/Abstract] OR androgens[Title/Abstract] OR androgen metabolites[Title/Abstract] OR dehydroepiandrosterone[Title/Abstract] OR androstenedione[Title/Abstract] OR androstenediol[Title/Abstract] OR androsterone[Title/Abstract] OR testosterone[Title/Abstract] OR progesterone[Title/Abstract] OR sex hormone binding globulin[Title/Abstract] OR sex hormone-binding globulin[Title/Abstract] OR estrogen receptor[Title/Abstract] OR progesterone receptor[Title/Abstract] OR hormone receptor[Title/Abstract] OR estrobolome[Title/Abstract] |
| #3 | #1 AND #2 |
| #4 | (animal[MeSH Terms] NOT human[MeSH Terms]) |
| #5 | (animal[Title/Abstract]NOT human[Title/Abstract]) |
| #6 | #4 OR #5 |
| #7 | #3 NOT #6 |
| #8 | (Corrigendum[Title/Abstract] OR letter[Title/Abstract] OR comment[Title/Abstract] OR erratum[Title/Abstract] OR Comment in[Title/Abstract] OR Comment on[Title/Abstract] OR Erratum in[Title/Abstract] OR Erratum for[Title/Abstract] OR Expression of concern in[Title/Abstract] OR Expression of concern for[Title/Abstract]) |
| #9 | #7 NOT #8 |
| #10 | (narrative review*[Title/Abstract] OR systematic review*[Title/Abstract] OR scoping review*[Title/Abstract] OR review*[Title/Abstract] OR meta-analys*[Title/Abstract] OR meta analys*[Title/Abstract] OR metaanalys*[Title/Abstract]) |
| #11 | #9 NOT #10 |
| #12 | (book[Title/Abstract] OR books[Title/Abstract] OR pmcbook[Title/Abstract] OR book chapters[Title/Abstract] OR pmcbooktitle[Title/Abstract] OR pmcbookchapter[Title/Abstract] OR chapter[Title/Abstract] OR chapters[Title/Abstract] OR chapter*[Title/Abstract]) |
| #13 | #11 NOT #12 |
| #14 | #11 with language filter: English |
| #15 | #11 with language filter: German |
| #16 | #11 with language filter: French |
| #17 | #11 with language filter: Dutch |
| #18 | #14 OR #15 OR #16 OR #17 |
| #19 | #13 with age filter: 19+ years |

Supplementary table S2: Search Body fatness to sex steroids in Web Of Science

| | |
|---|---|
| #1 | (TI=(body fatness OR adiposity OR obesity OR obese OR abdominal obesity OR morbid obesity OR metabolic obesity OR overweight OR over-weight OR over weight OR BMI OR body mass index OR waist-Hip ratio OR waist to hip ratio OR waist-height ratio OR waist to height ratio OR waist hip ratio* OR intra-abdominal fat OR adipocytes OR adipose tissue OR weight loss OR weight gain OR anthropometry OR body composition OR body mass OR skinfold measurement* OR skinfold thickness OR bio-impedence OR waist circumference OR hip circumference OR body composition OR body constitution) OR<br>AB=(body fatness OR adiposity OR obesity OR obese OR abdominal obesity OR morbid obesity OR metabolic obesity OR overweight OR over-weight OR over weight OR BMI OR body mass index OR waist-Hip ratio OR waist to hip ratio OR waist-height ratio OR waist to height ratio OR waist hip ratio* OR intra-abdominal fat OR adipocytes OR adipose tissue OR weight loss OR weight gain OR anthropometry OR body composition OR body mass OR skinfold measurement* OR skinfold thickness OR bio-impedence OR waist circumference OR hip circumference OR body composition OR body constitution)) *AND*<br>**LANGUAGE:** (English OR Dutch OR French OR German) *AND* **DOCUMENT TYPES:** (Article)<br>*Indexes=SCI-EXPANDED Timespan=All years* |
| #2 | TI=(Estrogens OR Estrogen OR Oestrogens OR Oestrogen* OR Oestrogen* OR Estrone OR Oestrone OR Estradiol OR Oestradiol OR Estrone OR Oestrone OR sex hormone* OR sex steroid* OR estriol OR Oestriol OR estrogen metabolites OR androgens OR androgen metabolites OR dehydroepiandrosterone OR androstenedione OR androstenediol OR androsterone OR testosterone OR progesterone OR sex hormone binding globulin OR sex hormone-binding globulin OR estrogen receptor OR progesterone receptor OR hormone receptor OR estrobolome) OR<br>AB=(Estrogens OR Estrogen OR Oestrogens OR Oestrogen* OR Oestrogen* OR Estrone OR Oestrone OR Estradiol OR Oestradiol OR Estrone OR Oestrone OR sex hormone* OR sex steroid* OR estriol OR Oestriol OR estrogen metabolites OR androgens OR androgen metabolites OR dehydroepiandrosterone OR androstenedione OR androstenediol OR androsterone OR testosterone OR progesterone OR sex hormone binding globulin OR sex hormone-binding globulin OR estrogen receptor OR progesterone receptor OR hormone receptor OR estrobolome)) *AND*<br>**LANGUAGE:** (English OR Dutch OR French OR German) *AND* **DOCUMENT TYPES:** (Article)<br>*Indexes=SCI-EXPANDED Timespan=All years* |
| #3 | (#1 AND #2) *AND*<br>**LANGUAGE:** (English OR Dutch OR French OR German)<br>*Indexes=SCI-EXPANDED Timespan=All years* |
| #4 | (TS=(human OR humans OR subject* OR patient* OR child* OR participant OR participants OR cohort OR population OR adult* OR individual* OR women OR woman OR men OR elderly OR adolescents OR female OR females OR male OR males OR people OR person OR maternal OR youth OR girls OR boys OR controls OR control OR survivors OR african americans OR elders OR black OR chinese OR white OR americans OR american OR fetal OR south asians OR asians OR westerners OR european* OR caucasian* OR runners OR families OR twins OR employe* OR worker* OR teachers OR nurses OR volunteers)) *AND*<br>**LANGUAGE:** (English OR Dutch OR French OR German)<br>*Indexes=SCI-EXPANDED Timespan=All years* |
| #5 | (#3 AND #4) *AND*<br>**LANGUAGE:** (English OR Dutch OR French OR German)<br>*Indexes=SCI-EXPANDED Timespan=All years* |
| #6 | ((TS=(animal models OR animal OR animals OR disease models OR Animal Diseases OR Disease Models Animal OR Models Animal OR canine OR canines OR dog OR dogs OR feline OR hamster OR hamsters OR lamb OR lambs OR mice OR mouse OR monkey* OR monkeys OR murine OR murins OR murinae OR pig OR pigs OR guinea pig OR guinea pigs OR piglet OR piglets OR porcine OR porcines OR primate OR primates OR |

| | |
|---|---|
| | rabbit OR rabbits OR rat OR rats OR rodent OR rodents OR gorilla OR gorillas OR sheep OR sheeps OR fish OR drosophila OR veterinary* OR macaca OR macaque OR horse OR horses))) *AND*<br>**LANGUAGE:** (English OR Dutch OR French OR German)<br>*Indexes=SCI-EXPANDED Timespan=All years* |
| #7 | (#5 NOT #6) *AND* **LANGUAGE:** (English OR Dutch OR French OR German)<br>*Indexes=SCI-EXPANDED Timespan=All years* |
| #8 | ((TS= (Corrigendum OR letter OR comment OR erratumOR Comment in OR Comment on OR Erratum in OR Erratum for OR Expression of concern in OR Expression of concern for))) *AND*<br>**LANGUAGE:** (English OR Dutch OR French OR German)<br>*Indexes=SCI-EXPANDED Timespan=All years* |
| #9 | (#7 NOT #8) *AND*<br>**LANGUAGE:** (English OR Dutch OR French OR German)<br>*Indexes=SCI-EXPANDED Timespan=All years* |
| #10 | (TS=(narrative review* OR systematic review* OR scoping review OR review* OR meta-analys* OR meta analys* OR metaanalys*)) *AND*<br>**LANGUAGE:** (English OR Dutch OR French OR German)<br>*Indexes=SCI-EXPANDED Timespan=All years* |
| #11 | (#9 NOT #10) *AND*<br>**LANGUAGE:** (English OR Dutch OR French OR German)<br>*Indexes=SCI-EXPANDED Timespan=All years* |
| #12 | (#11) *AND*<br>**LANGUAGE:** (English OR Dutch OR French OR German) *AND*<br>**DOCUMENT TYPES:** (Art Exhibit Review OR Bibliography OR Biographical-Item OR Book OR Book Chapter OR Book Review OR Chronology OR Correction OR Correction, Addition OR Dance Performance Review OR Data Paper OR Database Review OR Discussion OR Editorial Material OR Excerpt OR Fiction, Creative Prose OR Film Review OR Hardware Review OR Item About an Individual OR Letter OR Meeting Abstract OR Meeting Summary OR Music Performance Review OR Music Score OR Music Score Review OR News Item OR Note OR Poetry OR Proceedings Paper OR Record Review OR Retracted Publication OR Retraction OR Review OR Script OR Software Review OR TV Review, Radio Review OR TV Review, Radio Review Video OR Theater Review)<br>*Indexes=SCI-EXPANDED Timespan=All years* |
| #13 | (#11 NOT #12) *AND*<br>**LANGUAGE:** (English OR Dutch OR French OR German)<br>*Indexes=SCI-EXPANDED Timespan=All years* |

#### Supplementary table S3: Sex steroids units.

| Biomarkers | Harmonized units |
|---|---|
| Testosterone total | nmol/L |
| Testosterone free | pmol/L |
| Testosterone bio | nmol/L |
| Estrone | pmol/L |
| Estradiol | pmol/L |
| SHBG | nmol/l |
| DHEA(S) | nmol/l |
| Androstendione | pmol/l |
| Progesterone | nmol/L |

SHBG: Sex hormone-binding globulin, DHEA(S): Dehydroepiandrosterone (Sulfate)

#### Supplementary table S4: Summary table of Studies included.

| **Anthor, year, PMID, country, study design** | **Population, age, BMI, menopausal status** | **Comparaison groups, Biomarkers** | **Main findings, Adjustments** |
|---|---|---|---|
| Surgical interventions | | | |
| Mingrone 2002<br>(1)<br>Italy<br>RCT<br>Follow-up: 12 months | Sample size n= Surgical intervention: 21<br><br>Obese women, diet intervention, BMI: 48.4 (SD = 8.9), surgical intervention, BMI: 48.3 (SD = 6.3); Age: 30-45<br><br>Pre-menopause<br>(Follicular) | Exposures<br>Fat mass (kg, DEXA), Weight (kg), BMI (kg/m^2)<br><br>Outcomes<br>SHBG (nmol/l, Immunoassay) | Change Fat mass: -39.61 (Mean = 65.9 (SD = 10.2) to 39.8 (SD = 12.7))<br>Change Weight: -28.01 (Mean = 125.3 (SD = 12.8) to 90.2 (SD = 15))<br>Change BMI: -27.12 (Mean = 48.3 (SD = 6.3) to 35.2 (SD = 7.6))<br>SHBG: Mean = 16.3 (SD = 5.5) to 39.7 (SD = 17.6) |
| Friedman 1982<br>(63)<br>USA<br>Non-randomized intervention<br>Follow-up: Not reported. But until they reach stable weight loss mean of 7.9 months | Sample size n= 15<br><br>morbidly obese, in excess of 50% over ideal body weight;<br>Age: 35.2 (SD=7)<br><br>Pre-menopause<br>(Unknown) | Exposures<br>Weight (kg)<br><br>Outcomes<br>Free Testosterone (pg/ml, RIA)<br>SHBG (nM, charcoal absorption assay)<br>Testosterone (ng/dl, RIA) | Change Weight: -17.57 (Mean = 110.4 (SD = 17) to 91 (SD = 16)<br>free Testosterone: Mean = 9.7 (SD = 4.3) to 7.3 (SD = 2.8)<br>Testosterone: Mean = 35.8 (SD = 12.7) to 35.7 (SD = 10.4)<br>SHBG: Mean = 20.8 (SD = 11.1) to 32.6 (NR) |
| Di Carlo 1999<br>(64)<br>Italy<br>Non-randomized intervention<br>Follow-up: 3 months | Sample size n= 8<br><br>Severely obese women in whom amenorrhea developed after the procedure, BMI: 47 (SD = 4.4);<br>Age: 26.9 (SD= 5.3)<br><br>Pre-menopause<br>(Unknown) | Exposures<br>Weight (kg), BMI (kg/m^2)<br><br>Outcomes<br>Androstendione (ng/ml, RIA) Estradiol (pg/ml, RIA) Testosterone (ng/ml, RIA)<br>DHEAS (µg/dl, RIA) | Change Weight: -25.29 (Mean = 130.1 (SD = 9.8) to 97.2 (SD = 9))<br>Change BMI: -25.32 (Mean = 47 (SD = 4.4) to 35.1 (SD = 3.8))<br>Androstenedione: Mean = 157.4 (SD = 18.2) to 158.6 (SD = 22.3)<br>Testosterone: Mean = 0.5 (SD = 0.1) to 0.5 (SD = 0.1)<br>DHEAS: Mean = 118.2 (SD = 19.8) to 125 (SD = 17.4) |
| Savastano 2005<br>(65)<br>Italy<br>Non-randomized intervention<br>Follow-up: 24 (followup also at 6 & 12) months | Sample size n= 30<br><br>Healthy but morbid obese women, BMI: 37- 62;<br>Age: 36.7 (SD=6.4)<br><br>Pre-menopause<br>(Follicular) | Exposures<br>BMI (kg/m^2), Fat mass (kg, BIA), Weight (kg)<br><br>Outcomes<br>DHEAS (µmol/l, RIA), Cortisol/DHEAS molar ratio (calculated from cortisol and DHEAS values) | Baseline to 6 Month<br>Change fat mass: -28.77 (Mean = 64.3 (SD = 13.4) to 45.8 (10.3))<br>Change BMI: -15.5 (Mean = 48.4 (SD = 7.1) to 40.9 (5.3))<br>Change Weight: -15.12 (Mean = 126.3 (SD = 17.3) to 107.2 (13.7))<br>DHEAS: Mean = 2.7 (SD = 0.8) to 5 (0.6)<br>Cortisol/DHEAS ratio: Mean = 198.5 (SD = 59.5) to 95.7 (13.1)<br><br>Baseline to 12 Month<br>Change fat mass: -45.88 (Mean = 64.3 (SD = 13.4) to 34.8 (4.9))<br>Change BMI: -25.83 (Mean = 48.4 (SD = 7.1) to 35.9 (3.6))<br>Change Weight: -25.73 (Mean = 126.3 (SD = 17.3) to 93.8 (7.3))<br>DHEAS: Mean = 2.7 (SD = 0.8) to 5 (0.5) |

| | | | |
|---|---|---|---|
| | | | Cortisol/DHEAS ratio: Mean = 198.5 (SD = 59.5) to 94.4 (9.7)<br><br>Baseline to 24 Month<br>Change fat mass: -49.77 (Mean = 64.3 (SD = 13.4) to 32.3 (3.3))<br>Change BMI: -29.13 (Mean = 48.4 (SD = 7.1) to 34.3 (3))<br>Change Weight: -28.42 (Mean = 126.3 (SD = 17.3) to 90.4 (6))<br>DHEAS: Mean = 2.7 (SD = 0.8) to 5 (0.4)<br>Cortisol/DHEAS ratio: Mean = 198.5 (SD = 59.5) to 95.6 (7.9) |
| Kopp 2006<br>(66)<br>Austria<br>Non-randomized intervention<br>Follow-up: 8-26 months | Sample size n= 43<br><br>Severly obese women, BMI: 48 (SD=7);<br>Age: 41 (SD=7)<br><br>Pre-menopause<br>(Unknown) | Exposures<br>BMI (kg/m^2), Weight (kg)<br><br>Outcomes<br>Free Androgen Index (calculated as Total Testosterone/SHBG ratio)<br>SHBG (nmol/l, ELISA)<br>Androstendione (ng/ml, RIA)<br>Testosterone (nmol/l, electrochemiluminescence nce Immunoassay)<br>DHEAS (µmol/l, ELISA) | Change BMI: -31.25 (Mean = 48 (SD = 7) to 33 (6))<br>Change Weight: -33.08 (Mean = 133 (SD = 20) to 89 (16))<br><br>Free Androgen Index: Mean = 0.2 (SD = 0.1) to 0 (0)<br>SHBG: Mean = 17 (SD = 12) to 70 (30)<br>Androstenedione: Mean = 2.1 (SD = 0.8) to 1.4 (0.5)<br>Testosterone: Mean = 1.9 (SD = 0.8) to 1.2 (0.6)<br>DHEAS: Mean = 1.7 (SD = 0.9) to 1.5 (0.8)<br><br>stat used for the association = paired t-test, correlation coefficients with Bonferroni-Holm correction and univariate linear regression, Pearson correletion: change BMI and Free Androgen Index = 0.07, change BMI and SHBG = -0.5, change BMI and Androstendione = 0.09, change BMI and Testosterone = 0.28, change BMI and DHEAS = -0.03 |
| Paul 2020<br>(67)<br>Sweden<br>Non-randomized intervention<br>Follow-up: 12 months | Sample size n= 68<br><br>Healthy women, BMI: 47 (IQR = 32 to 69);<br>Age: median=37 (range=20-48)<br><br>Pre-menopause<br>(Mix) | Exposures<br>Weight (kg), BMI (kg/m^2)<br><br>Outcomes<br>Testosterone (nmol/l, electro-chemiluminescence immunoassay with competitive principle)<br>SHBG (nmol/l, electro-chemiluminescence immunoassay with sandwich principle) | Change Weight: -29.71 (Mean = 116.67 (IQR = 76 to 163) to 82 (IQR = 49 to 122))<br>Change BMI: -36.88 (Mean = 47 (IQR = 32 to 69) to 29.67 (IQR = 21 to 41))<br>Testosterone: Mean = 1.1 (SD = 0.6) to 0.8 (0.4)<br>SHBG: Mean = 33.3 (SD = 11.1) to 71.7 (26.7) |
| Bastounis 1998<br>(37)<br>Greece<br>Non-randomized intervention<br>Follow-up: 12 months | Sample size n= 38<br><br>Morbidly obese patients, BMI>40;<br>Age: 34.3 (SD=5.9)<br><br>Pre-menopause<br>(Luteal) | Exposures<br>Weight (kg), BMI (kg/m^2)<br><br>Outcomes<br>Estradiol (pg/ml, RIA)<br>Progesterone (ng/ml, RIA)<br>Testosterone (ng/ml, RIA) | Change Weight: -39.9% (Mean = 148 (SD = 21) to 88.9 (12.7))<br>Change BMI: -39.9% (Mean = 56.7 (SD = 7.7) to 34.1 (4.8))<br><br>Estradiol: -22.4% (Mean = 94.9 (SD = 34.3) to 73.6 (26.8)), S<br>Progesterone: 22% (Mean = 3.6 (SD = 2.5) to 4.4 (2.7)), NS<br>Testosterone: -40% (Mean = 0.5 (SD = 0.3) to 0.3 (0.2)), S<br>Free Testosterone:-36.0 (Mean = 2.5 (SD = 1.3) to 1.6 (0.9)), S<br>Androstenedione:-16.7 (Mean = 1.8 (SD = 0.9) to 1.5 (0.4)), S<br>DHEAS: -9% (Mean = 222 (SD = 147) to 202 (150)), NS<br>SHBG: 93.3% (Mean = 32.8 (SD = 10.8) to 63.4 (20.7)), S |

| | | | |
|---|---|---|---|
| | | Free Testosterone (pg/ml, RIA)<br>Androstendione (ng/ml, RIA)<br>DHEAS (µg/dl, RIA)<br>SHBG (nmol/l, IRMA) | |
| Ernst 2013<br>(34)<br>Switzerland<br>Non-randomized intervention<br>Follow-up: 12 months | Sample size n= 36, premenopausal: 30, postmenopausal: 6<br><br>Severly obese women, BMI: 38.7-55.1;<br>Age: 41.2 (SD=9.6)<br><br>Pre (Unknown) and Post-menopausal (different arms) | Exposures<br>BMI (kg/m^2)<br>Waist circumference (cm)<br>Hip circumference (cm)<br><br>Outcomes<br>SHBG (nmol/l, chemiluminescence immunoassay), Testosterone (nmol/l, chemiluminescence immunoassay), Bioavailable Testosterone (nmol/l, calculated by using the method of Vermeulen et. al method), Free Testosterone (nmol/l, chemiluminescence immunoassay), DHEAS (µmol/l, chemiluminescence immunoassay), Free Androgen index (calculated as the total testosterone/SHBG ratio x 100) | Premenopausal women:<br>Change BMI: -40.69 (Mean = 46.7 (SD = 4.3) to 27.7(3.3))<br>Change waist circumference: -26.48 (Mean = 125.4 (SD = 12) to 92.2 (12))<br>Change hip circumference: -25.57 (Mean = 140 (SD = 13.7) to 104.2 (11.5))<br><br>SHBG: Mean = 30.5 (SD = 13.7) to 71.9 (35.6)<br>Testosterone: Mean = 1.9 (SD = 1.1) to 1.1 (0.6)<br>bioavailable Testosterone: Mean = 0.9 (SD = 0.5) to 0.3 (0.2)<br>Free Testosterone: Mean = 0 (SD = 0) to 0 (0)<br>Free Androgen Index: Mean = 7 (SD = 4.1) to 1.8 (1.3)<br>DHEAS: Mean = 3.5 (SD = 2.2) to 2.8 (1.6)<br><br>Postmenopausal women:<br>Change BMI: -33.71 (Mean = 43.9 (SD = 4.2) to 29.1 (3.9))<br>Change waist circumference: -25.02 (Mean = 120.3 (SD = 12.7) to 90.2 (9.8))<br>Change hip circumference: -22.14 (Mean = 127.4 (SD = 13.2) to 99.2 (11.3))<br><br>SHBG: Mean = 36.2 (SD = 21.8) to 96.8 (55.8)<br>Testosterone: Mean = 1.3 (SD = 0.5) to 1 (0.3)<br>bioavailable Testosterone: Mean = 0.6 (SD = 0.3) to 0.2 (0.2)<br>Free Testosterone: Mean = 0 (SD = 0) to 0 (0)<br>Free Androgen Index: Mean = 5 (SD = 3.3) to 1.3 (0.9)<br>DHEAS: Mean = 2.8 (SD = 1.7) to 2.6 (2.2) |
| Sarwer 2018<br>(68)<br>USA<br>Non-randomized intervention<br>Follow-up: 48 months | Sample size n= 106<br><br>Obese women who met to undergo bariatric surgery, BMI: 36.4–66.5;<br>Age: 25-60<br><br>Not reported | Exposures<br>Weight (kg)<br><br>Outcomes<br>Testosterone (ng/dl, not reported)<br>SHBG (nmol/l, not reported)<br>DHEAS (µg/dl, not reported) | Baseline to year 3:<br>Change weight: -32.3 % (95% CI = 30.4% - 34.3%), Baseline (Mean) = 124.1 (IQR = 113.2 to 135.5)<br>Testosterone: Mean = 47.6 (SD = 33.5) to 17.8 (11.7)<br>SHBG: Mean = 42.8 (SD = 37.7) to 87.6 (32)<br>DHEAS: Mean = 119.7 (SD = 85.9) to 79.8 (42.9)<br><br>Baseline to year 4<br>Change weight: -30.6% (95% CI = 28.5% - 32.8%), Baseline (Mean) = 124.1 (IQR = 113.2 to 135.5)<br>Testosterone: Mean = 47.6 (SD = 33.5) to 22.1 (31.4)<br>SHBG: Mean = 42.8 (SD = 37.7) to 87.4 (34.4)<br>DHEAS: Mean = 119.7 (SD = 85.9) to 74.6 (38.8) |

| | | | |
|---|---|---|---|
| Beiglböck 2020<br>(69)<br>Austria<br>Non-randomized intervention<br>Follow-up: 19.3 ± 11.5 months | Sample size n= 81<br><br>Healthy women, BMI: 44.4 (SD=7.1);<br>Age: 40 (SD=11)<br><br>Not reported, seems like both pre + post Mix | Exposures<br>BMI (kg/m^2)<br><br>Outcomes<br>Testosterone (ng/ml, not reported)<br>SHBG (nmol/l, not reported)<br>Bioavailable Testosterone (ng/ml, not reported)<br>DHEAS (µg/ml, not reported)<br>Androstendione (ng/ml, not reported) | Change BMI: -33.78 (Mean = 44.4 (SD = 7.1) to 29.4 (5))<br><br>Testosterone: Mean = 0.2 (SD = 0.1) to 0.2 (0.1)<br>bioavailable Testosterone: Mean = 0.1 (SD = 0.1) to 0.4 (0.4)<br>DHEAS: Mean = 1.4 (SD = 0.7) to 1.2 (0.6)<br>Androstenedione: Mean = 1.2 (SD = 0.7) to 1.1 (0.6)<br>SHBG: Mean = 48.9 (SD = 39.1) to 103.5 (51.1) |
| Linkov 2017<br>(70)<br>USA<br>Non-randomized intervention<br>Follow-up: 6 months | Sample size n= 107<br><br>wome, BMI>=35;<br>Age: mean=43.9 (SD=11.7)<br><br>PRE + POST Mix | Exposures<br>Weight(kg), BMI (kg/m^2),<br>Waist_Circum(cm),<br>WHR<br><br>Outcomes<br>SHBG (not reported, not reported) | linear regression between change in SHBG and change in BMI=-0.041 (p=0.01)<br><br>Weight: Mean = 123.9 (SD= 19.7) to 93.7 (NR)<br>BMI: Mean= 45.5 (6.2) to 34.5 (NR)<br>SHBG: Mean = 10.7 (0.13) to 11.3 (0.03) |
| Menezes 2021<br>(71)<br>Brazil<br>Non-randomized intervention<br>Follow-up: 12 months | Sample size n= 40<br><br>women, BMI: mean=39.2;<br>Age: mean=34.1 (SD=7.2); eligible for bariatric surgery<br><br>Not reported | Exposures<br>Weight(kg), BMI (kg/m^2)<br><br>Outcomes<br>Testosterone (ng/dl, chemiluminescence),<br>Free Testosterone (ng/dl, calculated)<br>Androstendione (ng/dl, chemiluminescence)<br>DHEAS (ng/dl, RIA) | Weight: Mean = 103.8 (NR) to 71 (NR)<br>BMI: Mean= 39.2 (NR) to 27.1 (NR), Testosterone: Mean= 32.3 (17.3) to 28.2 (15), Free Testosterone: Mean= 0.49 (0.3) to 0.33 (0.2), Androstendione: Mean= 2 (2.8) to 1.3 (1.3), DHEAS: Mean= 3.4 (1.4) to 4.2 (2.1) |
| Soykan 2025<br>(72)<br>Turkey<br>Non-randomized intervention<br>Follow-up: 6 months | Sample size n= 67<br><br>wome, BMI>=35;<br>Age=18-49; scheduled to robotic bariatric surgery<br><br>Pre-menopause | Exposures<br>Weight(kg), BMI (kg/m^2)<br><br>Outcomes<br>Testosterone (not reported)<br>SHBG (not reported)<br>DHEAS (not reported)<br>Androstendione ( not reported) | Weight: Mean = 112.34 (19.32) to 71.66 (11.4), BMI: Mean= 42.25 (7.74) to 26.97 (4.67), Androstendione: Mean= 2.41 (1.29) to 2.06 (0.97), Testosterone: Mean= 0.58 (0.3) to 0.45 (0.26), SHBG: Mean= 35.54 (31.47) to 46.32 (33.8), DHEAS: Mean= 229.72 (133.45) to 249.43 (124.07) |
| Lifestyle interventions | | | |

| | | | |
|---|---|---|---|
| Smith 2011<br>(73)<br>USA<br>Randomized clinical trial<br>Follow-up: 4 ± 2 weeks months | Sample size n= Exercisers: 212 (FU=165);<br>controls: 179 (FU=153)<br><br>Premenopausal (follicular phase)<br><br>BMI=18–40 kg/m2;<br><br>Age: exercisers=25.4 (SD=3.4), controls=25.2 (SD=3.5)<br><br>Pre-menopause (follicular) | Exposures<br>BMI (kg/m^2), Weight (kg), Fat mass (kg, DEXA), % Body fat (%, DEXA)<br><br>Outcomes<br>Estradiol (total, free, bio)and testosterone (total, free, bio), pg/mL<br>Estrone sulfate and Progesterone, ng/mL<br>SHBG, nmol/L | Exercisers changes baseline to 4 months<br>Weight and BMI NS change<br>Fat mass: means (SE), mean change=24.4 (0.5) to 24.1 (0.5), -0.57 (1.2%), S<br>Body fat%=36.9 (0.4) to 35.9 (0.4), -0.95 (2.7%), S<br><br>Age- and BMI-adjusted geometric means (95%CI), mean change :<br>Estrone sulfate: 2.0 (1.8 to 2.2) to 2.0 (1.9 to 2.2), -0.04, NS<br>Estradiol: 56 (50-63) to 60 (56-64), 2.3, NS<br>Bioavailable estradiol: 39 (36-41) to 40 (37-43), 1.4, NS<br>Free estradiol: 1.3 (1.2-1.4) to 1.4 (1.3-1.4), 0.05, NS<br>Testosterone: 451 (423-482) to 446 (419-475), -8.8, NS<br>Bioavailable testosterone: 204 (187-222) to 204 (188-222), -2.7, NS<br>Free testosterone: 8.8 (7.9-9.7) to 8.7 (8.0-9.5), -0.13, NS<br>Progesterone: 12 (10-14) to 10 (8-11), -2.2, S<br>SHBG: 27 (25-30) to 26 (24-29), -1.3, NS |
| Mingrone 2002<br>(1)<br>Italy<br>RCT<br>Follow-up: 12 months | Sample size n= Surgical intervention: 21; Diet intervention: 31<br><br>Obese women, diet intervention, BMI: 48.4 (SD = 8.9), surgical intervention, BMI: 48.3 (SD = 6.3); Age: 30-45<br><br>Pre-menopause<br>(Follicular) | Exposures<br>Fat mass (kg, DEXA), Weight (kg), BMI (kg/m^2)<br><br>Outcomes<br>SHBG (nmol/l, Immunoassay) | Dietary intervention<br>Change fat mass: -8.688 (Mean = 63.3 (SD = 16.2) to 57.8 (SD = 16.5)<br>Change BMI: -9.5 (Mean = 48.4 (SD = 8.9) to 43.8 (SD = 7.7))<br>SHBG: Mean = 17.3 (SD = 8) to 21.6 (SD = 9.4) |
| Gonzalo-Encabo 2021<br>(8)<br>Canada<br>RCT<br>Follow-up: 12,24 months | Sample size n= 214<br><br>Women randomised in 2 exercises groups<br>Age 59.1 (56.5 to 63.0)<br>BMI between 22–40 kg/m2<br><br>Post-menopause | Exposures<br>Body weight (kg, balance beam scale)<br>Fat mass (kg, DXA)<br><br>Outcomes<br><br>Estradiol, Free estradiol, and Estrone (pg/mL, radioimmunoassay)<br>SHBG (nmol/L, solid-phase, two-site chemiluminescent immunometric assays on the Immulite analyzer) | median (IQR)<br>Baseline to 12 months (n=214)<br>All weight loss (n=214 including the 36 who regain >5 kg between 12 and 24 months FU)<br>Fat mass 29.1 (23.9–36.1) to 25.8 (21.1–31.5) (-11%)<br>Estradiol 9.45 (7.29–12.62) to 8.6 (6.7–11.4)<br>Estrone 37.91 (30.71–46.27) to 35.1 (28.4–45.0)<br>SHBG 45 (34–60.7) to 49.1 (38–67.3)<br>Free estradiol 0.21 (0.15–0.31) to 0.20 (0.14–0.28)<br><br>12 to 24 months for those who regain >=5 kg (n=36)<br>Body weight 73.4 (66.6–81.9) to 82.5 (74.2–89.1) (+12%)<br>Fat mass 27.3 (22.3–32.1) to 34.1 (26.8–37.7) (+25%)<br>Estradiol 8.58 (7.1–11.96) to 9.85 (7.59–14.3)<br>Estrone 36.3 (28.4–45.01) to 35 (28.6–43.4)<br>SHBG 49.2 (39.3–76.5) to 37.4 (31.8–55.9)<br>Free estradiol 0.19 (0.15–0.30) to 0.27 (0.20–0.36)<br><br>12 to 24 months for those who regain >=2.6 until 4.9 kg (n=51)<br>Body weight 74.2 (67.1–82.1) to 69.1 (63.2–76.9)(-7%)<br>Fat mass 28.5 (23.9–34.7) to 24.9 (20.4–30.2)(-12%)<br>Estradiol 8.88 (7.08–11.59) to 8.8 (6.7–10.6) |

| | | | |
|---|---|---|---|
| | | | Estrone 35.6 (27.4–45.3) to 34.4 (26.4–44.4)<br>SHBG 46.1 (32.6–61.5) to 54.6 (38.1–67.5)<br>Free estradiol 0.21 (0.15–0.29) to 0.19 (0.14–0.25)<br><br>A TER <1.0 indicated lower biomarker concentrations in the weight regain category versus the reference group during follow-up; a ratio >1.0 indicated higher biomarker concentrations in the weight regain categories versus the<br>reference group during follow-up; and a ratio equal to 1.0 indicated no change in biomarker concentrations between weight categories.<br>ref group those who continue to loss weight or maintain it.<br><br>Weight regain during follow-up was significantly associated with increases in estradiol (TER = 1.03; 95% CI, 1.01–1.04; P < 0.001), estrone (TER = 1.02; 95% CI, 1.01–1.03; P < 0.001), free estradiol (TER = 1.04; 95% CI, 1.02–1.05; P < 0.001), and decreases in SHBG (TER = 0.98; 95% CI, 0.97–0.99; P < 0.001) levels (Fig. 2).<br><br>no |
| Elsayed 2022<br>(50)<br>Egypt<br>RCT<br>Follow-up: 3 months | Sample size n= 30+30<br><br>Women with mild cognitive impairment (MCI) to dementia.<br>- Experimental diet mean (SD) n=30, age=65.4 (2.8), BMI=35.8 (2.9)<br>- Control n=30, age=65.1 (3.2), BMI=35.8 (2.7)<br><br>Post-menopause | Exposures<br>BMI (kg/m^2)<br><br>Outcomes<br>Estradiol (pg/ml, chemiluminescence)<br>Total testosterone (ng/ml, chemiluminescence)<br>Free testosterone (pg/ml, chemiluminescence)<br>SHBG (nmol/ml, chemiluminescence) | Experimental group n=30, mean (SD)<br>BMI 35.77 (2.87) to 31.67 (2.85), t-test: p< 0.01<br>Estradiol 27.69 (2.73) to 22.29 (2.96), p< 0.01<br>Total testosterone 0.40 (0.09) to 0.30 (0.08), p< 0.01<br>Free testosterone 1.93 (0.63) to 1.34 (0.50), p< 0.01<br>SHBG 28.13 (5.45) to 34.54 (6.71), p< 0.01<br><br>Control group n=30<br>BMI 35.83 (2.74) to 33.49 (2.80), p< 0.05<br>Estradiol 27.70 (2.88) to 25.23 (3.00), p< 0.05<br>Total testosterone 0.40 (0.08) to 0.37 (0.08), p< 0.05<br>Free testosterone 1.79 (0.68) to 1.65 (0.62), p< 0.05<br>SHBG 27.81 (5.55) to 30.25 (5.88), p< 0.05<br><br>no |
| Goodfriend 1999<br>(2)<br>USA<br>Non-randomized intervention<br>Follow-up: 4 months | Sample size n= 21<br><br>Women with BMI>=30 kg/m2;<br>Age: 73 (SD=1)<br><br>Pre-menopause<br>(Unknown) | Exposures<br>Fat mass (kg)<br><br>Outcomes<br><br>DHEAS (ng/dl, RIA) | Mean weight loss was 12.2±1.0 kg (no basline data), average BMI following weight loss was 29.5 kg/m2 (not mentioned if for women, men or both), VAT decreased (no value mentioned)<br><br>DHEAS: Mean = 139 (SEM = 19) to 128 (16)<br><br>not reported in full text |
| Kovacikova 2008<br>(47)<br>France<br>Non-randomized intervention<br>Follow-up: 3 months | Sample size n= 32<br><br>Overweight and obese women, BMI: 31.2 (SEM=0.9);<br>Age: 42.1 (SEM=1.9)<br><br>Pre-menopause<br>(Unknown) | Exposures<br>Waist circumference (cm)<br>WHR<br>Fat mass (kg, multifrequency bioimpedance)<br>BMI (kg/m^2) | Change Waist circumference: -8.741 (Mean = 96.1 (SD = 2.1) to 87.7 (1.8))<br>Change WHR: 0 (Mean = 0.8 (SD = 0) to 0.8(0))<br>Change Fat mass: Mean = 35.6 (SD = 1.8) to 30.4(1.9))<br>Change BMI: -8.013 (Mean = 31.2 (SD = 0.9) to 28.7(0.9))<br><br>Total Testosterone: Mean = 1.8 (SD = 0.2) to 1.6 (0.2)<br>Free Testosterone: Mean = 5.3 (SD = 0.5) to 4.2 (0.5)<br>SHBG: Mean = 60.1 (SD = 6.3) to 75.5 (7.6) |

| | | | |
|---|---|---|---|
| | | Outcomes<br>Total testosterone (nmol L-1, calculated)<br>Free Testosterone (nmol L-1, RIA)<br>SHBG (nmol L-1, RIA) | |
| Harvie 2011<br>(35)<br>UK<br>RCT<br>Follow-up: 6 months | Sample size n= Intermittent energy restriction: 53,<br>Continous energy restriction: 54<br><br>Women with adult weight gain > 10kg since the age of 20, BMI=24-40 kg/m2;<br>Age: 40.1 (SD=4.1) Intermittent energy restriction, 40.0 (SD=3.9) Continous energy restriction<br><br>Pre-menopause<br>(Unknown) | Exposures<br>% Body fat (%, impedance)<br>Weight (kg)<br>Fat mass (kg)<br><br>Outcomes<br>SHBG (nmol/l, non-competitive IRMA)<br>DHEAS (µmol/l, liquid chromatography and tandem mass spectrometry)<br>Androstendione (µmol/l, liquid chromatography and tandem mass spectrometry)<br>Testosterone (nmol/l, liquid chromatography and tandem mass spectrometry)<br>Free Androgen index (calculated by the equation: serum Testosterone/(serum SHBG x 100)) | Intermittent energy restriction (IER):<br>Baseline to 3 Month:<br>Change % Body fat: -4.938 (Mean = 40.5 (SD = 5.6) to 38.5 (6.9))<br>Change weight: -5.031 (Mean = 81.5 (SD = NR) to 77.4 (NR))<br>Change Fat mass: -8.929 (Mean = 33.6 (SD = 10.2) to 30.6 (10.8))<br>SHBG: geomean = 43.2 (SD = 20.1) to 48.6 (23.3)<br>Androstenedione: geomean = 2.7 (SD = 1.1) to 2.8 (1)<br>DHEAS: geomean = 3.2 (SD = 1.7) to 3.3 (1.5)<br>Testosterone: geomean = 0.8 (SD = 0.4) to 0.8 (0.3)<br>Free Androgen Index: geomean = 1.7 (SD = 1.1) to 1.6 (0.9)<br><br>Basline to 6 Month:<br>Change % Body fat: -7.901 (Mean = 40.5 (SD = 5.6) to 37.3 (7.2))<br>Change weight: -6.994 (Mean = 81.5 (SD = NR) to 75.8 (NR))<br>Change Fat mass: -13.39 (Mean = 33.6 (SD = 10.2) to 29.1 (10.4))<br>SHBG: geomean = 43.2 (SD = 20.1) to 49.2 (21.3)<br>Androstenedione: geomean = 2.7 (SD = 1.1) to 2.9 (1)<br>DHEAS: geomean = 3.2 (SD = 1.7) to 3.3 (1.7)<br>Testosterone: geomean = 0.8 (SD = 0.4) to 0.8 (0.3)<br>Free Androgen Index: geomean = 1.7 (SD = 1.1) to 1.6 (0.8)<br><br>Continous energy restriction (CER):<br>Baseline to 3 Month:<br>Change % Body fat: -3.704 (Mean = 40.5 (SD = 6.8) to 39 (7.4))<br>Change weight: -3.555 (Mean = 84.4 (SD = NR) to 81.4 (NR))<br>Change Fat mass: -6.799 (Mean = 35.3 (SD = 12.8) to 32.9 (13.1))<br>SHBG: geomean = 42 (SD = 17.6) to 44.3 (16.3)<br>Androstenedione: geomean = 3.1 (SD = 1.1) to 3 (1.2)<br>DHEAS: geomean = 3.4 (SD = 1.5) to 3.2 (1.4)<br>Testosterone: geomean = 0.9 (SD = 0.4) to 0.8 (0.4)<br>Free Androgen Index: geomean = 2 (SD = 1.1) to 1.8 (0.7)<br><br>Baseline to 6 Month:<br>Change % Body fat: -6.173 (Mean = 40.5 (SD = 6.8) to 38 (7.9))<br>Change weight: -5.332 (Mean = 84.4 (SD = NR) to 79.9 (NR))<br>Change Fat mass: -10.2 (Mean = 35.3 (SD = 12.8) to 31.7 (13.3))<br>SHBG: geomean = 42 (SD = 17.6) to 44.6 (18.4)<br>Androstenedione: geomean = 3.1 (SD = 1.1) to 3.1 (1.05)<br>DHEAS: geomean = 3.4 (SD = 1.5) to 3.2 (1.4)<br>Testosterone: geomean = 0.9 (SD = 0.4) to 0.8 (0.4)<br>Free Androgen Index: geomean = 2 (SD = 1.1) to 1.8 (1.1) |

| | | | |
|---|---|---|---|
| Swora-Cwynar 2016<br>(48)<br>Poland<br>RCT<br>Follow-up: 3 months | Sample size n= 39<br><br>Obese women, BMI ≥ 30;<br>Age: 31.2 (SD=8.3)<br><br>Pre-menopause<br>(Unknown) | Exposures<br>Weight (kg)<br>Fat mass (kg, BIA)<br>% Body fat (BIA)<br>BMI ( kg/m^2)<br><br>Outcomes<br>Testosterone (nmol/l, electrochemiluminescent immunoassay)<br>DHEAS (µg/dl, electrochemiluminescent immunoassay)<br>Estradiol (pg/ml, electrochemiluminescent immunoassay) | Change Weight: -4.573 (Mean = 100.6 (SD = 16.4) to 96 (14.3))<br>Change Fat mass: -10.17 (Mean = 47.2 (SD = 10.8) to 42.4 (12))<br>Change % Body fat: -3.863 (Mean = 46.6 (SD = 5.6) to 44.8 (5.3))<br>Change BMI: -4.432 (Mean = 36.1 (SD = 5.6) to 34.5 (4.9))<br>Testosterone: Mean = 1.5 (SD = 0.5) to 1.2(0.6)<br>DHEAS: Mean = 296.3 (SD = 123.4) to 249.6 (133.8)<br><br>Wilcoxon, Spearman correlation:<br>Change fatmass and Testosterone =0.64, Change fatmass and DHEAS =0.21, Change %Body fat and testosterone= 0.21, Change %Body fat and DHEAS= 0.35,<br>Change BMI and Testosterone = 0.55, Change BMI and DHEAS = -0.14 |
| Leenen 1994<br>(3)<br>Netherlands<br>Non-randomized intervention<br>Follow-up: 6 months | Sample size n= 33<br><br>Healthy women, BMI: 31.3 (SD=2.2);<br>Age: 39 (SD=5)<br><br>Pre-menopause<br>(Follicular) | Exposures<br>WHR (cm2, MRI)<br>VAT (cm2, MRI)<br>Weight (kg)<br>Abdominal fat area (cm2, MRI)<br>Fat mass (kg)<br>BMI (kg/m^2)<br><br>Outcomes<br>Estrone (pmol/l, RIA)<br>Estradiol (pmol/l, RIA)<br>Free Estradiol (pmol/l, calculated from SHBG and Total Estradiol)<br>Testosterone (nmol/l, RIA)<br>SHBG (nmol/l, IRMA)<br>Free Testosterone (nmol/l, calculated from Total Testosterone and SHBG)<br>DHEAS (µmol/l, RIA)<br>Androstenedion (nmol/l,RIA) | Change baseline - 6 months FU<br>WHR: -4.598 (Mean = 0.87 (SD = 0.07) to 0.83 (NR))p<0.0001<br>VAT: -32.04 (Mean = 103 (SD = 35) to 70 (NR)), p<0.0001<br>Weight: -14.27 (Mean = 86.9 (SD = 7.6) to 74.5 (NR), p<0.0001<br>SAT: -29.62 (Mean = 395 (SD = 111) to 278 (NR), p<0.0001<br>Fat mass: -29.79 (Mean = 38.6 (SD = 6.3) to 27.1 (NR), p<0.0001<br>BMI: -14.38 (Mean = 31.3 (SD = 2.2) to 26.8 (NR)), p<0.0001<br><br>Baseline to FU (calculated using changes)<br>Estrone: Mean = 216 (SD = 66) to 215 (NR), p=NS<br>Estradiol: Mean = 254 (SD = 143) to 203 (NR), p<0.05<br>Free Estradiol: Mean = 254 (SD = 143) to 203 (NR), p<0.01<br>Testosterone: Mean = 1.2 (SD = 0.3) to 1 (NR), p<0.0001<br>Free Testosterone: Mean = 0.03 (SD = 0.01) to 0.02 (NR), p<0.0001<br>SHBG: Mean = 30 (SD = 14) to 37 (NR), p<0.01<br>DHEAS Mean=5.3 (SD=2.4) to 3.4 (NR), p<0.01<br>Androstenedione Mean=5.2 (SD=1.3) to 3.8 (NR), p<0.0001 |
| Kraemer 2003<br>(55)<br>USA<br>RCT<br>Follow-up: 9 months | Sample size n= Non periodized resistance training: 10<br>Periodized resistance training: 9<br><br>Women tennis players, Weight: 60.8 (SD=7.8) Non periodized | Exposures<br>% Body fat (%, determined using body-density and Siri equation)<br>Body mass (kg) | Non-periodized resistence training<br>Baseline to 4 Month:<br>Change % Body fat: -10.97 (Mean = 23.7 (SD = 4.9) to 21.1 (2.4)), S<br>SHBG: Mean = 60 (SD = 39) to 61 (39), NS<br><br>Baseline to 6 Month: |

| | | | |
|---|---|---|---|
| | resistance training, 60.5 (SD=7.7) Periodized resistance training;<br>Age: 18.6 (SD=1.3) Non periodized resistance training, 19.2 (SD=1.1) Periodized resistance training<br><br>Pre-menopause<br>(Follicular) | Outcomes<br>SHBG (nmol/l, double antibody liquid phase RIA) | Change % Body fat: -10.127 (Mean = 23.7 (SD = 4.9) to 21.3 (3.4)), S<br>SHBG: Mean = 60 (SD = 39) to 62 (39), NS<br><br>Baseline to 9 Month:<br>Change % Body fat: -8.86 (Mean = 23.7 (SD = 4.9) to 21.6 (2.9)), S<br>SHBG: Mean = 60 (SD = 39) to 65 (34), NS<br><br>Periodized resistence training<br>Baseline to 4 Month:<br>Change % Body fat: -12.664 (Mean = 22.9 (SD = 3.9) to 20 (3.4)), S<br>SHBG: Mean = 75.8 (SD = 47.3) to 81.5 (47.8), NS<br><br>Baseline to 6 Month:<br>Change % Body fat: -11.79 (Mean = 22.9 (SD = 3.9) to 20.2 (3.5)), S<br>SHBG: Mean = 75.8 (SD = 47.3) to 80.5 (39.4), NS<br><br>Baseline to 9 Month:<br>Change % Body fat: -16.594 (Mean = 22.9 (SD = 3.9) to 19.1 (3.6)), S<br>SHBG: Mean = 75.8 (SD = 47.3) to 76 (36.5), NS |
| Boyden 1983<br>(49)<br>USA<br>Non-randomized intervention<br>Follow-up: average 13.5 months | Sample size n= 19<br><br>Healthy, menstruating regularly, Weight: 56.8 (SEM=1.2;<br>Age: 29.3 (range=24 to 37)<br><br>Pre-menopause<br>(Follicular) | Exposures<br>Fat mass (kg)<br>% relative fat mass (%, body fat percentage estimated based on the ratio of height and waist measurements)<br><br>Outcomes<br>Estrone (pg/ml, RIA)<br>Estradiol (pg/ml, RIA)<br>Testosterone (pg/ml, RIA)<br>Free testosterone ( %,estimated by centrifugal ultrafiltration dialysis with the use of the MPS-1 micropartition system) | Baseline to 30 Miles per week:<br>Change Fat mass: -9.091 (Mean = 14.3 (SD = 3.5) to 13 (3.1))<br>Change % relative fat: -8.434 (Mean = 24.9 (SD = 4.8) to 22.8 (4.4))<br>Estrone: Mean = 52.6 (SE = 13.9) to 48.8 (2.8)<br>Estradiol: Mean = 70.6 (SE = 60.6) to 53.6 (8.7)<br>Testosterone: Mean = 242 (SE = 127.3) to 206.3 (20.8)<br>Free Testosterone: Mean = 1.1 (SE = 0.4) to 1.2 (0.1)<br><br>Baseline to 50 Miles per week:<br>Change fat mass: -11.89 (Mean = 14.3 (SD = 3.5) to 12.6 (2.6))<br>Change % relative fat: -11.65 (Mean = 24.9 (SD = 4.8) to 22 (3.1))<br>Estrone: Mean = 52.6 (SE = 13.9) to 44.7 (3.4)<br>Estradiol: Mean = 70.6 (SE = 60.6) to 33.6 (4.8)<br>Testosterone: Mean = 242 (SE = 127.3) to 222.5 (32.6)<br>Free Testosterone: Mean = 1.1 (SE = 0.4) to 1.2 (0.1)<br><br>Pearson correlation of Fatmass and Estrone= 0,51, p <0,03 (significant) (statused: analysis of variance for autocorrelated repeated measures, paired t-test, and linear regression) |
| Aubuchon 2016<br>(9)<br>USA<br>RCT<br>Follow-up: 4 - 6 months | Sample size n= 30<br><br>Women, non smokers, BMI: 32.9 (SD=4.2);<br>Age: 40 (SD=5.9)<br><br>Pre-menopause<br>(Unknown but same phase) | Exposures<br>Weight (kg)<br>logVAT(cm^2, computed tomography)<br>% Body fat (%, DEXA)<br>BMI (kg/m^2)<br><br>Outcomes | Weight loss - 4 to 6 mo interevntion<br>Change weight: -9.551 (Mean = 89.0 (SD = 13.9) to 80.5 (12.6)), S<br>Change logVAT: -5.439 (Mean = 4.8 (SD = 0.4) to 4.5 (0.6)), S<br>Change % Body fat: -11.92 (Mean = 38.6 (SD = 4.5) to 34 (4.5)), S<br>Change BMI: -9.422 (Mean = 32.9 (SD = 4.2) to 29.8 (3.8)), S<br><br>logTestosterone: Mean = 3.2 (SD = 0.7) to 3.1 (0.7), NS<br>logSHBG: Mean = 3.9 (SD = 0.8) to 4.1 (0.7), S |

| | | | |
|---|---|---|---|
| | | logTestosterone (ng/dl, Immulite 1000 solid phase chemiluminescent immunoassay)<br>logSHBG (nmol/l, Immulite 1000 solid phase chemiluminescent immunoassay)<br>logFree Androgen Index (calculated from Testosterone and SHBG)<br>Testosterone (ng/dl, Immulite 1000 solid phase chemiluminescent immunoassay)<br>SHBG (nmol/l, Immulite 1000 solid phase chemiluminescent immunoassay) | logFree Androgen Index: Mean = 0.5 (SD = 1) to 0.3 (1), S<br>Testosterone: Mean = 30.5 (SD = 19.3) to 28.3 (16.4), NS<br>SHBG: Mean = 67 (SD = 59.9) to 76.5 (60.8), NS<br><br>After intervention, 4 to 6 mo with otr without PA, Weight regain<br>Change weight: (Mean = 80.5 (12.6) to 85.1 (13.4)), S<br>Change logVAT: -5.439 (Mean = 4.5 (0.6) to 4.6 (0.5)), S<br>Change % Body fat: (Mean = 34 (4.5) to 36.6 (4.8)), S<br>Change BMI: (Mean = 29.8 (3.8) to 31.5 (4.1)), S<br><br>logTestosterone: Mean = 3.1 (0.7) to 3.2 (0.7), NS<br>logSHBG: Mean = 4.1 (0.7) to 3.9 (0.7), S<br>logFree Androgen Index: Mean = 0.3 (1) to 0.5 (1.2), S<br>Testosterone: Mean = 28.3 (16.4) to 31.2 (22.5), NS<br>SHBG: Mean = 76.5 (60.8) to 68.5 (71.8), NS |
| Van Rossum 2000<br>(56)<br>USA<br>Non-randomized intervention<br>Follow-up: 6 months | Sample size n= 54<br><br>Obese women, BMI: 32.0 (SD=4.5); Age: 60 (SD=6)<br><br>Post-menopause | Exposures<br>Fat mass (unit =kg, DEXA)<br>% Body fat (%, DEXA)<br>BMI (kg/m^2)<br>IAF (cm2, CT-scan)<br>SAT (cm2, CT-scan)<br><br>Outcomes<br>SHBG (nmol/l, IRMA) | Change Fat mass: -13.333 (Mean = 39 (SD = 9.1) to 33.8 (10.1))<br>Change % Body fat: -8.0679 (Mean = 47.1 (SD = 5.4) to 43.3 (6.6))<br>Change BMI: -6.875 (Mean = 32 (SD = 4.5) to 29.8 (4.7))<br>Change intra-abdominal fat: Mean = 154.7 (SD = 51.2) to 129.8 (47.2))<br>Change SAT: -11.198 (Mean = 449.2 (SD = 124) to 398.9 (121.2))<br>SHBG: Mean = 122 (SD = 75.3) to 136.1 (99.6) |
| Ricci 2001<br>(60)<br>USA<br>RCT<br>Follow-up: 6 months | Sample size n= 14<br><br>Obese women, BMI: 33.0 (SD=3.8); Age: 55.0 (SD=8.0)<br><br>Post-menopause | Exposures<br>Fat mass (kg, DEXA)<br><br>Outcomes<br>SHBG (nmol/l, 2-site IRMA)<br>Estrone (pmol/L, RIA) | Weight loss group:<br>Change Fat mass: -18.7 (Mean = 40.3 (SD = 7.2) to 32.8 (4.6))<br>SHBG: Mean = 73.8 (SD = 35.2) to 101.1 (15.8)<br>Estrone: Mean = 94.4 (SD = 52.5) to 100 (30.7)<br><br>Association between loss of fat mass (Percent) and the percentage change in SHBG: R^2 = 0.34 (p<0.05);<br>Association between loss of fat mass (Percent) and the percentage change in estrone: R^2=0.5 (p<0.01);<br><br>For measurements before and after weight loss, we performed a two-way ANOVA. If a main effect or interaction was significant linear contrast analysis (Scheffeâ€™s test) was performed. Values were compared before and after treatment within a group with Student paired t tests. |
| Bhargava 2006<br>(58) | Sample size n= 615 | Exposures<br>Hip circumference (m) | Controls<br>Weight: 75.92 (12.6) to 75.53 (12.62), S |

| | | | |
|---|---|---|---|
| USA<br>RCT<br>Follow-up: 12 months | Healthy, BMI: 28.77 (SD=4.43);<br>Age: 60.63 (SD=6.63)<br><br>Post-menopause | Waist circumference (m)<br>BMI (unti = kg/m^2)<br><br>Outcomes<br>SHBG (nmol/l, IRMA) | BMI: 28.90 (4.65) to 28.75 (4.68), S<br>SHBG: 92.57 (71.59) to 102.68 (76.61), S<br><br>Intervention<br>Weight: 75.95 (12.43) to 73.86 (12.6), S<br>BMI: 8.77 (4.43) to 27.98 (4.47), S<br>SHBG: 89.82 (65.19) to 105.99 (76.18),S |
| O'Dea 1979<br>(54)<br>USA<br>Non-randomized intervention<br>Follow-up: 3 - 4 months | Sample size n= 12<br><br>Obese women, Weight 86.2-130.2;<br>Age: 50-63<br><br>Post-menopause | Exposures<br><br>Weight (kg)<br><br>Outcomes<br>Estradiol (pg/ml, RIA)<br>Testosterone (ng/dl, measured as unfractionated 17beta-ol androgens)<br>SHBG (not reported, DHT saturation) | 2.4 to 8.3 weeks<br>Change weight: -8.125 (Mean = 105.2 (SD = 15.6) to 96.7 (15.6))<br>Testosterone: Mean = 38 (SD = 8) to 40 (8)<br>SHBG: Mean = 0.5 (SD = 0.2) to 1.2 (0.5)<br><br>Follow-up 11 to 17 weeks<br>Change weight: -3.174 (Mean = 84.1 (SD = 18) to 81.5 (15.3))<br>Testosterone: Mean = 38 (SD = 8) to 39 (5)<br>SHBG: Mean = 0.5 (SD = 0.2) to 1.4 (0.5) |
| Caballero MJ 1992<br>(59)<br>Spain<br>Non-randomized intervention<br>Follow-up: 4 months | Sample size n= 18<br><br>Women at least one year post-menopausal, Weight: 68.6 (SD=7.5);<br>Age: 54 (SD=7)<br><br>post-menopausal | Exposures<br>Suprailiac skinfold thickness (Harperiden skinfold caliper)<br>Subscapular Skinfold thickness (Harperiden skinfold caliper)<br>% Body fat (%)<br>Triceps skinfold thickness (Harperiden skinfold caliper)<br><br>Outcomes<br>SHBG (nmol/l, endogeneous steroids were removed from plasma with charcoal suspension, then incubation and free steroid was removed using charcoal suspension, SHBG was expressed in terms of a molar concentration of bound 3H-DHT after substraction of non-specific binding) | Change Suprailiac skinfold thickness: -31.148 (Mean = 18.3 (SD = 7.4) to 12.6 (4.1))<br>Change Subscapular Skinfold thickness: -12.054 (Mean = 22.4 (SD = 7.5) to 19.7(6.4))<br>Change Triceps skinfold thickness: -15.421 (Mean = 21.4 (SD = 5.7) to 18.1 (4.2))<br>Change % Body fat: -11.735 (Mean = 19.6 (SD = 3.6) to 17.3 (2.9))<br>SHBG: Mean = 55.3 (SD = 20.9) to 48.3 (21) |
| Kim 2012<br>(57) | Sample size n= 15 | Exposures<br>WC (cm) | %change, mean (SD), diff S |

| | | | |
|---|---|---|---|
| South Korea<br>RCT<br>Follow-up: 4 months | Obese and sedentary women, no menstruation at least 1 year, > 32% body fat, plasma FSH levels > 30 mIU/mL;<br>Age: 54.53 (SD=2.82)<br><br>Post-menopause | % Body fat (%, BIA)<br>BMI (kg/m^2)<br>VAT (cm^2,BIA)<br><br>Outcomes<br>SHBG (µg/ml, IRMA) | Exercise intervention group, decrease body fat<br>Change waist circumference: -1.39%, 86.2 (1.5) to 85 (1.4), S<br>Change % Body fat: -6.9%, 36.0 (3.0) to 33.5 (3.3), S<br>Change BMI: -3.2%, 25 (1.3) to 24.2 (1.2), S<br>Change VAT: -5.12%, 103.5 (10.1) to 98.2 (7.7), S<br>Weight: -3.6%, 60.9 (4.0) to 58.7 (3.8), S<br>SHBG: 6.2%, 43.3 (9.2) to 46 (10)<br><br>Control group, increase body fat<br>Change waist circumference: 1.04%, 85.9 (1.7) to 86.8 (1.7)), S<br>Change % Body fat: 2.6%, 36.6 (1.7) to 37.5 (2.4), S<br>Change BMI: 3.3%, 25.1 (1.5) to 25.9 (1.4), S<br>Change VAT: 5.1%, 100.5 (8.4) to 105.7 (8.6), S<br>Weight: 2.5%, 61.6 (3.9) to 63.1 (4.3), S<br>SHBG: -4.6%, 44.2 (8.1) to 42.2 (8.8), S<br><br>Regression analysis of SHBG and WC: beta= -0,19, t value= 0,586 (NS) (adjusted for TG, HDL, SBP, DBP, Glucose) |
| van Gemert 2017 (61)<br>Netherlands<br>RCT<br>Follow-up: 4 months | Sample size n= 243<br><br>Overweight women, BMI: 25–35;<br>Age: 60 (SD=4.8)<br><br>Post-menopause | Exposures<br>SAAT (cm2, MRI)<br>IAAT (cm2, MRI)<br>Fat mass (kg, DEXA)<br><br>Outcomes<br>Free Estradiol (pg/ml, LC-MS)<br>Free Testosterone (pg/ml, LC-MS)<br>SHBG (nmol/l, RIA) | Baseline to 4 months FU change<br>%Change, mean (SD)<br>SAAT: -11.23 (310 (68) to 275.2 (NR))<br>IAAT: -10.28 (144 (47) to 129.2 (NR))<br>Fat mass: -10.29 (34 (6.2) to 30.5 (NR))<br><br>Free Estradiol: geomean = 0.09 (95% CI = 0.09 to 0.10) to 0.07 (NR)<br>Free Testosterone: geomean = 2.53 (95% CI = 2.38 to 2.69) to 2.18 (NR)<br>SHBG: geomean = 48.6 (95% CI = 45.9 to 51.4) to 55.3 (NR)<br><br>Univariable linear regression models for associaiton between change in fat and change in biomarker, Multivariable with the 3 body composition type in the model (fat mass, SAAT, IAAT):<br>SAAT<br>Free estradiol (St-b= 0.11 , p=0.107), NS<br>Free testosterone (St-b= 0.26 , p=<0.001), NS<br>SHBG (St-b= -0.26, p=<0.001), NS<br><br>IAAT<br>Free estradiol (St-b= 0.07, p=0.291), NS<br>Free testosterone (St-b= 0.18, p= 0.007), NS<br>SHBG (St-b= -0.29, p=<0.001), NS<br><br>Fat mass<br>Free estradiol (St-b= 0.18 , p=0.007), NS<br>Free testosterone (St-b= 0.27, p=<0.001), NS<br>SHBG (St-b= -0.39, p=<0.001), -0.34 (-0.59;-0.10) |

| | | | |
|---|---|---|---|
| Duggan 2019<br>(74)<br>USA<br>RCT<br>Follow-up: 30 months | Sample size n= 31 (group that lost >10% weight)<br><br>Healthy, overweight (BMI > 25 kg/m2) and sedentary women; Age: 50 - 75<br><br>Post-menopause | Exposures<br>Weight change (%) data was only extracted for group with weigth loss >=10%<br>Weight loss category ≤0%, 0-5%, 5-10%, ≥10%<br><br>Outcomes<br>Free Estradiol (pg/ml, calculated using measured values for Total Estradiol, SHBG, and an assumed constant for albumin)<br>SHBG (nmol/l, quantified via chemiluminescent immunometric assay using the Immulite Analyzer)<br>Estradiol (pg/ml, RIA after organic solvent extraction and Celite column partition chromatography)<br>Testosterone (ng/dl, RIA after organic solvent extraction and Celite column partition chromatography)<br>Estrone (pg/ml, RIA after organic solvent extraction and Celite column partition chromatography)<br>Free Testosterone (pg/ml, calculated using measured values for Total Testosterone, SHBG, and an assumed constant for albumin) | Weight loss ≥10% (geomean (SD)), ptrend across category loss weight.<br>Free Estradiol: 0.3 (0.1) to 0.1 (0),change=-0.18, p=0.005, ptrend=0.02<br>SHBG: 35.3 (15.1) to 73.5 (25.4), 38.1,<0.001, <0.001<br>Estradiol: 10.6 (5) to 5.8 (2.7), -4.8, 0.15, 0.28<br>Testosterone: 22.8 (11.1) to 25.5 (13.8), 2.7,0.95, 0.84<br>Estrone: 30.5 (13.9) to 33.3 (12.2), 2.8, 0.79, 0.72<br>Free Testosteorne: 4.9 (2.6) to 4 (2.2), -0.9, 0.03, 0.04 |
| Svendsen 1995<br>(52)<br>Denmark<br>RCT<br>Follow-up: 3 months | Sample size n= n-estimated:<br>gynoid: lowest tertiles of WHR, 98/3 -> n~32<br>android: highest tertiles of WHR, 98/3 -> n~32 | Exposures<br>Weight (kg)<br>VAT (cm^2, predicted with an equation)<br>Fat mass (kg, DEXA) | Gynoid group<br>Change VAT: -45.33 (Mean = 43.9 (SD = 46.5) to 24 (NR))<br>Change Weight: -12.31 (Mean = 78 (SD = 1.5) to 68.4 (NR))<br>Change Fat mass: -26.57 (Mean = 33.5 (SD = 1.2) to 24.6 (NR))<br>SHBG: Mean = 53.9 (SEM = 4.4) to 84.5 (SEM change = 4.9)<br>Testosterone: Mean = 1.36 (SEM = 0.09) to 1.23 (SEM change = 0.07) |

| | | | |
|---|---|---|---|
| | healthy, overweight, postmenopausal women, BMI: 25-42;<br>Age: 53.8 (range=49-58) -> for n=98<br><br>Post-menopause | Outcomes<br>SHBG (nmol/l, IRMA)<br>Testosterone (nmol/l, RIA)<br>Androstendione (nmol/l, RIA)<br>Estradiol (pmol/l, RIA) | Androstenedione: Mean = 2.5 (SEM = 0.2) to 2.3 (SEM change = 0.1)<br><br>Android group<br>Change VAT: -23.08 (Mean = 134.3 (SD = 3.2) to 103.3 (NR))<br>Change Weight: -12.45 (Mean = 80.3 (SD = 1.8) to 70.3 (NR))<br>Change Fat mass: -25.77 (Mean = 32.6 (SD = 1.2) to 24.2 (NR))<br>SHBG: Mean = 51.4 (SEM = 3.6) to 83.7 (SEM change = 4.1)<br>Testosterone: Mean = 1.3 (SEM = 0.08) to 1.3 (SEM change = 0.07)<br>Androstenedione: Mean = 2.6 (SEM = 0.2) to 2.5 (SEM change = 0.1)<br><br>Statused: Student's unpaired t-test (diff gynoid/Android); The changes (increase) in SHBG levels were negatively correlated with changes (reductions) in weight, FTM (r = -0.4 to -0.5, P < 0.01), the WC, the trunk, abdominal and legs FTM, VAT, and the WTH (r= -0.3 to -0.4, P < 0.01). The initial values or changes in serum total-testosterone, androstendione, or estradiol were not significantly correlated to initial values or changes in weight, FTM, or any of the fat distribution parameters studied (P > 0.01). |
| Stolzenberg-Solomon 2012<br>(7)<br>USA<br>RCT<br>Follow-up: 6 months | Sample size n= 278<br><br>Overweight and obese postmenopausal, BMI: 33.4 (SD=4.9);<br>Age: 59.3 (range= 46 - 78)<br><br>Post-menopause | Exposures<br>Weight (kg)<br><br>Outcomes<br>Androstendione (pg/ml, RIA)<br>Estrone (pg/ml, RIA)<br>Estradiol (pg/ml, RIA)<br>Testosterone (ng/dl, RIA)<br>DHEAS (µg/dl, RIA)<br>SHBG (nmol/l, RIA)<br>Free Estradiol (pg/ml, calculated from measured Estradiol and SHBG with albumin assumed to be a constant (40 g/l) using the method by Sodergard and colleagues according to the law of mass action)<br>Free Testosterone (pg/ml, calculated from measured Testosterone and SHBG with albumin assumed to be a constant (40 g/l) using the method by Sodergard and colleagues according to the law of mass action) | Baseline to 6 months (End of weight loss phase), percent change<br>Change weight: -7.7 (Mean = 89.1 (SD = 14.8) to 81.4 (14.4)), p<0.0001<br>Androstendione: -0.5 (Mean = 437 (SE = 12.1) to 434 (11.9)), p=0.76<br>Estrone: -5.7 (Mean = 38.1 (SE = 1.1) to 35.9 (1)), p=0.006<br>Estradiol: -9.9 (Mean = 9.4 (SE = 0.3) to 8.4 (0.3)), p<0.0001<br>Testosterone: -3.5 (Mean = 24.6 (SE = 0.9) to 23.7 (0.8)), p=0.13<br>DHEAS: 1.7 (Mean = 47 (SE = 1.8) to 47.8 (1.8)), p=0.31<br>SHBG: 16.2 (Mean = 35 (SE = 1) to 40 (1)), p<0.0001<br>Free Estradiol: -13.4 (Mean = 0.26 (SE = 0.01) to 0.22 (0.01)), p<0.0001<br>Free Testosterone: -9.9 (Mean = 5.3 (SE = 0.2) to 4.8 (0.2)), p<0.0001<br><br>6 to 18 months (Weight gain)<br>Change weight: 2.2 (Mean = 81.4 (SD=14.4) to 83.6 (15.6)), p<0.0001<br>Androstenedione: 0.8 (Mean =434 (SD=11.9) to 438 (12.9)), p=0.68<br>Estrone: -6.4 (Mean=35.9 (SD=1.0) to 33.6 (1.0)), p=0.003<br>Estradiol: 0.5 (Mean =8.4 (SD=0.3) to 8.5 (0.3)), p=0.85<br>Testosterone: 1.1 (Mean =23.7 (SD=0.8) to 24.0 (0.8)), p=0.62<br>DHEAS: -2.5 (Mean =47.8 (SD=1.8) to 46.6 (1.9)), p=0.18<br>SHBG: -8.0 (Mean =40 (SD=1.0) to 37 (1.0)), p<0.0001<br>Free Estradiol: 3.7 (Mean = 0.22 (SD=0.01) to 0.23 (0.01)), p=0.18<br>Free Testosterone: 5.0 (Mean = 4.8 (SD=0.2) to 5.1 (0.2)), p=0.02 |

| | | | |
|---|---|---|---|
| Van Gemert 2015<br>(53)<br>Netherlands<br>RCT<br>Follow-up: 4 months | Sample size n= Diet: 97,<br>Exercise: 98<br><br>Healthy women, BMI: 25-35;<br>Age: 50-59<br><br>Post-menopause | Exposures<br>Waist circumference (cm)<br>Hip circumference (cm)<br>% Body fat (%, DEXA)<br>BMI (kg/m^2)<br>Weight (kg)<br>Fat mass (kg, DEXA)<br><br>Outcomes<br>Estradiol (pg/ml, LC-MS)<br>Estrone (pg/ml, LC-MS)<br>Free Estradiol (pg/ml, calculated by using the total hormone levels, SHBG and a constant for albumin)<br>Testosterone (pg/ml, LC-MS)<br>Androstendione (pg/ml, LC-MS)<br>Free Testosterone (pg/ml, calculated by using the total hormone levels, SHBG and a constant for albumin)<br>SHBG (nmol/l, double-antibody RIA) | Exercise group<br>%Change baseline-16weeks, mean (SD)<br>Fat mass: -15.29 (34 to 28.8 (NR)<br>Estradiol: geomean = 3.69 to 3.22<br>Estrone: geomean = 19.9 to 18.5<br>Free Estradiol: geomean = 0.09 to 0.07<br>Testosterone: geomean = 186 to 172<br>Androstenedione: geomean = 573 to 488<br>Free Testosterone: geomean = 2.44 to 2.01<br>SHBG: geomean = 49.3 to 58.6<br><br>Diet group<br>Fat mass: -10.88 (34 to 30.3 (NR))<br>Estradiol: geomean = 4.2 to 3.62<br>Estrone: geomean = 20.40 to 20.10<br>Free Estradiol: geomean = 0.1 to 0.08<br>Testosterone: geomean = 197 to 189<br>Androstenedione: geomean = 562 to 537<br>Free Testosterone: geomean = 2.53 to 2.25<br>SHBG: geomean = 50.7 to 57.1 |
| Campbell 2012<br>(51)<br>Canada<br>RCT<br>Follow-up: 12 months | Sample size n= Diet: 117,<br>Exercise: 118,<br>Diet+Exercise: 117<br><br>BMI≥25.0 kg/m2 (≥23.0 kg/m2 if Asian-American), no menstrual cycles for > 1 year or follicle-stimulating hormone level of≥23.0 IU/L for women 50 to 59 years of age without a uterus, < 100 minutes/wk moderate-intensity physical activity;<br>Age: diet=58.1 (SD=6.0), exercise=58.1 (SD=5.0), diet+exercise= 58.0 (SD=4.5)<br><br>Post-menopause | Exposures<br>Weight (kg)<br>% Body fat (%, DEXA)<br><br>Outcomes<br>Estrone (pg/ml, RIA)<br>Estradiol (pg/dl, RIA)<br>Testosterone (ng/dl, RIA)<br>SHBG (nmol/l, chemiluminescent immunometric assay using the Immulite Analyzer)<br>Free Estradiol (pg/ml, calculated using the measured values for Estradiol, SHBG, and an assumed constant for | Diet group<br>Change Weight: -10.83 (Mean = 84 (SD = 11.8) to 74.9 (12.3))<br>Change % Body fat: -10.43 (Mean = 47 (SD = 4.3) to 42.1 (6.4))<br>Estrone: geomean = 35.2 (95% CI = 32.7 to 37.9) to 31.8 (95% CI = 29.4 to 34.4)<br>Estradiol: geomean = 11.6 (95% CI = 10.7 to 12.5) to 9.7 (95% CI = 8.9 to 10.6)<br>Testosterone: geomean = 23.9 (95% CI = 21.9 to 26.0) to 23.6 (95% CI = 21.6 to 25.8)<br>SHBG: geomean = 35.8 (95% CI = 33.0 to 38.8) to 43.8 (95% CI = 40.4 to 47.5)<br>Free Estradiol: geomean = 0.31 (95% CI = 0.28 to 0.34) to 0.24 (95% CI = 0.22 to 0.27)<br>Androstenedione: geomean = 51.1 (95% CI = 47.1 to 55.3) to 51.8 (95% CI = 47.7 to 56.2)<br>Free Testosterone: geomean = 5.1 (95% = 4.7 to 5.6) to 4.6 (95% CI = 4.2 to 5.1)<br><br>Exercise group<br>Change Weight: -3.345 (Mean = 83.7 (SD = 12.3) to 80.9 (12.2))<br>Change % Body fat: -3.805 (Mean = 47.3 (SD = 4.1) to 45.5 (5))<br>Estrone: geomean = 34.8 (95% CI = 32.4 to 37.4) to 32.9 (95% CI = 30.5 to 35.5)<br>Estradiol: geomean = 11.5 (95% CI = 10.6 to 12.5) to 11 (95% CI = 10.1 to 11.9)<br>Testosterone: geomean = 24.8 (95% CI = 23.0 to 26.7) to 23.6 (95% CI = 21.6 to 25.7)<br>SHBG: geomean = 39.1 (95% CI = 35.9 to 42.6) to 38.8 (95% CI = 35.6 to 42.4)<br>Free Estradiol: geomean = 0.3 (95% CI = 0.27 to 0.33) to 0.29 (95% CI = 0.26 to 0.32) |

| | | | |
|---|---|---|---|
| | | albumin)<br>Androstendione (ng/dl, RIA)<br>Free Testosterone (pg/ml, calculated using the measured values for Total Testosterone, SHBG, and an assumed constant for albumin) | Androstenedione: geomean = 50.2 (95% CI = 46.6 to 54.1) to 49.6 (95% CI = 45.6 to 54.0)<br>Free Testosterone: geomean = 5.1 (95% CI = 4.7 to 5.5) to 4.9 (95% CI = 4.5 to 5.3)<br><br>Diet & Exercise group<br>Change Weight: -11.88 (Mean = 82.5 (SD = 10.8) to 72.7 (10.9))<br>Change % Body fat: -13.29 (Mean = 47.4 (SD = 4.5) to 41.1 (7))<br>Estrone: geomean = 33.9 (95% CI = 31.5 to 36.6) to 30.2 (95% CI = 28.0 to 32.5)<br>Estradiol: geomean = 11.5 (95% CI = 10.6 to 12.5) to 9.2 (95% CI = 8.4 to 10.0)<br>Testosterone: geomean = 23.9 (95% CI = 22.1 to 25.8) to 22.5 (95% CI = 20.8 to 24.3)<br>SHBG: geomean = 34.1 (95% CI = 31.9 to 36.4 ) to 42.9 (95% CI = 40.2 to 45.6 )<br>Free Estradiol: geomean = 0.32 (95% CI = 0.29 to 0.35 ) to 0.23 (95% CI = 0.21 to 0.26)<br>Androstenedione: geomean = 52.6 (95% CI = 49.1 to 56.4) to 50.8 (95% CI = 47.1 to 54.7)<br>Free Testosterone: geomean = 5.3 (95% CI = 4.9 to 5.7) to 4.5 (95% CI = 4.1 to 4.8) |
| Carpenter 2012<br>(6)<br>USA<br>weight-loss lifestyle intervention<br>Follow-up: 3 months | Sample size n= 7<br><br>Healthy obese postmenopausal women,<br><br>BMI: 33.6 kg/m2(SD=5.3)<br><br>Age: 55.3 (SD=5.5)<br><br>Post-menopause | Exposures<br>BMI (kg/m2), fat mass (kg, DEXA)<br><br>Outcomes<br>Estrone sulfate, nmol/L, RIA<br>Estradiol, pmol/L, EIA | Non-periodized resistence training<br>Baseline to 3 Months, mean (SD), change<br>fat mass: 41.3 (11.9) to 35.5 (10.9), -14.4 (-18.5,-10.3), S<br>BMI: 33.6 (5.3) to 29.8 (3.8), -11.0 (-13.9,-8.1), S<br>Weight: 89.8 (15.3) to 80.3 (12.2), -10.3 (-12.6, -8.0), S<br><br>Estrone sulfate: 5.8 (1.8) to 4.9 (2.8), -9.9 (-54.2,34.3), NS<br>Estradiol: 157.9 (49.7) to 101.6 (27.9), -24.4 (-65.7, 17.0), NS<br><br>Not reported |
| Rock 2016<br>(62)<br>USA<br>RCT<br>Follow-up: 12 months | Sample size n= low fat: 68, walnut rich: 65, low carbohydrate: 61<br><br>Obese women, BMI/Weight: only change reported;<br>Age: low fat group=50 (range=25-68), walnut rich=50 (range=22-67), low carboydrate=50 (range=25-72)<br><br>STRAT PRE POST for estradiol only but estradiol not reported. They just mentioned no change in estradiol | Exposures<br>Weight (kg)<br><br>Outcomes<br>SHBG (nmol/l, chemiluminescent sandwich techniques) | Low fat insulin sensitive<br>Change Weight: -8.651 (Mean = 86.7 (SD = 10) to 79.2 (7.9))<br>SHBG: Mean = 63 (SD = 32.7) to 88 (7)<br><br>Wallnut Insulin sensitive<br>Change weight: -9.278 (Mean = 87.3 (SD = 10.9) to 79.2(7.1))<br>SHBG: Mean = 63 (SD = 32.7) to 80 (6)<br><br>Low carb insulin sensitive<br>Change weight: -4.92 (Mean = 87.4 (SD = 13.7) to 83.1 (6.1))<br>SHBG: Mean = 63 (SD = 32.7) to 71 (7) |
| Longitudinal | | | |
| Goss 2012<br>(11)<br>USA<br>Cohort<br>Follow-up: 24 months | Sample size n= 18<br><br>Healthy women, post-menopausal > 6months, experienced a natural menopause, BMI: 25.6 (SD=3.9); Age: 51.2 (SD=2.8)<br><br>Post-menopause | Exposures<br>IAAT (cm2, CT)<br><br>Outcomes<br>Estradiol (pg/ml, RIA or IRMA)<br>Estrone (pg/ml, RIA or IRMA)<br>Free Testosterone (pmol/L, RIA or IRMA) | Significant change<br>IAAT: (Mean = 115.2 (SD = 41.9) to 127.4 (52.4), 10.6%<br><br>Change in biomarkers - no mentionned if significant we took 10% threshold:<br>Estradiol: Mean = 9.1 (SD = 6.3) to 13.6 (18.8), 50%<br>Estrone: Mean = 39.5 (SD = 35.8) to 48.1 (68.2), 21%<br>Free Testosterone: Mean = 6.9 (SD = 3.4) to 7.3 (4.9), 7%<br>Androstenedione: Mean = 1.2 (SD = 0.6) to 0.9 (0.4), -25%<br>DHEAS: Mean = 84.1 (SD = 47.1) to 82.3 (42.4), -2% |

| | | | |
|---|---|---|---|
| | | Androstendione (ng/ml, RIA or IRMA)<br>DHEAS (µg/dl, RIA or IRMA)<br>Testosterone (ng/dl, RIA or IRMA)<br>SHBG (ng/ml, RIA or IRMA) | Testosterone: Mean = 21.1 (SD = 11.2) to 21.1 (6.8), 0%<br>SHBG: Mean = 89.8 (SD = 32.7) to 97.6 (58.8), 8.6% |
| Observational studies – Direct measures of adiposity | | | |
| Critchlow 2025<br>(75)<br>Australia<br>Cross-sectional +<br>Cohort<br>Follow-up: follow up visit 4–6 years months | Sample size n= Females cross-sectional (n = 319)<br><br>Females > 50 years longitudinal analysis (n = 83) follow up visit 4–6 years later<br><br>Data collected between 2003 and 2023<br><br>Females cross-sectional<br>Age mean (SD)=60.20 (15.95), range=24.00–89.00<br>BMI=26.51 (5.02), 17.83–45.74<br><br>Females > 50 years longitudinal analysis<br>Age=<br>t0=64.41 (10.99)<br>t1=69.24 (10.95)<br>BMI<br>t0=27.23 (4.62)<br>t1=27.34 ( 5.13)<br><br>PRE + POST Mix | Exposures<br>Total body fat mass (TBFM, kg, whole-body DXA)<br>subcutaneous fat area (cm2),<br>Intramuscular fat area (cm2),<br><br>Outcomes<br>Fasted (12 h)<br><br>Total testosterone (nmol/L, high-performance liquid chromatography–tandem mass spectrometry (LC/MS))<br><br>SHBG (nmol/L, immunoradiometric assay)<br>bioavailable testosterone (nmol/L, ammonium sulphate was used to separate SHBG-bound hormones from albumin-bound and free hormones) | Multiple and binomial linear regression was used to map cross-sectional (n=319) and longitudinal (n=83) associations<br><br>Adjusted cross-sectional<br>BioT was positively associated with total body fat percentage (β = 0.18, $p < 0.05$).<br>TT was negatively associated with total body fat percentage (β = −0.14, $p < 0.05$).<br><br>Adjusted longitudinal<br>Decreases in TT and BioT were associated with an increase in total body fat (β = −0.25 for both, $p < 0.05$)<br><br>adjusted for age, ethnicity, height, ACCI, total physical activity levels in the past 2 weeks, and time between the baseline and follow-up visit. Physical function measures were only assessed in females ≥ 50 years old |
| Yuan 2025<br>(76)<br>USA<br>Cross-sectional (+MR)<br>Follow-up: months | Sample size n= 2309 women<br><br>NHANES data from 2013 to 2016<br><br>2309 Women age (mean (se)) 30.5 (0.3)<br><br>BMI 28.1 (0.2) | Exposures<br>hole-body dual-energy X-ray absorptiometry (DXA) scans<br><br>Android/gynoid measurements<br>Android fat mass, gram<br>Gynoid fat mass, gram<br>Subcutaneous fat mass, | SHBG behave as fairly consistent protective factors against adiposity in women (negative associations), while testosterone and FAI show more complex, non-linear (inverted U-shaped) relationships rather than straightforward negative or positive trends.<br><br>No significant association between total testosterone levels and whole-body adiposity measures and body measurements ($P > 0.05$), and the trends were inconsistent across tertile levels.<br><br>Inverted U-shaped associations of both total testosterone and FAI with obesity-related indicators were observed, which were consistent with the results of the two models. |

| | | | |
|---|---|---|---|
| | premenopausal females<br>(menstrual cycle unknown) | gram<br>Total abdominal fat mass, gram<br>Visceral adipose tissue mass, gram<br><br>Whole body measurements<br>Trunk fat, gram<br>Total fat, gram<br><br>Outcomes<br>Testosterone (ng/dL, isotope dilution liquid chromatography–tandem mass spectrometry)<br><br>SHBG (nmol/L, chemiluminescent detection)<br><br>FAI=TT/SHBG | SHBG showed an inverse trend in the smoothed curves with the obesity-related indicators, and was significantly negatively correlated with 16 indicators at the 1 st, and 2nd tertile levels in both models ($P < 0.05$).<br><br>Baseline measures in women:<br>Total testosterone, mean(SE) =26.2 (0.4)<br>SHBG= 66.3 (1.0)<br>FAI= 1.9 (0.04)<br>Android fat mass, gram=2243.4 (26.9)<br>Gynoid fat mass, gram=5030.6 (39.5)<br>Subcutaneous fat mass, gram=1828.7 (16.7)<br>Total abdominal fat mass, gram=2227.5 (20.9)<br>Visceral adipose tissue mass, gram=398.8 (5.5)<br>Trunk fat, gram=12,914.0 (133.3)<br>Total fat, gram=28,496.6 (248.6)<br><br>Multivariate linear regression (Cross-sectional)<br>Tertile Q3 vs Q1, β (95%CI), p-value<br><br>Testosterone:<br>Android/gynoid measurements<br>Android fat mass<br>Model 1: 0.479(-1.136,2.094), 0.55<br>Model 2: 1.211(-0.226,2.647). 0.09<br><br>Gynoid fat mass<br>Model 1: 0.762(-1.151,3.037), 0.50<br>Model 2: 1.750(-0.361,3.861),0.10<br><br>Subcutaneous fat mass<br>Model 1:0.724(-0.278,1.726), 0.15<br>Model 2:1.200(0.237,2.163), 0.01<br><br>Total abdominal fat mass<br>Model 1: 0.303(-0.837,1.442), 0.59<br>Model 2:0.931(-0.107,1.970), 0.08<br><br>Visceral adipose tissue mass<br>Model 1: -0.422(-0.994,0.151), 0.14<br>Model 2: 0.303(-0.837,1.442), 0.59<br><br>Whole body measurements<br>Trunk fat<br>Model 1: 0.567(-8.082,9.216),089<br>Model 2:4.318(-3.519,12.154),0.27<br><br>Total fat |

| | | | |
|---|---|---|---|
| | | | Model 1:2.353(-10.411,15.118),0.71<br>Model 2:9.339(-2.211,20.890),0.11<br><br>SHBG<br>Android/gynoid measurements<br>Android fat mass<br>Model 1:0.711(-1.388,2.809), 049<br>Model 2: 0.781(-0.991,2.552), 0.37<br><br>Gynoid fat mass<br>Model 1: 1.730(-0.810,4.269),0.17<br>Model 2: 1.429(-0.832,3.689), 0.21<br><br>Subcutaneous fat mass<br>Model 1: 0.713(-0.628,2.053), 0.29<br>Model 2: 0.711(-0.423,1.845), 0.21<br><br>Total abdominal fat mass<br>Model 1: 0.639(-1.034,2.312), 0.44<br>Model 2:0.707(-0.689,2.103), 0.31<br><br>Visceral adipose tissue mass<br>Model 1: -0.074(-0.457,0.310), 0.70<br>Model 2: -0.004(-0.325,0.317), 0.98<br><br>Whole body measurements<br>Trunk fat<br>Model 1: 0.679(-5.612,14.970), 0.36<br>Model 2: 4.993(-3.691,13.677), 0.25<br><br>Total fat<br>Model 1: 9.956(-7.798,27.709), 0.26<br>Model 2: 9.812(-5.333,24.957), 0.19<br><br>Model 1: adjusted for age and ethnicity<br><br>Model 2: adjusted for age, ethnicity, education level, poverty status, smoking status, HEI-2015, alcohol consumption and leisure-time physical activity |
| Ma 2024<br>(77)<br>USA<br>Cross-sectional<br>Follow-up: months | Sample size n= 1525 women<br><br>NHANES 2011-2016<br><br>1525 women aged mean= 35.7 (SD= 12.3) years old<br><br>BMI=29.0 (7.7) | Exposures<br>total percent fat (TPF), android percent fat (APF), gynoid percent fat (GPF), android to gynoid ratio (A/G) and lean mass percent (LMP). | Multivariable linear regression models β (95%CI)<br>TPF (%) — Total percent fat<br><br>Model 1: 0.08 (−0.05, 0.21), P = 0.24<br>Model 2: 0.19 (0.06, 0.33), P < 0.01<br>Model 3: 0.20 (0.07, 0.33), P < 0.01 |

| | | | |
|---|---|---|---|
| | Unknown | DXA<br><br>Outcomes<br>Testosterone (ng/dL, isotope dilution liquid chromatography tandem mass spectrometry (ID-LC-MS/MS) | APF (%) — Android percent fat<br><br>Model 1: 0.08 (−0.01, 0.18), P = 0.09<br>Model 2: 0.15 (0.05, 0.25), P = 0.0043<br>Model 3: 0.17 (0.07, 0.26), P < 0.01<br><br>GPF (%) — Gynoid percent fat<br><br>Model 1: 0.08 (−0.08, 0.24), P = 0.31<br>Model 2: 0.15 (−0.01, 0.31), P = 0.06<br>Model 3: 0.15 (−0.00, 0.30), P = 0.06<br><br>testosterone levels positively associated with body fat percentage and negatively associated with muscle mass,<br><br>Model 1: no adjustement<br><br>Model 2: age, race<br><br>Model 3: age, race, hypertension, diabetes, hypercholesteremia, smoking status, and vigorous work activity |
| Leenen 1994<br>(3)<br>Netherlands<br>Non-randomized intervention<br>Follow-up: 6 months | Sample size n= 33<br><br>Healthy obese women, BMI: 28-38;<br>Age: 39 (SD=5)<br><br>Pre-menopause<br>(Follicular) | Exposures<br>VAT (cm2, MRI)<br><br>Outcomes<br>SHBG (nmol/l, IRMA,)<br>Estrone (pmol/l, RIA)<br>Estradiol (pmol/l, RIA)<br>Free Estradiol (pmol/l, calculated)<br>Testosterone (nmol/l, RIA)<br>Free Testosterone (pmol/l, calculated)<br>DHEAS (µmol/l, RIA)<br>Androstenedione (nmol/l, RIA) | Pearson correlation (adjusted for age and fat mass):<br>SHBG:VAT (r = -0.51, p<0.01)<br>Estrone: VAT (r = -0.14, not significant)<br>Estradiol: VAT (r = -0.31, not significant)<br>Free Estradiol: VAT (r = -0.21, not significant)<br>Testosterone: VAT (r = -0.01, not significant)<br>Free Testosterone: VAT (r = 0.35, p<0.05)<br>DHEAS: VAT (r = -0.01, not significant)<br>Androstenedione: VAT (r = 0.19, not significant)<br>VAT:free Estradiol/free Testosterone (r = -0.36, p<0.05)<br><br>Pearson correlation adujsted for age and fat mass |
| Sowers 2001<br>(10)<br>USA<br>Cohort<br>Follow-up: 36 months | Sample size n= 511<br><br>Women who are offsprings of the Tecumseh Community Health Study or resident of Tecumseh (Michigan), no parent in original study, BMI: 26.8 (6.2);<br>Age: 38 (range=25-45)<br><br>PRE + PERI Mix<br>(Follicular) | Exposures<br>Fat mass (kg, DEXA)<br>% Body fat (DEXA)<br>BMI (kg/m2, clinician)<br><br>Outcomes<br>Testosterone (pg/ml, RIA) | Significant increases in percent body fat , weight, and BMI over the 3years period (p-trend<0.05)<br><br>Mean total testosterone concentrations (pg/ml) by category of body composition<br>%Body fat in categories ≤34, >34 to ≤42, >42 to ≤48, >48<br><br>1992–1993: mean 170, 192, 226, 268<br>1993–1994: 174, 205, 211, 248<br>1994–1995: 201, 205, 231, 247<br>significant trend in testosterone mean across %body fat and fat mass p<0.01 |

| | | | |
|---|---|---|---|
| Goodfriends 1999<br>(2)<br>USA<br>Non-Randomized<br>Intervention<br>Follow-up: months | Sample size n= 28,<br>obese: 21,<br>lean: 7<br><br>Obese and lean women, who had maintained a stable body weight for at least several months prior to participation, BMI: >27;<br>Age: 35 +/- 2 years (lean women), 37 +/- 1 year (obese women)<br><br>Pre-menopause<br>(Unknown) | Exposures<br>Fat mass (kg, DEXA)<br>VAT (cm2,CT)<br><br>Outcomes<br>DHEAS (ng/dl, RIA) | Pearson correlation:<br>DHEAS: VAT (r = 0.36, p = 0.2)<br><br>Not reported |
| Tchernof 1999<br>(78)<br>USA<br>cross-sectional<br>Follow-up: months | Sample size n= 52<br><br>Premenopausal women, BMI: 23.3 (SEM=0.5);<br>Age: 46.7 (SEM=0.4)<br><br>Pre-menopause<br>(Unknown) | Exposures<br>VAAT (cm2, DEXA)<br>SAAT (kg, DEXA)<br>Fat mass (kg, DEXA)<br><br>Outcomes<br>SHBG (nmol/l, RIA),<br>Testosterone (ng/ml, RIA) | Pearson correlation:<br>VAAT:SHBG (r = -0.39; p<0.008)<br>SAAT:SHBG (r = -0.37; p<0.005)<br>Fat mass:SHBG (r = -0.41; p<0.005)<br>Free androgen index (testosterone/SHBG * 100) same as SHBG<br><br>Adiposity measures * Testosterone, NS |
| Garaulet 2000<br>(79)<br>Spain<br>cross-sectional<br>Follow-up: months | Sample size n= 22<br><br>Premenopausal obese women admitted for abdominal surgery or laparoscopy, BMI: 27-35;<br>Age: 36 (SD=8)<br><br>Pre-menopause<br>(Mix) | Exposures<br>Subcutaneous adipocyte diameter (µm, microscopy)<br><br>Outcomes<br>Androstendione (ng/ml, RIA)<br>DHEAS (µg/dl, RIA) | Significant correlation between subcutaneous adipocyte diameter and androstenedione (r=0.51, p=0.015)<br>Significant correlation between subcutaneous adipocyte diameter and DHEAS (r=0.44, p=0.041)<br><br>Not reported |
| Puder 2006<br>(80)<br>USA<br>cross-sectional<br>Follow-up: months | Sample size n= 38,<br>regularly menstruating exercising control women: 19,<br>regularly menstruating normally active control women: 19<br><br>Two age- and body mass index-matched groups: regularly menstruating exercising control women, BMI: 21.7 (SD=0.5) exercising control, 21.6 (SD=0.3) normally active control;<br>Age: 25 (SD=1.3) exercising control, 24 (SD=1) normally active control<br>The third group was excluded because this group contains | Exposures<br>Trunk to extremity fat ratio (DEXA)<br>% Trunk fat (DEXA)<br>% Extremity fat (DEXA)<br><br>Outcomes<br>Estradiol (pg/ml, solid-phase chemiluminescent immunoassay) | Pearson correlation for all women (both control groups):<br>significat correlation between Trunk to Extremity fat ratio and Estradiol (r=-0.4, p=0.02)<br>significant correlation between % Trunk fat and Estradiol (r=-0.39, p=0.02)<br>significant correlation between % Extremity fat and Estradiol (r=0.41, p=0.02)<br><br>Pearson correlation for regularly exercising control women:<br>Trunk to Extremity fat ratio:Estradiol (r = -0.54, p=0.02)<br>% Trunk fat:Estradiol (r = -0.52, r = 0.03)<br>% Extremity fat:Estradiol (r = 0.05, p=0.02)<br><br>Not reported |

| | | | |
|---|---|---|---|
| | women with exercise-induced amenorrhea.<br><br>Pre-menopause<br>(Follicular) | | |
| Nutter 1991<br>(81)<br>USA<br>cross-sectional<br>Follow-up: months | Sample size n= 15<br><br>Normal cycling women who were running 10 to 35 miles per week, Weight: 60.27 (SEM=1.63);<br>Age: 25.8 (SEM=0.8)<br><br>Pre-menopause<br>(Luteal) | Exposures<br>% Body fat (hydrostatic weighing)<br><br>Outcomes<br>Beta-17-Estradiol (pg/ml, RIA) | Pearson correlation:<br>non significant correlation between % Body fat and beta-17-Estradiol ($r=0.27$, $p\geq0.05$)<br><br>Not reported |
| Thurston 2008<br>(16)<br>USA<br>cross-sectional<br>Follow-up: months | Sample size n= 461,<br>Women who reported hot flashes: 217<br>Women without hot flashes: 244<br><br>Women with intact uterus, at least one ovary and who had menstruated within the past 3 months, women reporting and not reporting hot flashes, BMI: 29.8 (SD=6.1) with hot flashes, 28.4 (SD=6.1) without hot flahes;<br>Age: 50.8 (SD=2.9) with hot flashes, 50.0 (SD=2.8) without hot flahes<br><br>Pre-menopause<br>(Follicular) | Exposures<br>VAAT (not reported, CT)<br>SAAT (not reported, CT)<br>TAAT (not reported, CT)<br><br>Outcomes<br>Estradiol (pg/ml, double-antibody chemiluminscent immunoassay)<br>Free estradiol index (caculated) | Linear regression (beta coefficients):<br>VAAT:Estradiol (b=-0.08, p=0.11)<br>VAAT:free Estradiol index (b=0.14 p=0.009)<br>TAAT:Estradiol (b=-0.07, p=0.19)<br>TAAT:free Estradiol index (b=0.14, p=0.03)<br>SAAT:Estradiol (b=0.05, p=0.30)<br>SAAT:free Estradiol index (b=0.12, p=0.03)<br><br>Adjusted for age, site, race/ethnicity, education, smoking, hormone use, and cycle day of blood draw |
| Utz 2008<br>(82)<br>USA<br>cross-sectional<br>Follow-up: months | Sample size n= 34,<br>lean: 11,<br>overweight: 12,<br>obese: 11<br><br>Eumenhorreic premenopausal women, BMI: <25 (lean), >= 25 and < 30 (overweight), >= 30 (obese);<br>Age: 30.7 (SEM = 1.3)<br><br>Pre-menopause<br>(Follicular) | Exposures<br>VAT (mm2, CT), IAAT (mm2, CT), SAT (mm2, CT)<br>Fat mass (kg, DEXA)<br><br>Outcomes<br>Estradiol (pg/ml, RIA), Testosterone (ng/dl, RIA), Estrone (pg/ml, RIA).<br>Free Testosterone was calculated from total Testosterone and SHBG by the laws of mass action | Spearman correlation:<br>VAT:Estradiol (r = 0.04, p = 0.42)<br>VAT:Testosterone (r=0.20, p=0.25)<br>VAT:free Testosterone (not reported)<br>VAT:Estrone (r=0.27, p=0.11)<br>Fat mass:Estradiol (r=0.08, p=0.66)<br>Fat mass:Estrone (r=0.31, p=0.07)<br>Fat mass:Testosterone (r=0.20, p=0.25)<br>Fat mass:free Testosterone (r=0.31, p=0.08)<br>IAAT:Estradiol (r=0.14, p=0.44)<br>IAAT:Estrone (r=0.38, p=0.03)<br>IAAT:Testosterone (r=0.11, p=0.38)<br>IAAT:free Testosterone (r=0.38, p=0.03)<br>SAT:free Testosterone (r=0.36, p=0.04)<br>SAT:Estradiol (r=0.14, p=0.42)<br>SAT:Estrone (r=0.41, p=0.02) |

| | | | |
|---|---|---|---|
| | | | SAT:Testosterone (r=0.20, p=0.25)<br><br>Not reported |
| Nayeem 2009<br>(83)<br>USA<br>cross-sectional<br>Follow-up: months | Sample size n= 241<br><br>Premenopausal women, BMI: 28.3 (95%CI=27.7, 29.0);<br>Age: 36.2 (95%CI=35.9, 36.6)<br><br>Pre-menopause<br>(Luteal) | Exposures<br>Fat mass (kg, DEXA)<br><br>Outcomes<br>SHBG (nmol/l, ELISA) | Pearson correlation:<br>Fat mass:SHBG (r = -0.38, p>0.0001)<br><br>Not reported |
| Yeung 2010<br>(84)<br>USA<br>cross-sectional<br>Follow-up: months | Sample size n= 246,<br>white women: 146,<br>non-white women: 100<br><br>Multiethnic premenopausal women (White women, non-white women including black, asian and other races), 33% of the women were overweight or obese (BMI > 25 kg/m2);<br>Age: 18 - 44<br><br>Pre-menopause<br>(Follicular) | Exposures<br>% Body fat (DEXA)<br>% Trunk fat (DEXA)<br>Trunk to leg fat ratio (calculated by dividing trunkal fat mass by leg fat mass)<br><br>Outcomes<br>SHBG (not reported, not reported) | Regression (beta-coefficient):<br>White women: % Body fat:SHBG (b = -0.031, SEM = 0.006, p<0.05)<br>Non-white women: % Body fat:SHBG (b = -0.016, SEM = 0.007, p<0.05)<br>White women: % Trunk fat:SHBG (b = -0.029, SEM = 0.004, p<0.05)<br>Non-white women: % Trunk fat:SHBG (b = -0.020, SEM = 0.006, p<0.05)<br>White women: Trunk to leg fat ratio:SHBG (b = -0.363, SEM = 0.057, p<0.05)<br>Non-white women: Trunk to leg fat ratio:SHBG (b = -0.474, SEM = 0.090, p<0.05)<br><br>Spearman correlation:<br>White women: % Body fat:SHBG (r = -0.35, p<0.05)<br>Non-white women: % Body fat:SHBG (r = -0.13, not significant)<br>White women: % Trunk fat:SHBG (r = -0.43), p<0.05)<br>Non-white women: % Trunk fat:SHBG (r = -0.22, p<0.05)<br>White women: Trunk to leg fat ratio:SHBG (r = -0.43, p<0.05)<br>Non-white women: Trunk to leg fat ratio:SHBG (r = -0.34, p<0.05)<br><br>Adjusted for age |
| Keller 2011<br>(85)<br>USA<br>cross-sectional<br>Follow-up: months | Sample size n= 30<br><br>Non-obese, healthy, premenopausal, cycling women, BMI: 23 (SE=0.5);<br>Age: 27.3 (SE=0.8)<br><br>Pre-menopause<br>(Luteal) | Exposures<br>% Body fat (DEXA), Fat mass (kg, DEXA), TAAT (kg, DEXA)<br>VAT (cm2, CT)<br><br>Outcomes<br>DHEA (ng/ml, RIA), Andorstenedione (pg/ml, RIA), free Testosterone (pg/ml, RIA), Testosterone (ng/dl, RIA)<br>SHBG (nmol/l, chemiluminescence immunoanalysis),<br>DHEAS (µg/ml, chemiluminescence immunoanalysis) | Pearson correlation:<br>% Body fat:DHEA (r = 0.195, p≥0.05)<br>% Body fat:Androstenedione (r = 0.097, p≥0.05)<br>% Body fat:SHBG (r = 0.325, p≥0.05)<br>% Body fat:free Testosterone (r = 0.162, p≥0.05)<br>% Body fat:Testosterone (r = 0.354, p≥0.05)<br>Fat mass:Testosterone (r = 0.377, p=0.04)<br>Fat mass:DHEA (r = 0.153, p≥0.05)<br>Fat mass:SHBG (r = 0.252, p≥0.05)<br>Fat mass:DHEAS (r = -0.058, p≥0.05)<br>TAAT:Testosterone (r = 0.192, p≥0.05)<br>TAAT:free Testosterone (r = 0.167, p≥0.05)<br>TAAT:DHEAS (r = -0.09, p≥0.05)<br>TAAT:Androstenedione (r = -0.113, p≥0.05)<br>TAAT:SHBG (r = 0.006, p≥0.05)<br>VAT:free Testosterone (r = -0.120, p≥0.05)<br>VAT:DHEA (r = -0.569, p=0.02)<br>Fat mass:free Testosterone (r = 0.226, p≥0.05)<br>VAT:DHEAS (r = -0.377, p≥0.05)<br>% Body fat:DHEAS (r = -0.033, p≥0.05) |

| | | | |
|---|---|---|---|
| | | | Fat mass:Androstenedione (r = 0.036, p≥0.05)<br>VAT:Androstenedione (r = -0.410, p≥0.05)<br>VAT:Testosterone (r = -0.375, p≥0.05)<br>VAT:SHBG (r = -0.380, p≥0.05)<br>TAAT:DHEA (r = -0.087, p≥0.05)<br><br>Adjusted for age |
| Azrad 2012<br>(86)<br>USA<br>Cohort (longitunidal study, but here only values for baseline)<br>Follow-up: months | Sample size n= 107<br><br>Premenopausal women with normal menstrual cylces, BMI: 27-40;<br>Age: 34.3 ± 0.62<br><br>Pre-menopause<br>(Follicular) | Exposures<br>IAAT (cm2, CT)<br>Fat mass (kg, DEXA)<br><br>Outcomes<br>SHBG (nmol/l, IRMA) | Simple correlation:<br>IAAT:SHBG (r=-0.43, p=0.0001)<br>Fat mass:SHBG (r=-0.191, p=0.05)<br><br>Not reported |
| Aydin 2013<br>(36)<br>Turkey<br>cross-sectional<br>Follow-up: months | Sample size n= 28<br><br>Age and BMI-matched healthy controls for interventional study on PCOS, BMI: 21.0 (SD=2.4);<br>Age: 22.6 (SD=1.9)<br><br>Pre-menopause<br>(Follicular) | Exposures<br>Trunk fat (kg, BIA)<br>% Trunk fat (BIA)<br>Fat mass (kg, BIA)<br>% Body fat (BIA)<br><br>Outcomes<br>Free Androgen Index (calculated)<br>SHBG (nmol/l, IRMA) | Pearson correlation:<br>Trunk fat:Free Androgen Index (r=0.215, p≥0.05)<br>% Trunk fat:free Androgen Index (r=0.244, p≥0.05)<br>Fat mass:free Androgen Index (r=0.210, p≥0.05)<br>% Body fat:free Androgen Index (r=0.255, p≥0.05)<br><br>Trunk fat:SHBG (r=-0.339, p≥0.05)<br>% Trunk fat:SHBG (r=-0.367, p≥0.05)<br>Fat mass:SHBG (r=-0.354, p≥0.05)<br>% Body fat:SHBG (r=-0.356, p≥0.05)<br><br>Not reported |
| Li 2016<br>(23)<br>China<br>cross-sectional<br>Follow-up: months | Sample size n= 103<br><br>Age- and race-matched healthy premenopausal controls for study on PCOS women, BMI: 20.3 (SD=2.5);<br>Age: 25.8 (SD=2.3)<br><br>Pre-menopause<br>(Unknown) | Exposures<br>% Body fat (BIA)<br><br>Outcomes<br>Free Androgen Index (calculated) | Simple correlation:<br>% Body fat * free Androgen Index (r=0.439, p<0.001)<br><br>Multiple adjusted regression with FAI as dependent variable (adjusted for DHEAS, LH/FSH, Irisin:<br>% Body fat * free Androgen Index (b = 0.013, p<0.001)<br><br>Not reported |
| Peiris 1989<br>(87)<br>USA<br>cross-sectional<br>Follow-up: months | Sample size n= 31<br><br>Healthy premenopausal women, Weight: 89.4 (SD=20.7);<br>Age: 32.8 (SD=3.4)<br><br>Pre-menopause<br>(Follicular) | Exposures<br>Fat mass (kg, hydrostatic weighing)<br><br>Outcomes<br>SHBG (nmol/l, displacement technique) | Linear regression:<br>Fat mass * SHBG (r=-0.51, p<0.01)<br>Fat mass * Testosterone (r=-0.26, p<0.01)<br><br>Not reported |

| | | | |
|---|---|---|---|
| | | Testosterone (nmol/l, IRMA) | |
| Ambroziak 2017 (12)<br>Poland<br>cross-sectional<br>Follow-up: months | Sample size n= 29<br><br>Healthy premenopausal women controls for study on PCOS women, BMI: 23.3 (SD=5.5); Age: 27.9 (SD=7.5)<br><br>Pre-menopause<br>(Follicular) | Exposures<br>% Body fat (BIA, DEXA)<br><br>Outcomes<br>Testosterone (nmol/l, LC-MS/MS)<br>free Androgen Index (calculated)<br>Dihydrotestosterone (nmol/l, LC-MS/MS)<br>Androstenedione (nmol/l, LC-MS/MS)<br>DHEAS (µmol/l, LC-MS/MS)<br>T/DHT ratio (LC-MS/MS) | Pearson correlation:<br>% Body fat*Testosterone (p>0.05)<br>% Body fat*free Androgen Index (p>0.05)<br>% Body fat*Dihydrotestosterone (p>0.05)<br>% Body fat*Androstenedione (p>0.05)<br>% Body fat*DHEAS (p>0.05)<br>% Body fat*T/DHT Index (r=0.64, p<0.001)<br><br>Not reported |
| Marchand 2018 (32)<br>Canada<br>cross-sectional<br>Follow-up: months | Sample size n= 42,<br>for SAAT and VAAT: 41,<br>for Mean visceral adipose diameter: 28<br>for Mean subcutaneous adipose diameter: 36<br><br>Premenopausal women receiving hysterectomy, BMI: 27.1 (SD=4.2); Age: 45.8 (SD=3.4)<br><br>Pre-menopause<br>(Mix) | Exposures<br>Adipocyte visceral mean diameter (µm, microscopy),<br>Adipocyte subcutaneous mean diameter (µm, microscopy),<br>VAAT (not reported, CT),<br>SAAT (not reported, CT),<br>% Body fat (measuement: DEXA)<br><br>Outcomes<br>Dihydrotestosterone (nM, ESI-LC_MS/MS)<br>DHEAS (nM, ESI-LC_MS/MS)<br>DHEA (nM, ESI-LC_MS/MS)<br>Androstenedione (nM, ESI-LC_MS/MS)<br>Androsterone (nM, ESI-LC_MS/MS) | Pairwise correlation:<br>Mean visceral adipocyte diameter:<br>Dihydrotestosterone (r=-0.36, p≤0.1)<br>DHEA (r=-0.24, NS)<br>Androstenedione (r=-0.45, p≤0.05)<br>Androsterone (r=-0.34, p≤0.1)<br>Testosterone (r=-0.48, p≤0.01)<br>DHEAS (r=-0.21, NS)<br><br>Mean subcutaneous adipocyte diameter:<br>Dihydrotestosterone (r = -0.15, NS)<br>DHEA (r = -0.13, NS)<br>Androstenedione (r = -0.30, NS)<br>Androsterone (r = -0.28, NS)<br>Testosterone (r = -0.29, NS)<br>DHEAS (r = -0.20, NS)<br><br>VAAT:<br>Dihydrotestosterone (r=-0.16, NS)<br>Androsterone (r=-0.26, , p≤0.1)<br>DHEAS (r=-0.05, NS)<br>DHEA (r=-0.17, NS)<br>Androstenedione (r=-0.35, , p≤0.05)<br>Testosterone (r=-0.26, p≤0.1)<br><br>SAAT:<br>Dihydrotestosterone (r = -0.27, NS) |

| | | | |
|---|---|---|---|
| | | Testosterone (nM, ESI-LC_MS/MS) | Androsterone (r = -0.30, p≤0.05)<br>DHEAS (r = -0.20, NS)<br>DHEA (r = -0.28, p≤0.1)<br>Androstenedione (r = -0.33, p≤0.05)<br>Testosterone (r = -0.08, NS)<br><br>% Body fat:<br>Dihydrotestosterone (r=-0.37, p≤0.01)<br>DHEA (r=-0.28, p≤0.1)<br>DHEAS (r=-0.29, p≤0.1)<br>Androstenedione (r=-0.39, p≤0.05)<br>Androsterone (r=-0.28, p≤0.1)<br>Testosterone (r=-0.06, NS)<br><br>Not reported |
| Pedersen 1995<br>(33)<br>Denmark<br>cross-sectional<br>Follow-up: months | Sample size n= 25<br><br>Premenopausal women with a wide range of BMI: 19.3-48.1; 15 out 25 women are obese<br>Age: 33.2 (SEM= 1.5)<br><br>Pre-menopause<br>(unknown) | Exposures<br>Trunk fat (not reported, DEXA)<br>% Body fat (DEXA)<br>% Trunk fat: DEXA)<br><br>Outcomes<br>SHBG (not reported, not reported)<br>Testosterone (not reported, not reported)<br>free Testosterone (not reported, not reported)<br>Dihydrotestosterone (not reported, not reported)<br>Androstenedione (not reported, not reported)<br>DHEAS (not reported, not reported) | Pearson correlation:<br>Trunk fat: Testosterone (r=-0.02)<br>Trunk fat: SHBG (r=-0.81,p<0.001)<br>Trunk fat: Androstenedione (r=0.02)<br>Trunk fat: DHEAS (r=-0.04)<br>Trunk fat: free Testosterone (r=0.46, p<0.05)<br>Trunk fat:Dihydrotestosterone (r=-0.4, p<0.05)<br>% Body fat: Testosterone (r=-0.09)<br>% Body fat: Androstenedione (r=-0.08)<br>% Body fat: free Testosterone (r=0.36)<br>% Body fat: DHEAS (r=-0.13)<br>% Body fat: SHBG (r=-0.77, p<0.001)<br>% Body fat: Dihydrotestosterone (r=-0.45, p<0.05)<br>% Trunk fat: SHBG (r=-0.85 p<0.001)<br>% Trunk fat: Testosterone (r=0.13)<br>% Trunk fat: Androstenedione (r=0.17)<br>% Trunk fat: DHEAS (r=0.11)<br>% Trunk fat: free Testosterone (r=0.57, p< 0.01)<br>% Trunk fat: Dihydrotestosterone (r=-0.30)<br><br>Not reported |
| De Pergola 1994<br>(88)<br>Italy<br>cross-sectional<br>Follow-up: months | Sample size n= 40<br><br>Premenopausal obese women, BMI >25);<br>Age: 29.5 (SD=8.1) | Exposures<br>IAAT/SAAT thickness (CT)<br>IAAT thickness (mm, CT)<br>SAAT thickness (mm, CT) | Correlation/ Multiple Regression:<br><br>IAAT/SAAT thickness<br>SHBG (r=-0.142, p≥0.05)<br>Androstenedione (r=0.053, p≥0.05)<br>DHEAS (r=-0.139, p≥0.05)<br>Testosterone (r=-0.253, p≥0.05) |

| | | | |
|---|---|---|---|
| | Pre-menopause<br>(Follicular) | Outcomes<br>SHBG (ng/ml, RIA)<br>Androstenedione (ng/ml, RIA)<br>DHEAS (µg/ml, RIA)<br>Testosterone (ng/ml, RIA)<br>free Testosterone (pg/ml, RIA)<br>Estradiol (pg/ml, RIA) | Free Testosterone (r=0.173, p≥0.05)<br>Estradiol (r=0.075, p≥0.05)<br><br>IAAT thickness<br>Testosterone (r=-0.324, p<0.01)<br>Free Testosterone (r=0.286, p≥0.05)<br>DHEAS (r=-0.324, p<0.05)<br>Androstenedione (r=0.01, p≥0.05)<br>Estradiol (r=0.108, p≥0.05)<br>SHBG (r=-0.286, p≥0.05)<br><br>SAAT thickness<br>Testosterone (r=-0.490, p<0.001)<br>Free Testosterone (r=-0.081, p≥0.05)<br>DHEAS (r=-0.258, p≥0.05)<br>Estradiol (r=0.006, p≥0.05)<br>Androstenedione (r=-0.169, p≥0.05)<br>SHBG (r=-0.168, p≥0.05)<br><br>Not reported |
| Zamboni 1994<br>(89)<br>Italy<br>cross-sectional<br>Follow-up: months | Sample size n= 19<br><br>Obese hospitalized premenopausal women, BMI: 36.6 (SD=6.5);<br>Age: 37 (SD=14)<br><br>Pre-menopause<br>(Follicular) | Exposures<br>Total fat area (cm2, CT)<br>SAT (cm2, CT)<br>VAT/SAT (CT)<br>VAT (CT, cm2)<br><br>Outcomes<br>Testosterone (µg/ml, RIA)<br>free Testosterone (pm m-1, RIA) | Simple/ partial correlation:<br>non significant correlation between total fat area and free Testosterone (r=0.23, NS)<br>significant correlation between SAT and Testosterone (r=0.48, p<0.05)<br>non significant correlation between VAT/SAT and free Testosterone (r=-0.14, NS)<br>non significant correlation between VAT and free Testosterone (r=-0.08, NS)<br>non significant correlation between VAT/SAT and Testosterone (r=-0.07, NS)<br>significant correlation between total fat area and Testosterone (r=0.53, p<0.05)<br>non significant correlation between SAT and free Testosterone (r=0.25, NS)<br>non significant correlation between VAT and Testosterone (r=0.23, NS)<br><br>Not reported |
| Armellini 1994<br>(90)<br>Italy<br>cross-sectional<br>Follow-up: months | Sample size n= 36<br><br>Fertile Women; BMI: 27-52;<br>Age: 34 (SD= 11)<br><br>Pre-menopause<br>(Follicular) | Exposures<br>Fat mass (kg, DEXA)<br>VAT/SAT (CT)<br>Total fat area (cm2, CT)<br>VAT (cm2, CT)<br><br>Outcomes<br>Testosterone (nmol/l, RIA)<br>free Testosterone (pmol/ml, RIA)<br>% free testosterone (RIA) | Simple linear correlation:<br>non signicant correlation between Fat mass and Testosterone (r=-0.227, p>0.05)<br>significant correlation between VAT/SAT and Testosterone (r=-0.401, p<0.05)<br>significant correlation between VAT and Testosterone (r=-0.401, p<0.05)<br>non significant correlation between total fat area and Testosterone (r=-0.162, p>0.05)<br>non significant correlation between VAT/SAT and free Testosterone(r=-0.302, p>0.05)<br>non significant correlation between VAT and % free Testosterone (r=-0.202, p>0.05)<br>non significant correlation between VAT and free Testosterone (r=-0.18, p>0.05)<br>non signifcant correaltion between VAT/SAT and % free Testosterone (r=0.006, p>0,05)<br>non significant correlation between total fat area and % free Testosterone (r=0.25, p>0.05)<br>non significant correlation between fat mass and free Testosterone(r=0.126, p>0.05)<br>non significant correlation between fat mass and % free Testosterone (r=0.313, p>0.05<br>non significant correlation between total fat area and free Testosterone (r=-0.125, p>0.05)<br><br>Not reported |
| Cigolini 1996<br>(91) | Sample size n= 87 | Exposures<br>SAAT (cm3, CT) | Pearson correlation:<br>non significant correlation between SAAT and Testosterone (r=0.2, p≥0.05) |

| | | | |
|---|---|---|---|
| Italy<br>cross-sectional<br>Follow-up: months | Clinically healthy women with light to moderte alcohol consumption, BMI: 24.2 (SD=3.8);<br>Age: 38<br><br>Pre-menopause<br>(Unknown) | VAAT (cm3, CT)<br>TAAT (cm3, CT)<br><br>Outcomes<br>Testosterone (ng/ml, H-RIA)<br>free testosterone (pg/ml, solid-phase I-RIA) | significant correlation between VAAT and Testosterone (r=0.48, p=0.01)<br>non signifcant correlation between SAAT and free Testosterone (r=0.4, p≥0.05)<br>significant correlation between VAAT and free Testosterone (r=0.58, p=0.01)<br>non significant correlation between TAAT and Testosterone (r=0.29, p≥0.05)<br>signiciant correlation between TAAT and free Testosterone (r=0.52, p<0.05)<br><br>Adjusted for BMI |
| Yeung 2013<br>(39)<br>USA<br>cohort study<br>Follow-up: months | Sample size n= 239<br><br>Multiethnic premenopausal women, obese (BMI>=30), overweight (BMI 25-30) and normal weight (BMI<25);<br>Age: 27 (SD=8)<br><br>Pre-menopause<br>(Ovulatory) | Exposures<br>Trunk to leg fat ratio (DEXA)<br><br>Outcomes<br>Estradiol (pg/ml, competitive chemiluminescent enzymatic immonoassay)<br>free Estradiol (pg/ml, calculated)<br>Progesterone (ng/ml, competitive chemiluminescent enzymatic immonoassay) | Generalized linear mixed model:<br>percent change of log sex steroids between tertile 1 and 3 of ratio trunk/leg fat, adjusted for age, race, energy intake and physical activity<br><br>trunk/leg fat ratio: Estradiol (-13.5%, ptrend=0.003)<br>trunk/leg fat ratio: Free estradiol (6.4%, ptrend=0.20)<br>trunk/leg fat ratio: Progesterone (-6.1%, ptrend=0.09)<br><br>Adjusted for age, race, energy intake and physical activity |
| Williams 1993<br>(5)<br>USA<br>Non-Randomized Intervention<br>Follow-up: 18 months | Sample size n= 96<br><br>Caucasian premenopausal women, BMI: 22.3 (SD=2.9);<br>Age: 34.3 (SD=3.0)<br><br>Pre-menopause<br>(Follicular) | Exposures<br>% Body fat (DEXA)<br>% Total fat on trunk (DEXA)<br>% Total fat on legs (DEXA)<br>Fat free mass (kg, DEXA)<br><br>Outcomes<br>DHEAS (µmol/l, RIA) | Simple Correlation (unadjusted):<br>% Body fat: DHEAS (r = 0.14, p=0.188)<br>% Total fat on trunk: DHEAS (r = 0.28, p=0.006)<br>% Total fat on legs: DHEAS (r = -0.24, p=0.020)<br>Fat free mass: DHEAS (r = 0.03, p=0.741)<br><br>Partial correlation (adjusted for age, smoking and fasting):<br>% Body fat:DHEAS (r = 0.13, p=0.214)<br>% Total fat on trunk: DHEAS (r = 0.32, p=0.002)<br>% Total fat on legs: DHEAS (r = -0.25, p=0.015)<br>Fat free mass: DHEAS (r = 0.07, p=0.514)<br><br>Simple Correlation - unadjusted, Partial correlation - adjusted for age, smoking and fasting |
| Fan 2019<br>(92)<br>China<br>cross-sectional<br>Follow-up: months | Sample size n= 292,<br>perimenopausal women (80 with obesity and 80 without obesity): 160,<br>premenopausal women (67 with obesity and 65 without obesity): 132<br><br>Pre- and perimenopausal, obese | Exposures<br>VAAT (cm2, CT)<br>SAAT (cm2, CT)<br><br>Outcomes<br>Estradiol (pg/ml, ELISA) | Pearson correlation (obese premenopausal women):<br>VAAT: Estradiol (r=-0.223, p=0.07)<br>SAAT: Estradiol (r=0.156, p=0.209)<br><br>Pearson correlation (obese perimenopausal women):<br>VAAT: Estradiol (r=0.145, p=0.2)<br>SAAT: Estradiol (r=0.136, p=0.228) |

| | | | |
|---|---|---|---|
| | and non-obese women, BMI: > 28 (obese women);<br>Age: 32.21 (SD=5.08) obese, 31 (SD=5.87) non-obese women<br><br>PRE + PERI Mix (Follicular) | | Not reported |
| Wu 2001<br>(93)<br>New Zealand<br>cross-sectional<br>Follow-up: months | Sample size n= 70<br><br>Healthy postmenopausal white women, Weight: 65.5 (SD=10.4); Age: 58.4 (SD=5.7)<br><br>Post-menopause | Exposures<br>Android fat mass (kg, DEXA)<br>Fat mass (kg, DEXA)<br>Andorid/gynoid fat ratio (DEXA)<br>Gynoid fat mass (kg, DEXA)<br><br>Outcomes<br>SHBG (nmol/l, RIA) | Bivariate correlation:<br>Android fat mass:SHBG (r = -0.42; p<0.001)<br>Fat mass:SHBG (r = -0.35; p<0.05)<br>Android/Gynoid fat ratio:SHBG (r = -0.41; p<0.001)<br>Gynoid fat mass:SHBG (r = -0.21, NS)<br><br>Not reported |
| Berman 2001<br>(94)<br>USA<br>cross-sectional<br>Follow-up: months | Sample size n= Caucasian women: 55<br>African American women: 35<br><br>Sedentary, overweight or obese (48% Body fat) Caucasian and African American women, 13 women are using anti-hypertensive medication;<br>Age: 59 (SD = 7) Caucasion, 57 (SD=6) African American women<br><br>Post-menopause | Exposures<br>IAAT (cm2, CT)<br>Fat mass (kg, DEXA)<br>% Body fat (DEXA)<br><br>Outcomes<br>Free Testosterone (pmol/l, RIA)<br>SHBG (nmol/l, IRMA) | Pearson correlation<br>Caucasian women:<br>IAAT:SHBG (r = -0.34; p=0.01)<br><br>African American women:<br>IAAT:SHBG (r = 0.14; NS)<br>Fat mass:SHBG (correlation not reported; NS)<br>% Body fat:SHBG (correlation not reported; NS)<br><br>Fat mass:free Testosterone (correlation not reported; NS)<br>% Body fat:free Testosterone (correlation not reported; NS)<br><br>Not reported |
| Lee 2004<br>(95)<br>USA<br>cross-sectional<br>Follow-up: months | Sample size n= 34<br><br>Multiethnic healthy postmenopausal women, BMI: 26.1 (SEM=0.75);<br>Age: 72 (SEM=1)<br><br>Post-menopause | Exposures<br>% Body fat (DEXA)<br>Central fat (g, DEXA)<br><br>Outcomes<br>Testosterone (ng/ml, chemiluminiscence immunoanalysis)<br>SHBG (nmol/l, chemiluminiscence immunoanalysis)<br>DHEAS (μg/dL, RIA)<br>Androstenedione (ng/ml, RIA)<br>Testosterone free (pg/ml, RIA) | Univariate linear regression:<br>% Body fat:SHBG (r = -0.51; p<0.05)<br>% Body fat:DHEAS (r = 0.09; p≥0.05 NS)<br>% Body fat:Androstenedione (r = 0.02; p≥0.05 NS)<br>% Body fat:Testosterone (r = -0.03; p≥0.05 NS)<br>% Body fat:free Testosterone (r = 0.29; p≥0.05 NS)<br><br>Central fat:SHBG (r = -0.59; p=0.002)<br>Central fat:DHEAS (r = 0.14; p≥0.05 NS)<br>Central fat:Androstenedione (r = 0.06; p≥0.05 NS)<br>Central fat:Testosterone (r = -0.02; p≥0.05 NS)<br>Central fat:free Testosterone (r = 0.29; p≥0.05 NS)<br><br>Not reported |

| | | | |
|---|---|---|---|
| Mahabir 2006<br>(17)<br>USA<br>cross-sectional<br>Follow-up: months | Sample size n= 51<br><br>Multiethnic postmenopausal women, BMI: 27.8 (range=17.7-42.5);<br>Age: 59.7 (range=49.2-78.8)<br><br>Post-menopause | Exposures<br>% Body fat (DEXA)<br>Central fat (g, DEXA)<br>Peripheral fat (g, DEXA)<br><br>Outcomes<br><br>Estradiol (ng/dl, RIA)<br>bioavailable Estradiol (ng/dl, RIA)<br>Estrone (ng/dl, RIA)<br>Estrone sulfate (ng/dl, RIA)<br>Testosterone (ng/dl, RIA)<br>Androstenedione (ng/dl, RIA)<br>DHEA (ng/dl, RIA)<br>DHEAS (µg/dl, RIA)<br>Progesterone (ng/dl, RIA)<br>Androstendiol (ng/dl, RIA)<br>SHBG (nmol/l, RIA) | Linear regression (beta-coefficients) (adjusted for age, race, family history of breast cancer, parity, menarche <12 years, and additionaly for BMI after coma)<br>Higher level of % Body fat with<br>Estradiol (b=5.8, p < 0.05), (b=13.4, NS)<br>Bioavailable Estradiol (b=7.9, p < 0.005), (b=17.1,p < 0.05)<br>Estrone (b=2.9, p<0.0005), (b=3.4, NS)<br>Estrone sulfate (b=4.2, p<0.0005), (b=7.9, p<0.005)<br>Testosterone (b=0.6, NS), (b=3.8, NS)<br>Androstenedione (b=0.4, NS), (b=1, NS)<br>DHEA (b=-0.1, NS), (b=-0.5, NS)<br>DHEAS (b=-0.2, NS), (b=3.6, NS)<br>Progesterone (b=-0.4, NS), (b=2.9, NS)<br>Androstendiol (b=-0.2, NS), (b=0.8,NS)<br>SHBG (b=-3.4, p<0.0005), (b=-5.1, p < 0.05)<br><br>Higher level of Central fat with<br>Estradiol (b=8.6, p < 0.05), (b=22.7,p < 0.05)<br>Bioavailable Estradiol (b=12.2, p < 0.005), (b=26.3,p < 0.05)<br>Estrone (b=4, p<0.005), (b=6.8,p < 0.05)<br>Estrone sulfate (b=6.8, p<0.0005), (b=9.0,p < 0.005)<br>Testosterone (b=0.2, NS), (b=10.8,p < 0.05)<br>Androstenedione (b=0.3, NS), (b=3.9,NS)<br>DHEA (b=-0.6, NS), (b=2.4, NS)<br>DHEAS (b=0.003, NS), (b=3.8, NS)<br>Progesterone (b=-0.3, NS), (b=3.0, NS)<br>Androstendiol (b=-0.6, NS), (b=2.8, NS)<br>SHBG (b=-5.8, p<0.0005), (b=-3.2, NS)<br><br>Higher level of Peripheral fat with<br>Estradiol (b=9.7, p < 0.05), (b=11.2, NS)<br>Bioavailable Estradiol (b=12.7, p < 0.0005), (b=17.2, NS)<br>Estrone (b=4.5, p<0.0005), (b=1.4, NS)<br>Estrone sulfate (b=6.1, p<0.0005), (b=11.3, p<0.0005)<br>Testosterone (b=1.3, NS), (b=3.7, NS)<br>Androstenedione (b=0.6, NS), (b=1.8, NS)<br>DHEA (b=-0.6, NS), (b=2.4, NS)<br>DHEAS (b=-0.3, NS), (b=6.3, NS)<br>Progesterone (b=0.4, NS), (b=-1.7, NS)<br>Androstendiol (b=-0.4, NS), (b=2.6, NS)<br>SHBG (b=-4.9, p<0.0005), (b=-6.9, p<0.05)<br><br>adjusted for age, race, family history of breast cancer, parity, menarche <12 years<br><br>+BMI in second model |
| Baglietto 2009<br>(18)<br>Australia<br>cross-sectional<br>Follow-up: months | Sample size n= 770<br><br>Naturally postmenopausal women from the Melbourne Collaborative Cohort Study, BMI: 27.2 (SD=4.6); | Exposures<br>Fat mass (kg, BIA)<br><br><br>Outcomes | Linear Regression (beta-coefficients per 10 kg fat mass):<br>Adjusted for laboratory batches, age and country of birth:<br>Fat mass:Estradiol (b=7.8 [8 (3.7;11.9])<br>Fat mass:free Estradiol (b=18.4 [13.6;23.1])<br>Fat mass:Testosterone (b=4.4 [-1.6;10.5]) |

| | | | |
|---|---|---|---|
| | Age: 61 (range=46-70)<br><br><br>Post-menopause | Estradiol (pmol/l, electrochemiluminescence immunoassay)<br>free Estradiol (pmol/l, electrochemiluminescence immunoassay)<br>Testosterone (nmol/l, electrochemiluminescence immunoassay)<br>DHEAS (competitive immunoassay, umol/l)<br>Androstenedione (nmol/l, RIA)<br>SHBG (nmol/l, immunometric assay)<br>Estrone sulfate (nmol/l, RIA) | Fat mass:DHEAS (b=-2.1 [-8.0;3.7])<br>Fat mass:Androstendione (b=3 [-1.9;7.9])<br>Fat mass:SHBG (b=-17.4 [-20.1; -14.7])<br>Fat mass:Estrone sulfate (b = 10.1 [5.7; 14.4])<br><br>Adjusted for laboratory batches, age, country of birth, age at menarche, duration of lactation, parity, oral contraceptive use, HRT use, age at menopause, physical activity, alcohol consumption, total energy intake, smoking and level of education:<br>Fat mass:Estradiol (b = 9.1 [4.9; 13.4])<br>Fat mass:free Estradiol (b = 18.8 [13.8; 23.7])<br>Fat mass:Testosterone (b = 5 [-0.2;12.2])<br>Fat mass:DHEAS (b = -1.4 [-7.5;4.6])<br>Fat mass:Androstenedione (b = 3.6 [-1.5; 8.7])<br>Fat mass:SHBG (b = -16.2 [-19.0; -13.4])<br>Fat mass:Estrone sulfate (b = 10 [5.5;14.4])<br><br>1) adjusted for laboratory batches, age and country of birth;<br>2) adjusted for laboratory batches, age, country of birth, age at menarche, duration of lactation, parity, oral contraceptive use, HRT use, age at menopause, physical activity, alcohol consumption, total energy intake, smoking and level of education |
| Casson 2010<br>(31)<br>USA<br>cross-sectional<br>Follow-up: months | Sample size n= 29 (expecpt correlations for Dihydrotestosterone (n = 24) and Androsterone glucoronide (n = 26))<br><br>Postmenopausal normal weight to nonobese women, BMI: ≤ 30;<br>Age = 60.7 (SE = 1)<br><br>Post-menopause | Exposures<br>VAAT (cm2 , CT), TAAT (cm2 , CT), SAAT (cm2 , CT), VAT/SAT<br>Fat mass (kg, DEXA), % Body fat (DEXA)<br><br>Outcomes<br>DHEAS (µg/dl, chemiluminiscence immunoanalysis), SHBG (nmol/l, chemiluminiscence immunoanalysis)<br>Testosterone (ng/d, RIA), Androstenedione (ng/dl, RIA), DHEA (ng/ml, RIA)<br>Dihydrotestosterone (ng/dl, HPLC-MS), Androsterone glucoronide (ng/ml, HPLC-MS)<br>Free Testosterone (pg/ml) was calculated by a valited algorithm | Pearson correlation:<br>VAAT:DHEAS (r = 0.003)<br>VAAT:Testosterone (r = -0.11)<br>VAAT:free Testosterone (r = -0.115)<br>TAAT:Androstenedione (r = 0.027)<br>TAAT:DHEAS (r = 0.188)<br>TAAT:free Testosterone (r = -0.084)<br>TAAT:Testosterone (r = -0.09)<br>Fat mass:free Testosterone (r = -0.028)<br>VAAT:Androstenedione (r = -0.19)<br>Fat mass:Testosterone (r = -0.332, p<0.10)<br>% Body fat:Androstenedione (r = -0.202)<br>% Body fat:DHEAS (r = 0.081)<br>% Body fat:free Testosterone (r = -0.084)<br>% Body fat:Testosterone (r = -0.375, p=0.045)<br>VAAT:SHBG (r = 0.128)<br>VAAT:Dihydrotestosterone (r = 0.34)<br>TAAT:Dihydrotestosterone (r = 0.152)<br>TAAT:SHBG (r = -0.007)<br>Fat mass:SHBG (r = -0.58, p<0.01)<br>Fat mass:Dihydrotestosterone (r = -0.293)<br>% Body fat:SHBG (r = -0.536, p<0.01)<br>% Body fat:Dihydrotestosterone (r = -0.175)<br>SAAT:free Testosterone (r = -0.034)<br>SAAT:Dihydrotestosterone (r = 0.04)<br>VAT/SAT:free Testosterone (r = -0.03)<br>VAT/SAT:DHEA (r = -0.246)<br>SAAT:DHEA (r = 0.251)<br>VAT/SAT:Androstenedione (r = -0.311) |

| | | | |
|---|---|---|---|
| | | | SAAT:Androstenedione (r = 0.132)<br>VAT/SAT:Dihydrotestosterone (r = 0.224)<br>VAT/SAT:Testosterone (r = 0.044)<br>SAAT:Testosterone (r = -0.065)<br>% Body fat:Androsterone glucoronide (r = 0.033)<br>Fat mass:Androsterone glucoronide (r = -0.012)<br>TAAT: Androsterone glucoronide (r = -0.027)<br>VAAT: Androsterone glucoronide (r = -0.111)<br>SAAT: Androsterone glucoronide (r = 0.023)<br>VAT/SAT: Androsterone glucoronide (r = -0.134)<br><br>Not reported |
| Rariy 2011<br>(19)<br>USA<br>cross-sectional<br>Follow-up: months | Sample size n= 232<br><br>Multiethnic postmenopausal women, BMI: 27.2 (SD=5);<br>Age: 75.6 (SD=4.6)<br><br>Post-menopause | Exposures<br>Fat mass (kg, DEXA), % Body fat (DEXA)<br><br>Outcomes<br>Testosterone (ng/dl, RIA)<br>Free Testosterone (pg/ml, measured by sensitive equilibrium dialysis assay) | Multivariable linear regression: Crude Î² coefficient<br>Fat mass:Testosterone (b = 0.61, p = 0.39)<br>% Body fat:Testosterone (b =0.31, p=0.54)<br><br>Fat mass:free Testosterone (b =2.68, p=0.002)<br>% Body fat:free Testosterone (b=1.71, p=0.005)<br><br>Multivariable linear regression: Adjusted Î² coefficient (adjusted for age, race, weight, estrogen use)<br>Fat mass:Testosterone (b=0.23, p=0.73)<br>% Body fat:Testosterone (b=0.05, p=0.91)<br><br>Fat mass:free Testosterone (b = 2.11, p=0.007)<br>% Body fat:free Testosterone (b=1.39, p=0.02)<br><br>1) unadjusted<br>2) adjusted<br>3) adjusted + E2 adjusted |
| Gourlay 2012<br>(96)<br>USA<br>cross-sectional<br>Follow-up: months | Sample size n= 94<br><br>Multiethnic community dwelling postmenopausal women, BMI: 29.9 (SD=7.7);<br>Age: 57.6 (SD=3.6)<br><br>Post-menopause | Exposures<br>Fat mass (kg, DEXA)<br><br>Outcomes<br>free Estradiol (ng/dl, calculated)<br>SHBG (nmol/l, biochemical analyses)<br>free Testosterone (ng/dl, biochemical analyses) | Multiple linear regression (beta coefficients):<br>Fat mass:free Estradiol (b=2.586, p≥0.5)<br>Fat mass:SHBG (b = -0.131, p≥0.05)<br>Fat mass:free Testosterone ( b = -0.410, p≥0.05)<br><br>Spearman correlation:<br>Fat mass:free Estradiol (r=0.51, p<0.001)<br>Fat mass:SHBG (r = -0.48, p<0.001)<br>Fat mass:free Testosterone (r = 0.24, p<0.02)<br><br>Adjusted for age, race, years<br>since menopause, FSH, bio E2, bio T, LH, PTH, SHBG and urine NTx. |
| Cao 2013<br>(97)<br>China<br>cross-sectional<br>Follow-up: months | Sample size n= 212,<br>early postmenopausal women: 105,<br>late postmenopausal women: 107<br><br>Women who were recruited from | Exposures<br>Trunk to leg fat ratio (DEXA)<br>% Gynoid fat (DEXA)<br>% Android fat (DEXA) | Pearson correlation<br>Early postmenopausal women:<br>Trunk to leg fat ratio:DHEAS (r=0.03, p=0.975)<br>Trunk to leg fat ratio: Testosterone (r=0.76, p=0.474)<br>Trunk to leg fat ratio: free Testosterone (r=0.339, p=0.001)<br>Trunk to leg fat ratio: SHBG (r=-0.221, p=0.034) |

| | | | |
|---|---|---|---|
| | the community around the Gynecology & Obstetrics Hospital of Fudan University between 2009 and 2011, both groups subdivided into normal weight (BMI<24) and overweight and obese (BMI >=24) group;<br>Age: 46 to 85<br><br>Post-menopause | Outcomes<br>DHEAS (µg/dl, chemiluminescence immunoanalysis)<br>Testosterone (ng/ml, chemiluminescence immunoanalysis)<br>free Testosterone (pmol/l, calculated)<br>SHBG (nmol/l, chemiluminescence immunoanalysis) | % Gynoid fat: DHEAS (r=-0.017, p=0.874)<br>% Gynoid fat: Testosterone (r=0.017, p=0.874)<br>% Gynoid fat: free Testosterone (r=-0.231, p=0.027)<br>% Gynoid fat: SHBG (r=0.270, p=0.009)<br><br>% Android fat: DHEAS (r=0.034, p=0.749)<br>% Android fat: Testosterone (r=-0.026, p=0.805)<br>% Android fat: free Testosterone (r=0.227, p=0.030)<br>% Android fat: SHBG (r=-0.297, p=0.007)<br><br>Late postmenopausal women:<br>Trunk to leg fat ratio: DHEAS (r = 0.297, p = 0.007)<br>Trunk to leg fat ratio: Testosterone (r = -0.055, p = 0.68)<br>Trunk to leg fat ratio: free Testosterone (r = -0.05, p = 0.71)<br>Trunk to leg fat ratio: SHBG (r = 0.006, p = 0.964)<br><br>% Android fat: DHEAS (r = 0.282, p = 0.032)<br>% Android fat: Testosterone (r = 0.119, p = 0.373)<br>% Android fat: free Testosterone (r = 0.128, p = 0.34)<br>% Android fat: SHBG (r = -0.064, p = 0.631)<br><br>% Gynoid fat: DHEAS (r = -0.277, p = 0.035)<br>% Gynoid fat: Testosterone (r = -0.109, p = 0.413)<br>% Gynoid fat: free Testosterone (r = -0.122, p = 0.362)<br>% Gynoid fat: SHBG (r = 0.077, p = 0.564)<br><br>adjusted for age and BMI |
| Aguirre 2014<br>(15)<br>USA<br>cross-sectional<br>Follow-up: months | Sample size n= 92<br><br>Baseline characteristics of sedentary, frail, elderly obese women participating in a lifestyle therapy with diet with or without exercise, BMI: 36.6 (SD=25.7) men and women;<br>Age: 69.5 (SD=4.2) men and women<br><br>Post-menopause | Exposures<br>% Body fat (DEXA)<br><br>Outcomes<br>Free Estradiol index (pmol/nmol, RIA) | Women only:<br>ANOVA: free estradiol index (FEI) decreases across tertile of % body fat<br>FEI: 2.1 (1.7, 2.5) 1.2 (0.8, 1.5) 1.4 (0.9, 2.0) p=0.02<br><br>Not reported |
| Bann 2015<br>(22)<br>UK<br>cohort<br>Follow-up: months | Sample size n= 560<br><br>Study members of the MRC National Survey of Health and Development (NSHD), BMI: 27.5 (SD=5.0);<br>Age: 60-64 (women) | Exposures<br>Fat mass Index (kg/m), DEXA<br>Android/gynoid fat ratio (DEXA)<br><br>Outcomes | Linear Regression: (mean percentage differences in fat mass and android/gynoid fat ratio per 1 standard deviation in serum testosterone)<br><br>Values for age = 53;<br>Android/Gynoid fat ratio:Testosterone (3.29, p=0.02)<br>Android/Gynoid fat ratio:SHBG (-8.86, p<0.001)<br>Android/Gynoid fat ratio:free Testosterone (8.36, p<0.001) |

| | | | |
|---|---|---|---|
| | PRE + POST Mix | SHBG (nmol/l, immunoassay)<br>Testosterone (nmol/l, LC-MS/MS)<br>free Testosterone (nmol/l, calculated) | Fat mass index:Testosterone (4.57, p<0.01)<br>Fat mass index:SHBG (-9.11, p<0.001)<br>Fat mass index:free Testosterone (9.51, p<0.001)<br><br>Change Testosterone between age 53 and 60-64;<br>Android/Gynoid fat ratio:Change Testosterone (0.72, p=0.67)<br>Android/Gynoid fat ratio:Change SHBG (-13.76, p<0.001)<br>Android/Gynoid fat ratio:Change free Testosterone (4.2, p<0.01)<br><br>Fat mass index:Change Testosterone (0.47, p=0.8)<br>Fat mass index:Change SHBG (-13.21, p<0.001)<br>Fat mass index:Change free Testosterone (3.63, p=0.03)<br><br>For change between 53 and 60–64 years: analyses adjusted for hormone concentration at 53 years |
| Colleluori 2018<br>(98)<br>USA<br>cross-sectional<br>Follow-up: months | Sample size n= 252<br><br>Community-dwelling postmenopausal women, BMI: 29.8 (SD=6.5);<br>Age: 66.1 (SD=7.6)<br><br>Post-menopause | Exposures<br>Fat mass and trunk fat (kg, DEXA)<br>% Fat mass and trunk fat (DEXA)<br><br>Outcomes<br>Estradiol (pg/ml, RIA) | Anova of trunk fat mass across Estradiol category adjusted for age:<br>Trunk Fat mass p<0.01<br>Trunk fat mass percentage p=0.16<br><br>Correlation (adjustment not reported):<br>Fat mass* Estradiol (r=0.07, p=0.34), but U-shape<br>Fat mass percentage * Estradiol (r=-0.04, p=0.56), but U-shape<br><br>Not reported |
| Ofori 2019<br>(99)<br>Switzerland<br>cross-sectional<br>Follow-up: months | Sample size n= 35 (for % Body fat and fat mass)<br>34 (for VAAT, SAAT, TAAT, Thigh SAT, Thigh fat, Thigh intramuscular fat)<br><br>Postmenopausal weight stable caucasian women, BMI: 25.5 (SEM=1.0);<br>Age: 66.6 (SD=0.8)<br><br>Post-menopause | Exposures<br>VAAT (L, MRI)<br>TAAT (L, MRI)<br>SAAT (L, MRI)<br>Fat mass (kg, DEXA)<br>% Body fat (DEXA)<br>Thigh intermuscular fat (L, MRI)<br>Thigh fat (L, MRI)<br>Thigh SAT (L, MRI)<br><br>Outcomes<br>Testosterone (nmol/l, LC/MS) | Spearman correlation:<br>non significant correlation between VAAT and Testosterone (r=0.25, p=0.154)<br>significant correlation between TAAT and Testosterone (r=0.34, p=0.046)<br>non significant correlation between SAAT and Testosterone (r=0.37, p=0.034)<br>significant correlation between Fat mass and Testosterone (r=0.42, p=0.012)<br>significant correlation between % Body fat and Testosterone (r=0.38, P=0.025)<br>significant correlation between thigh intermusculuar fat and Testosterone (r=0.35, P=0.039)<br>significant correlation between thigh fat and Testosterone (r=0.49, P=0.003)<br>significant correlation between thigh SAT and Testosterone (r=0.50, P=0.003)<br><br>Not reported |
| Jensen 1985<br>(100)<br>Denmark<br>cross-sectional<br>Follow-up: months | Sample size n= 291,<br>early postmenopausal women: 144<br>late postmenopausal women: 147<br><br>Two large groups of postmenopausal women. In the younger group, women were between 46 and 56 (mea52.4, SD=2.3) years of age and in the | Exposures<br>Fat mass (kg, using formula from Boddy et al.)<br><br>Outcomes<br>Estradiol (pmol/l, RIA)<br>Estrone (pmol/l, RIA) | Correlation early postmenopausal women:<br>significant correlation between fat mass and Estradiol (r=0.31, p<0.001)<br>significant correlation between fat mass and Estrone (r=0.183, p<0.05)<br>Correlation late postmenopausal women:<br>significant correlation between fat mass and Estradiol (r=0.296, p<0.001)<br>non significant correlation between fat mass and Estrone (r=0.14, p>=0.05)<br><br>Not reported |

| | | | |
|---|---|---|---|
| | older group they were 70 (mea70, SD=0) years, Weight: younger=63.6 (SD=11.9), older group=64.2 (SD=9.7)<br><br>Post-menopause | | |
| Suzuki 1995<br>(101)<br>Japan<br>cross-sectional<br>Follow-up: months | Sample size n= 30<br><br>Healthy, postmenopausal women, BMI/weight: not reported;<br>Age: 65.0 (SD=9.3)<br><br>Post-menopause | Exposures<br>% Body fat (DEXA)<br><br>Outcomes<br>Estrone (pg/ml, RIA) | Simple correlation:<br>significant correaltion between % Body fat and Estrone (r=0.391, p<0.05)<br>Slope B=5.72 + 1.74x (positively related, S)<br><br>Not reported |
| Kleerekoper 1994<br>(102)<br>USA<br>cross-sectional<br>Follow-up: months | Sample size n= 278,<br>black women: 77,<br>white women: 201<br><br>Black and white postmenopausal women, BMI: 31.7 (SD=6.0) black, 29.2 (SD=5.9) white women;<br>Age: 66.1 (SD=4.9) black, 66.8 (SD=5.0) white women<br><br>Post-menopause | Exposures<br>Fat mass (kg, DEXA)<br><br>Outcomes<br>Estrone (pmol/l, RIA) | Pearson correlation:<br>significant correlation between fat mass and estrone in white women (r=0.323, p<0.001)<br>non significant correlation between fat mass and estrone in black women (r=0.182, p=0.108)<br><br>Not reported |
| Svendsen 1993<br>(103)<br>Denmark<br>cross-sectional<br>Follow-up: months | Sample size n= 121<br><br>Overweight postmenopausal women, BMI >= 25;<br>Age: 53.8 (SD=2.5)<br><br>Post-menopause | Exposures<br>% Abdominal fat (DEXA)<br>Fat mass (kg, DEXA)<br><br>Outcomes<br>SHBG (nmol/l, IRMA) | Pearson correlation:<br>% Abdominal fat:SHBG (r = -0.30, p<0.01)<br>Fat mass:SHBG (r = -0.06, NS)<br><br>Not reported |
| Abbasi 1998<br>(104)<br>USA<br>cross-sectional<br>Follow-up: months | Sample size n= 118 (women only)<br><br>White women aged 60 or more, BMI: 25.1 (SD=3.15);<br>Age: 66.6 (SD=4.8)<br><br>Post-menopause | Exposures<br>Fat mass (g, DEXA)<br>% Body fat (DEXA)<br><br>Outcomes<br>DHEAS (µg/dl, RIA) | Correlation<br>non significant correlation between fat mass and DHEAS (r=0.11, p=0.241)<br>non significant correlation between % Body fat and DHEAS (r=0.09, p=0.343)<br><br>Not reported |
| Paolillo 2014<br>(105)<br>Brazil<br>cross-sectional<br>Follow-up: months | Sample size n= 45,<br>Android goup: 32,<br>Gynoid group: 13<br><br>Caucasian postemenopausal, healthy, brazilian women, BMI: 26±4 (gynoid), 31±5 (android);<br>Age: 55 (SD=2) android, 56 (SD=2) gynoid groud | Exposures<br>% Body fat (BIA)<br><br>Outcomes<br>Estradiol (pg/ml, not reported) | Pearson correlation:<br>Significant correlation between % Body fat and Estradiol (r=0.41, p<0.01)<br><br>Not reported |

| | | | |
|---|---|---|---|
| | Post-menopause | | |
| Turcato E 1997<br>(4)<br>Italy<br>Non-Randomized<br>Intervention<br>Follow-up: 1 months | Sample size n= 26 (premenopausal women, 25 for adjusted VAT)<br>15 (postmenopausal women, 13 for adjusted VAT)<br><br>Obese premenopausal and postmenopausal women, BMI: 38.3 (SD = 6.4) premenopausal, 35.0 (SD = 4.9) postmenopausal women;<br>Age: 33.7 (SD = 10.2) premenopausal, 59.9 (SD = 5.9) postmenopausal women<br><br>STRAT PRE POST<br>(follicular) | Exposures<br>Body fat (kg, CT)<br>SAT (cm2, CT)<br>VAT (cm2, CT)<br><br>Outcomes<br>SHBG (unit:ng/ml^-1, RIA)<br>17-β-oestradiol (ng/ml^-1, RIA)<br>Total Testosterone (ng/ml^-1, RIA)<br>Free Testosterone (pg/ml^-1, RIA)<br>Androstenedione (ng/ml^-1, RIA) | Simple correlation for Premenopausal women:<br>Body fat: SHBG (r = -0.48, p<0.05)<br>Body fat: 17-β-oestradiol (r = 0.10, NS)<br>Body fat: Total Testosterone (r = -0.01, NS)<br>Body fat: Free Testosterone (r = 0.23, NS)<br>Body fat: Androstenedione (r = -0.05, NS)<br>SAT: SHBG (r = -0.41, p<0.05)<br>SAT: 17-β-oestradiol (r = 0.03, NS)<br>SAT: Total Testosterone (r = 0.14, NS)<br>SAT: Free Testosterone (r = 0.27, NS)<br>SAT: Androstenedione (r = 0.06, NS)<br>VAT: SHBG (r = -0.35, NS)<br>VAT: 17-β-oestradiol (r = 0.21, NS)<br>VAT: Total Testosterone (r = -0.42, p<0.05)<br>VAT: Free Testosterone (r = -0.04, NS)<br>VAT: Androstenedione (r = -0.31, NS)<br><br>Partial correlation for premenopausal women (adjusted for age):<br>VAT: SHBG (r = -0.59, p<0.01)<br>VAT: 17-β-oestradiol (r = 0.17, NS)<br>VAT: Total Testosterone (r = -0.11, NS)<br>VAT: Free Testosterone (r = 0.24, NS)<br>VAT: Androstenedione (r = -0.51, p<0.01)<br><br>Simple correlation for postmenopausal women:<br>Body fat: SHBG (r = -0.35, NS)<br>Body fat: Total Testosterone (r = -0.10, NS)<br>Body fat: Free Testosterone (r = 0.09, NS)<br>Body fat: Androstenedione (r = -0.12, NS)<br>SAT: SHBG (r = -0.01, NS)<br>SAT: Total Testosterone (r = -0.13, NS)<br>SAT: Free Testosterone (r = -0.01, NS)<br>SAT: Androstenedione (r = -0.29, NS)<br>VAT: SHBG (r = -0.62, p<0.01)<br>VAT: Total Testosterone (r = 0.15, NS)<br>VAT: Free Testosterone (r = -0.26, NS)<br>VAT: Androstenedione (r = 0.24, NS)<br><br>Partial correlation for postmenopausal women (adjusted for age):<br>VAT: SHBG (r = -0.70, p<0.001)<br>VAT: Total Testosterone (r = -0.03, NS)<br>VAT: Free Testosterone (r = 0.21, NS)<br>VAT: Androstenedione (r = 0.24, NS) |

| | | | |
|---|---|---|---|
| | | | Partial correlations between visceral AT and sex hormones are adjusted for age |
| Hajamor 2003<br>(106)<br>Canada<br>cross-sectional<br>Follow-up: months | Sample size n= Premenopausal women: 167<br>Postmenopausal women: 44<br><br>Non-smoking premenopausal women and postmenopausal women, BMI: 16.8 to 49.9;<br>Age: 34 (SD = 9.3) premenopausal, 56.5 (SD = 5.7) postmenopausal women<br><br>PRE + POST Mix | Exposures<br>IAAT (cm2, CT)<br>TAAT (cm2, CT)<br>SAAT (cm2, CT)<br>Fat mass (kg, calculated by Siri equation)<br><br>Outcomes<br>SHBG (nmol/l, RIA) | Spearman correlation for premenopausal women:<br>IAAT:SHBG (r = -0.38; p<0.005)<br>Fat mass:SHBG (r = -0.36; p<0.005)<br>TAAT:SHBG (r = -0.41; p<0.005)<br>SAAT:SHBG (r = -0.41; p<0.005)<br>Spearman correlation for postmenopausal women:<br>TAAT:SHBG (r = -0.23; p<0.005)<br>IAAT:SHBG (r = -0.15; p<0.05)<br>Fat mass:SHBG (r = -0.2; p<0.005)<br>SAAT:SHBG (r = -0.26; p<0.005)<br><br>Not reported |
| Philips 2008<br>(107)<br>USA<br>cross-sectional<br>Follow-up: months | Sample size n= 78,<br>premenopausal women: 58,<br>postmenopausal women: 20<br><br>Adult females recruited from a multiethnic community through advertisements, premenopausal and postmenopausal women, BMI: 24.1 (SEM=0.6) premenopausal, 27.9 (SEM=1.0) postmenopausal women;<br>Age: 32.9 (SEM=1.2) premenopausal, 61.4 (SEM=2.4) postmenopausal women<br><br>STRAT PRE POST | Exposures<br>VAT (kg, MRI)<br>SAT (kg, MRI)<br>Fat mass (kg, MRI)<br><br>Outcomes<br>Estrone<br>Estradiol (pg/ml, RIA)<br>Testosterone (ng/ml, RIA)<br>free Testosterone (pg/ml, RIA)<br>DHEAS (µg/dl, RIA)<br>SHBG (nmol/l, RIA) | Pearson Corrleation for premenopausal women:<br>VAT<br>Testosterone (r=0.11, p>0.05)<br>Free Testosterone (r=0.4, p≤0.005)<br>DHEAS (r=-0.04, p>.05)<br>SHBG (r=-0.38, p≤0.005)<br><br>SAT<br>Testosterone (r=0.22, p>0.05)<br>Free Testosterone (r=0.29, p≤0.05)<br>SHBG (r=-0.11, p>0.05)<br>DHEAS (r=-0.13, p>0.05)<br><br>Fat mass<br>Testosterone (r=0.22, p>0.05)<br>Free Testosterone(r=0.30, p≤0.05)<br>DHEAS (r=-0.13, p>0.05)<br>SHBG(r=-0.13, p>0.05)<br><br>Pearson correlation for postmenopausal women:<br>VAT<br>Testosterone (r=0.42, p>0.05)<br>Free Testosterone (r=0.51, p≤0.05)<br>SHBG (r=-0.39, p>0.05)<br>DHEAS (r=-0.06, p>0.05)<br>Estradiol (r=0, p>0.05)<br>Estrone (r=-0.09, p>0.05)<br><br>SAT<br>Estradiol (r=-0.08, p>0.05)<br>Estrone (r=-0.04, p>0.05)<br>Testosterone (r=0.41, p>0.05)<br>Free Testosterone (r=0.39, p>0.05) |

| | | | |
|---|---|---|---|
| | | | SHBG (r=-0.08, p>0.05)<br>DHEAS (r=-0.28, p>0.05)<br><br>Fat mass<br>Estradiol (r=-0.07, p>0.05)<br>Estrone (r=-0.05, p>0.05)<br>Testosterone (r=0.43, p>0.05)<br>Free Testosterone (r=0.42, p>0.05)<br>DHEAS (r=-0.26, p>0.05)<br>SHBG (r=-0.13, p>0.05)<br><br>age-adjusted |
| Veldhuis 2014<br>(21)<br>USA<br>cross-sectional<br>Follow-up: months | Sample size n= 120, premenopausal women: 60, postmenopausal women:60<br><br>Healthy nonpregnant pre- and postmenopausal women, BMI: 26 (SD=4.8);<br>Age: 49 (SD=17)<br><br>STRAT PRE POST | Exposures<br>VAAT (cm2, CT)<br>Fat mass (cm2, CT)<br><br>Outcomes<br>SHBG (nmol/l, chemiluminescence immunoanalysis) | Multivariate regression --<br>Premenopausal women:<br>VAAT and SHBG (b=-0.65, p<0.001)<br>Fat mass and SHBG (NS)<br><br>Postmenopausal women:<br>VAAT and SHBG (b=-0.048, p=0.002)<br>Fat mass and SHBG (NS)<br><br>Adjusted for age, BMI, TT, abumine, glucose, insulin, E1, E2, DHT, systolic and diastolic BP, cholesterol, triglycerides, HDL. |
| Seyfart 2018<br>(24)<br>Germany<br>cross-sectional<br>Follow-up: months | Sample size n= 520<br><br>German citizens in northeastern Germany, BMI: median=25.9 (range=23.1; 29.3);<br>Age: median=50 (range=20-97)<br><br>PRE + POST Mix | Exposures<br>SAAT (L, MRI)<br>VAAT (L, MRI)<br><br>Outcomes<br>Testosterone (nmol/l, LC/MS)<br>SHBG (nmol/l, chemiluminescence immunoanalysis)<br>Androstenedione (nmol/l, LC/MS)<br>DHEAS (mg/l, chemiluminescence immunoanalysis) | Linear regression (beta coefficients):<br>SAAT * Testosterone (b=0.549, p=0.3)<br>SAAT * SHBG (beta=-0.121, p<0.01)<br>SAAT * Androstenedione (b=-0.185, p=0.34)<br>SAAT * DHEAS (b=0.419, p=0.18)<br><br>VAAT * Testosterone (b=0.247, p=0.13)<br>VAAT * SHBG (b=-0.064, p<0.01)<br>VAAT * Androstenedione (b=0.056, p=0.28)<br>VAAT * DHEAS (b=0.142, p=0.17)<br><br>Linear regression model - adjusted for age, sex, smoking, physical activity, diabetes, hypertension, and cholesterol |
| Denti 1997<br>(108)<br>Italy<br>cross-sectional<br>Follow-up: months | Sample size n= 155<br><br>Healthy pre- and postmenopausal women, BMI<30;<br>Age: 18-92<br><br>PRE + POST Mix | Exposures<br>% Body fat (BIA)<br><br>Outcomes<br>DHEAS (µmol/l, RIA) | Unadjusted correlation:<br>% Body fat:DHEAS (r=-0.48, p=0.000)<br>Age-adjusted correlation:<br>% Body fat:DHEAS (r=-0.16, not significant)<br>Correlation adjusted for age and insulin:<br>% Body fat:DHEAS (r = 0.08, not significant)<br><br>Bivariate and Partial Correlation Analysis - unadjusted, adjusted for age and adjusted for age and insulin |

| | | | |
|---|---|---|---|
| Kunesová 2002<br>(109)<br>Czech Republik<br>cross-sectional<br>Follow-up: months | Sample size n= 94<br><br>Females with weight ranging from overweight to severe obesity,<br>Age= 44.2 [21-67]<br>BMI: 37.13 (SD=5.72);<br>Age: 44.2 (SD=11.2)<br><br>Not reported | Exposures<br>SAAT (cm2, CT)<br>IAAT (cm2, CT)<br><br>Outcomes<br>DHEA (µg/l, RIA)<br>DHEAS (µmol/l, RIA)<br>Testosterone (nmol/l, RIA) | Correlations:<br>SAAT*DHEA (r=0.03, NS)<br>SAAT*DHEAS (r=0.1, NS)<br>SAAT*Testosterone (r=0.27, p=0.01)<br><br>IAAT*DHEA (r=-0.3, p<0.01)<br>IAAT*DHEAS (r=-0.34, p<0.005)<br>IAAT* Testosterone (r=-0.17, NS)<br><br>Multiple Regression adjusted for c-peptide:<br>higher levels of IAAT is significantly associated with DHEA (b=-8.17, p<0.001)<br><br>Not reported |
| Guo 2023<br>(40)<br>China<br>cross-sectional<br>Follow-up: months | Sample size n= 250<br><br>Overweight or obese postmenopausal women who undergo bariatric surgery<br>BMI: mean 38.5 (SD=5.2)<br>Age: mean=32.5 (SD=7.5)<br><br>Pre-menopause (follicular) | Exposures<br>VAT (cm2, CT)<br>TAAT (cm2, CT)<br>SAT (cm2, CT)<br><br>Outcomes<br>Progesterone (nmol/l, ECL)<br>Estradiol (nmol/l, ECL)<br>Testosterone (nmol/l, ECL) | Linear regression adjusted for age and BMI:<br>Progesterone*VAAT b=-0.037 [-0.061 to -0.013] p=0.002<br>Estradiol*VAAT b=-0.056 [-0.096 to -0.017] p=0.005<br>Testosterone*VAAT b=0 [-0.061 to 0.06], p=0.98<br>Progesterone*TAAT b=0.001 [-0.013 to 0.016] p=0.84<br>Estradiol*TAAT b=0.012 [-0.013 to 0.036] p=0.35<br>Testosterone*TAAT b=0.015 [-0.022 to 0.052] p=0.42<br>Progesterone*SAAT b=0.014 [-0.007 to 0.035] p=0.19<br>Estradiol*SAAT b=0.036 [0.001 to 0.071] p=0.042<br>Testosterone*SAAT b=0.027 [-0.026 to 0.08] p=0.31<br>Progesterone*VAT/SAT b=-0.039 [-0.063 to -0.014] p=0.002<br>Estradiol*VAT/SAT b=-0.068 [-0.109 to -0.027] p=0.001<br>Testosterone*VAT/SAT b=-0.016 [-0.079 to 0.047] p=0.623<br><br>Spearman correlation unadjusted:<br>Progesterone*VAAT r=-0.249, p=0.001<br>Estradiol*VAAT r=-0.229, p=0.001<br>Progesterone*VAT/SAT r=-0.144, p=0.02<br>Estradiol*VAT/SAT r=-0.205, p=0.001<br><br>Spearman correlation adjusted for age and BMI:<br>Progesterone*VAAT r=-0.037, p=0.001<br>Estradiol*VAAT r=-0.056, p=0.005<br>Progesterone*VAT/SAT r=-0.039, p=0.002<br>Estradiol*VAT/SAT r=-0.068, p=0.001<br><br>Age and BMI |
| Follow-up: months | Sample size n=<br><br>OBSERVATIONAL ON BREAST FAT | Exposures<br><br>Outcomes |  |
| Boyd 2009<br>(41)<br>Canada | Sample size n= 280<br><br>Healthy daughters (aged | Exposures<br>Total breast fat<br>mean=277.5 cm2 | Linear regression:<br>log transformed total breast fat * log transformed sex steroids |

| | | | |
|---|---|---|---|
| Cross-sectional<br>Follow-up: NA months | mean=20.8 (sd=4.9) (range=15-30)) of 100 randomly sampled healthy women who had undergone mammography; healthy; white<br><br>Pre-menopause<br>(Follicular) | (sd=249.4)<br>MRI (calibrated - series of custom-built phantoms)<br><br>Outcomes<br>Plasma (measured by ECLIA)<br>SHBG (nmol/L)<br>Estradiol (pmol/L)<br>Progesterone (nmol/L)<br>Testosterone (nmol/L)<br><br>Calculated<br>Free testosterone (nmol/L)<br>Free estradiol (pmol/L) | SHBG<br>βu=-0.44; 95%CI=-0.66 to -0.22; p=0.0001<br>βa=-0.09; 95%CI=-0.23 to 0.06; p=0.23<br><br>Estradiol<br>βu=-0.13; 95%CI=-0.28 to 0.01; p=0.06<br>βa=0.01; 95%CI=-0.09 to 0.11; p=0.87<br><br>Progesterone<br>βu=-0.1; 95%CI=-0.28 to 0.08; p=0.28<br>βa=-0.12; 95%CI=-0.25 to 0.01; p=0.06<br><br>Testosterone<br>βu=-0.07; 95%CI=-0.26 to 0.11; p=0.44<br>βa=-0.1; 95%CI=-0.23 to 0.04; p=0.17<br><br>Free testosterone<br>βu=0.13; 95%CI=-0.01 to 0.27; p=0.06<br>βa=-0.02; 95%CI=-0.12 to 0.08; p=0.72<br><br>Free estradiol<br>βu=-0.02; 95%CI=-0.16 to 0.11; p=0.74<br>βa=0.04; 95%CI=-0.07 to 0.15; p=0.5<br><br>u=unadjusted<br>a=adjusted for time since last menstrual cycle (days), age (years), age at menarche (years), weight (kg), and height (cm). |
| Denholm 2018<br>(110)<br>England<br>Cross-sectional<br>Follow-up: NA months | Sample size n= All=117<br>BMI<25=85<br>BMI>=25=32<br><br>nulliparous women born from singleton pregnancies<br>age mean=21.5 (sd=0.9)<br><br>Pre-menopause<br>(Mix but adjusted for it) | Exposures<br>Total breast fat volume mean=408.9 cm3 (sd=353.8)<br>MRI left-right average fat volume<br><br>Outcomes<br>Plasma at age 21.5 years old<br><br>RIA<br>DHEA: mean=28.6 (nmol/l) (sd=12.8)<br>Androstenedione: mean=7.1 (nmol/l) (sd=3.1)<br>Testosterone: mean=1.6 (nmol/l) (sd=0.7)<br>SHBG: mean=67.5 | Linear regression shoed the relative difference (RD): Exponentiated estimated regression parameters and CI<br><br>Standardised plasma sex steroids z-score and breast measures log-transformed at age 21.5y<br><br>DHEA<br>βu=1; 95%CI=0.86 to 1.16, NS<br>βa=0.95; 95%CI=0.87 to 1.04, NS<br><br>Androstenedione<br>βu=1.01; 95%CI=0.87 to 1.17, NS<br>βa=0.98; 95%CI=0.89 to 1.07 NS<br><br>Testosterone<br>βu=0.98; 95%CI=0.85 to 1.13 NS<br>βa=0.97; 95%CI=0.89 to 1.06 NS<br><br>SHBG<br>βu=0.86; 95%CI=0.74 to 0.99, S<br>βa=1.03; 95%CI=0.93 to 1.14, NS |

| | | | |
|---|---|---|---|
| | | (nmol/l) (sd=32.9)<br><br>Indirect RIA<br>Estrone: mean=260.7 (pmol/l) (sd=159.9)<br>Estradiol: mean=318.6 (pmol/l) (sd=283.6) | Estrone<br>βu=0.94; 95%CI=0.80 to 1.09, NS<br>βa=0.92; 95%CI=0.83 to 1.01, NS<br><br>Estradiol<br>βu=0.89; 95%CI=0.77 to 1.03, NS<br>βa=0.88; 95%CI=0.81 to 0.98, S<br><br>βu: adjusted for assaybatch number, storage time, and age and menstrual phase at MRI examination;<br>βa: further adjusted for BMI at MRI examination |
| Linton L* 2016<br>(42)<br>Canada<br>Cross-sectional<br>Follow-up: NA months | Sample size n= 225<br><br>healthy whiteWhite young women aged 15–30<br>age mean=19.9 (sd=4.7)<br><br>Pre-menopause<br>(Luteal, follicular separate) | Exposures<br>Total breast fat volume mean=502.8 cm3 (sd=313.9)<br>MRI<br><br>Outcomes<br>Luteal phase plasma (ECLIA)<br>SHBG (nmol/l): median=62.7 (IQR=35.5)<br>Estradiol (pmol/l): median =434 (IQR =259)<br>Free estradiol (pmol/l): median =5.8 (IQR =3.2)<br>Progesterone (nmol/l): median =23 (IQR =29)<br>Testosterone (nmol/l): median =0.9 (IQR =0.7)<br>Free testosterone (nmol/l): median =0.008 (IQR =0.006)<br><br>Follicular phase<br>SHBG (nmol/l): median=54.5 (IQR=27.4)<br>Estradiol (pmol/l): median =187 (IQR =143)<br>Free estradiol (pmol/l): median =2.7 (IQR =1.8) | Linear regression<br>log transformed total breast fat volume * log-trnsformed SS (except testosterone)<br><br>Luteal phase<br>SHBG: βu=-0.62; p=0.0001 / βa=-0.24; p=0.09<br>Estradiol: βu=-0.11; p=0.32 / βa=-0.006; p=0.95<br>Free estradiol: βu=0.22; p=0.05 / βa=0.11; p=0.19<br>Progesterone: βu=-0.16; p=0.012 / βa=-0.09; p=0.066<br>Testosterone: βu=0.11; p=0.31 / βa=0.004; p=0.95<br>Free testosterone: βu=0.29; p=0.0001 / βa=0.08; p=0.18<br><br>Follicular phase<br>SHBG: βu=-0.58; p<0.0001 / βa=-0.22; p=0.013<br>Estradiol: βu=-0.047; p=0.57 / βa=-0.038; p=0.6<br>Free estradiol: βu=0.1; p=0.22 / βa=0.035; p=0.63<br>Progesterone: βu=-0.12; p=0.23 / βa=-0.12; p=0.11<br>Testosterone: βu=-0.038; p=0.44 / βa=-0.066; p=0.08<br>Free testosterone: βu=0.22; p=0.007 / βa=0.04; p=0.53<br><br>βu: unadjusted<br>βa: age at MRI ; age at Menarche; weight ; height and days since LMP (most recent menstrual period) for each phase |

| | | | |
|---|---|---|---|
| | | Progesterone (nmol/l): median =2.0 (IQR =1)<br>Testosterone (nmol/l): median =1.7 (IQR =1.1)<br>Free testosterone (nmol/l): median =0.015 (IQR =0.01) | |
| Nayeem 2014<br>(25)<br>USA<br>Cross-sectional<br>Follow-up: months | Sample size n= 137<br><br>Healthy premenopausal women of all major races/ethnicities, living within 80 km of Galveston, Texas, between 30 to 40 years with regular monthly menstrual cycles.<br><br>Pre-menopause<br>(Luteal) | Exposures<br>fat tissue volume (FV)<br><br>measured by five different methods: Histogram segmentation method (HSM-2D mammography), Full-field digital mammography (FFDM-2D mammography), Mathemathical algorithm (MATH-2D mammography), 3D gradient-echo MRI (3DGREE), Short tau inversion recovery MRI (STIR)<br><br>Outcomes<br>17b-estradiol (pg/ml), progesterone (ng/ml) - measured by radioactive immunoassay (RIA) | Standardized β-estimates (SE)<br>Fat tissue volume*<br>Estradiol 0.02 (0.06), NS<br>Progesterone −0.02 (0.05), NS<br><br>Multivariate linear regression:<br>Adjusted for: adjusted for BMI, age and reproductive variables known to influence BD, such as age of menarche and number of completed pregnancies,different biomarkers, race |
| Urine and other excreted matrices | | | |
| Rochester 2009<br>(43)<br>USA<br>Non-randomized intervention<br>Follow-up: 6-12 months | Sample size n= 43 but 9 post surgery<br><br>Obese pre-op: 23 (FU: 6); controls: 14<br><br>Premenopausal (all phases but considered)<br><br>Obese BMI >=35 kg/m2, planning to undergo bariatric surgery and controls<br><br>Age: intervention group= 36.6 ± | Exposures<br>BMI (kg/m^2)<br><br>Outcomes<br>Whole cycle E1c (ng/mg Cr, enzyme-linked immunosorbent assay);<br>Luteal Pdg (mcg/mg Cr, enzyme-linked immunosorbent assay) | Whole cycle E1c: mean (SD) (ng/mg Cr)<br>Baseline (contol vs obese)<br>1278.2 (475.0) vs 1026 (336.3), p=0.07<br><br>Obese (pre-post operation)<br>1026 (336.3) vs. 605.4 (438.3), p<0.001<br><br>Luteal Pdg: mean (SD) Pdg (mcg/mg Cr)<br>Baseline (contol vs obese)<br>151.7 (111.1) vs. 32.8 (10.9), p<0.001<br><br>Obese (pre-post operation)<br>32.8 (10.9) vs. 73.7 (30.5), p<0.05 |

| | | | |
|---|---|---|---|
| | 4.7, controls=31.7 ± 4.6<br><br>Pre-menopause (Luteal) | | BMI<br>Obese pre-op: 47.3 ± 5.2 vs.32.0 ± 2.9 kg/m2, change = 27.7% (range 25–37% at 6 months post-op)<br>Controls: 21.5 ± 3.4 kg/m2<br><br>Hormone concentrations were adjusted for glycerol and normalized to creatinine |
| Jain 2007<br>(44)<br>USA<br>cross-sectional<br>Follow-up: Full menstural cycle months | Sample size n= High BMI: 17;<br>Urine study controls: 11;<br>Luteinizing Hormone study controls: 12<br><br>Premenopause (all but normalised)<br><br>Eumenorrheic women, BMI >=35 kg/m2;<br>Age: High BMI=35.7 ±1.9, Urine study controls=31.5 ±1.5, Luteinizing Hormone study controls=24.9±1.4<br><br>Pre-menopause (all but normalised) | Exposures<br>BMI (kg/m2)<br><br>Outcomes<br>E1c (ng/mg Cr, ELISA)<br>Pdg (µg/mg Cr, ELISA)<br>Daily urinary hormone concentrations were summed, after normalization to a 28-d cycle. | E1c, p for difference=0.21<br>Obese: 944.5 ng/mg Cr (75.8)<br>Normal: 1097.5 ng/mg Cr (91.2)<br><br>Pdg, p for difference=0.002<br>Obese: 38.2 ng/mg Cr (2.1)<br>Normal: 181.3ng/mg Cr (35.1)<br><br>BMI:<br>High BMI=48.6±1.4 kg/m2, Urine study controls=21.3± 0.4 kg/m2, Luteinizing Hormone study controls=20.8 ±0.5 kg/m2<br><br>Hormone concentrations were adjusted for glycerol and normalized to creatinine |
| Santoro 2004<br>(45)<br>USA<br>cross-sectional<br>Follow-up: months | Sample size n= 848<br><br>Premenopausal (all phases not stratified)<br><br>Women selected from five ethnic groups in the US, Caucasian, African, Chinese, Japanese, and Hispanic;<br><br>mean BMI, 27.3 kg/m2;<br>Age: 47.2 (range= 43-53)<br><br>Pre-menopause (all not stratified) | Exposures<br>BMI (kg/m2)<br><br>Outcomes<br>E1c: estrone conjugate total cycle ( ng/mg Cr, chemiluminescents-assays)<br><br>Pdg: pregnanediol glucuronide total cycle (µg/mg Cr, chemiluminescents-assays) | E1c by BMI category<br>Low: 1307.2 (1252.9, 1363.9); Medium: 1272.7 (1201.1, 1348.6); High: 1253.3 (1180.9, 1330.0)<br>P=0.50<br><br>Pdg by BMI category<br>Low: 62.8 (59.3, 66.6); Medium: 52.5 (48.5, 56.8); High: 40.7 (37.5, 44.1)<br>p<0.0001<br><br>Adjusted for study center |
| Smith 2013<br>(27)<br>USA<br>Randomized clinical trial<br>Follow-up: 4 ± 2 weeks months | Sample size n= Exercisers: 212 (FU=165);<br>controls: 179 (FU=153)<br><br>Premenopausal (follicular phase)<br><br>BMI=18–40 kg/m2;<br><br>Age: exercisers=25.4 (SE=0.3), controls=25.2 (SE=0.3) | Exposures<br>BMI (kg/m^2), Weight (kg), Fat mass (kg, DEXA), % Body fat (%, DEXA)<br><br>Outcomes<br>Estrone (nmol/d, LC/MS-MS), Estradiol | Exercisers experienced significant decreases in fat mass (0.57 vs. 0.04 kg) and percent body fat (0.95% vs. 0.09%). Controls do not.<br><br>We only extract exercisers not the controls<br><br>Changes in sex steroids pre and post aerobic intervention of 16 weeks.<br>Age- and BMI-adjusted geometric means in Exercisers<br>E1: 23.1 (19.0–28.2) to 23.0 (18.9–28.0), NS<br>E2: 7.8 (6.8–8.8)to 8.3 (7.2–9.5), NS<br>E3: 21.3 (16.5–27.4) to 18.6 (14.4–24.2), NS |

| | | | |
|---|---|---|---|
| | Pre-menopause (follicular) | (nmol/d, LC/MS-MS), Estriol (nmol/d, LC/MS-MS), 2-OHE1 (nmol/d, LC/MS-MS), 2-OHE2 (nmol/d, LC/MS-MS), 16a-OHE1 (nmol/d, LC/MS-MS), plus further estrogen metabolites on the 2Me and 4OH and 4Me pathways (nmol/d, LC/MS-MS).<br><br>3 consecutive 24-hour periods in the midfollicular phase | 2-OHE1: 39.3 (33.7–45.8) to 44.2 (38.4–50.8),NS<br>16a-OHE1: 2.9 (2.3–3.7) to 2.6 (2.1–3.4),NS<br>2:16a-OHE1: 13.4 (10.4–17.2) to 16.8 (13.1–21.4), p=0.043<br><br>No associations observed for any of the other estrogen metabolites 4-OHE1, 2:4-OHE1, 2-MeOE1, 4-MeOE1, 2-OHE2, 4-OHE2, 2-MeOE2,4-MeOE2<br><br>Unadjusted comparisons of baseline characteristics. Changes from baseline comparisons compared on original scale. All comparisons adjusted for study design, age and BMI strata with a general linear model. If significant differences at baseline in an outcome, follow-up and change from baseline -> additionally adjusted for baseline values |
| Westerlind 2007<br>(111)<br>USA<br>Diet and exercise intervention<br>Follow-up: 4 months | Sample size n= 31<br><br>Premenopausal (all phases but stratified)<br><br>Sedentary<br><br>Age: 31.5 (range: 25-40, SE=0.9)<br><br>Pre-menopause (all but stratified) | Exposures<br>BMI, % body fat, fat mass (kg) (latter two by hydrostatic weighing)<br><br>Outcomes<br>2OHE1, 16α-OHE, 2OHE1/16α-OHE | Change pre- to post intervention<br>Weight: 63.3 (1.8) to 59.5 (1.7), p=0.001<br>BMI: 23.7 (0.5) to 22.2 (0.5), p=0.001<br>Body fat %: 31.6 (1.0) to 27.0 (1.2), p=0.001<br>Fat mass: 20.2 (1.1) to 16.3 (1.0), p=0.001<br><br>2-OHE1<br>Luteal 14.6(1.18) to 16.4 (2.2), p=0.25<br>Follicular 25.8 (2.8) to 21.3 (2.1), p=0.06<br><br>16α-OHE1<br>Luteal 8.9 (0.9) to 9.1 (0.8), p=0.42<br>Follicular, 14.6 (1.3) to 12.1 (1.2), p=0.02<br><br>2/16-OHE1<br>Luteal 1.9 (0.2) to 2.0 (0.2), p=0.24<br>Follicular 2.0 (0.2) to 1.9 (0.2), p=0.44<br>Luteal+Foll 2.0 (0.2) to 1.9 (0.2), p=0.12<br><br>Correlation at baseline<br>follicular 2-OHE1 *BMI: r=-0.49, p=0.02<br>Other phase and other body composition with 2OHE1, 16α-OHE1, or 2:16 NS<br><br>unadjusted |
| Campbell 2005<br>(112)<br>Canada<br>cross-sectional<br>Follow-up: months | Sample size n= High fitness: 17; Average fitness: 13<br><br>Premenopausal (all phases but stratified) | Exposures<br>Weight (kg), BMI (kg/m2), sum of skinfolds (mm)<br><br>Outcomes | Pearson correlations:<br>Follicular, BMI<br>2-OHE1 (r=-0.37, p=0.04)<br>16αOHE1 (r=0.25, p=0.18)<br>2:16αOHE1 (r=-0.40, p=0.03) |

| | | | |
|---|---|---|---|
| | Healthy women,<br><br>BMI=18–24 kg/m2;<br>Age: High fitness=29.5 (SE=1.7), average fitness=27.6 (SE=1.6)<br><br>Pre-menopause (all but stratified) | Follicular 16α-OHE1 (ng.mL-1.mg-1 creatinine, enzyme-linked immunoassay), Luteal 16α-OHE1 (ng.mL-1.mg-1 creatinine, enzyme-linked immunoassay), Follicular 2-OHE1 (ng.mL-1.mg-1 creatinine, enzyme-linked immunoassay), Luteal 2-OHE1 (ng.mL-1.mg-1 creatinine, enzyme-linked immunoassay), Follicular 2:16α-OHE1 (ng.mL-1.mg-1 creatinine, enzyme-linked immunoassay), Luteal 2:16α-OHE1 (ng.mL-1.mg-1 creatinine, enzyme-linked immunoassay) | Follicular, skinfolds<br>2-OHE1 (r=-0.33, p=0.07)<br>16αOHE1 (r=0.11, p=0.54)<br>2:16αOHE1 (r=-0.29, p=0.12)<br><br>Luteal, BMI<br>2-OHE1 (r=0.06, p=0.76)<br>16αOHE1 (r=0.33, p=0.07)<br>2:16αOHE1 (r=-0.23, p=0.20)<br><br>Luteal, skinfolds<br>2-OHE1 (r=0.01, p=0.94)<br>16αOHE1 (r=0.39, p=0.03)<br>2:16αOHE1 (r=-0.41, p=0.02)<br><br>Unadjusted |
| MacMahon 1982<br>(28)<br>USA<br>cross-sectional<br>Follow-up: months | Sample size n= 511<br><br>Premenopausal (all phases but stratified)<br><br>Aged 30-39 years<br><br>Pre-menopause (all but stratified) | Exposures<br>Quetelet's index (BMI (weight/height^2))<br><br>Outcomes<br>Estrone, estradiol, estriol<br><br>measured at follicualr phase (on the 10th day)<br><br>Luteal phase (n the 20 to 21th day) distributed as such<br>anovular (3-11 days prior menses, pregnanediol<1mg/L)<br>ovular (3-11 days prior menses, pregnanediol>1mg/L<br>inderterminate (out 3-11 days prior menses) | No significant association between BMI and follicular or luteal E1, E2, or E3; adjusted for age, age at menarche, and study center<br><br>Regression coefficients (SE) for BMI and log-gravity corrected estrogen<br>Women 30-39y:<br>Follicular (n = 321)<br>El -0.049 (0.061)<br>E2 -0.038 (0.063)<br>E3 -0.031 (0.081)<br><br>Luteal anovular (n = 17)<br>El +0.023 (0.404)<br>E2 +0.310 (0.287)<br>E3 -0.350 (0.768)<br><br>Luteal ovular (n = 262)<br>El +0.044 (0.056)<br>E2 +0.029 (0.051)<br>E3 -0.055 (0.073)<br><br>Luteal undetermined ovular or anovular (n = 41)<br>El +0.056 (0.159)<br>E2 +0.032 (0.180)<br>E3 -0.096 (0.213)<br>All NS |

| | | | |
|---|---|---|---|
| | | | Study center, age, age at menarche |
| Trichopoulos 1983<br>(29)<br>Greece<br>cross-sectional<br>Follow-up: months | Sample size n= women aged 18-23: 122; women aged 30-40: 37<br><br>Premenopausal (all phases but stratified)<br><br>Aged 18-23 (weight=57.2 kg) and never pregnant;<br><br>Aged 30-40 (weight=64.5 kg) have had a full-term pregnancy<br><br>Pre-menopause (all but stratified) | Exposures<br>Quetelet's index (BMI (weight/height^2))<br><br>Outcomes<br>estrone (µg/g creatinine, determined by the method described by Brown 1976), estradiol (µg/g creatinine, determined by the method described by Brown 1976) from urine samples collected on day 20 or 21 of menstrual cycle | Partial regression coefficient (SE) between BMI and log estrogen concentration:<br><br>Younger women 18-23y,<br>Luteal anovular (n=20)<br>Estrone -0.397 (0.238)<br>Estradiol -0.381 (0.274)<br>Estriol -0.245 (0.257)<br><br>Luteal ovular (n=63)<br>Estrone -0.119 (0.169)<br>Estradiol -0.004 (0.134)<br>Estriol 0.110 (0.143)<br><br>Luteal undetermined ovular or anovular (n=35)<br>Estrone -0.197 (0.222)<br>Estradiol -0.109 (0.222)<br>Estriol -0.029 (0.183)<br><br>Older women 30-40y<br>Luteal ovular (n=26)<br>Estradiol -0.052 (0.233)<br>Estrone -0.020 (0.205)<br>Estriol -0.204 (0.247)<br><br>All not statistically significant; similarly no associations for weight. Associations adjusted for age and age at menarche.<br><br>multiple regression - controlled for current age and age at menarche |
| Windham 2002<br>(46)<br>USA<br>Cross-sectional<br>Follow-up: Up to 6 months | Sample size n= 411;<br>Underweight: 8%; Average: 72%; Overweight: 20%<br><br>Premenopausal (all phases but stratified)<br><br>Underweight (BMI <19.1),<br>Average (BMI =19.1-27.3)<br>Overweight (BMI >27.3)<br><br>willing to collect and freeze urine samples first thing in the morning for up to 6 months;<br>Age: 18-39<br><br>Pre-menopause (all but stratified) | Exposures<br>BMI (kg/m2)<br><br>Outcomes<br>Pregnanediol-3-glucuronide (PdG) (µg/ml creatinine, enzyme immuno-assays), estrone conjugates [E1c] - estrone sulfate and estrone glucuronide - (ng/mg creatinine, enzyme immuno-assays) | Mixed models that account for repeated measures using each cycle as one unit of observation,<br><br>Beta (CI) relative - normal weight as reference category<br><br>PdG<br>Baseline<br>Underweight, -0.03 (-0.20 to 0.14)<br>Overweight, -0.09 (-0.21 to 0.02)<br><br>Follicular<br>Underweight, -3.09 (-11.8 to -5.66)<br>Overweight, -9.25 (-14.9 to -3.58), SS<br><br>Progesterone_Peak<br>Underweight, -0.16 (-1.20 to 0.89)<br>Overweight, -0.71 (-1.38 to -0.02), SS |

| | | | |
|---|---|---|---|
| | | | Luteal<br>Underweight, -0.19 (-0.92 to 0.53)<br>Overweight, -0.54 (-1.01 to -0.07), SS<br><br>E1c<br>Baseline<br>Underweight, 0.26 (-3.80 to 4.31)<br>Overweight, -0.73 (-3.43 to 1.97)<br><br>Follicular<br>Underweight, 54.3 (-38.0 to 146.5)<br>Overweight, 3.3 (-58.3 to 64.9)<br><br>Ovulation<br>Underweight, 2.25 (-7.91 to 12.4)<br>Overweight, 0.24 (-6.66 to 7.14)<br><br>Luteal<br>Underweight, 0.27 (-6.80 to 7.33)<br>Overweight, 3.12 (-7.73 to 1.50)<br><br>adjusted for age, race, education, employed, prior pregnancy, induced abortion, miscarrage, age at menarche |
| Xie 2012<br>(30)<br>USA<br>cross-sectional<br>Follow-up: months | Sample size n= 603<br><br>Premenopausal (luteal phase)<br>BMI: 25.0 (SD=5.5)<br>BMI at age 18: 21.1 (SD=2.9)<br>Age: 42.8 (SD=3.8)<br><br>Pre-menopause (luteal) | Exposures<br>Current BMI (kg/m2),<br>BMI at age 18 (kg/m2)<br><br>Outcomes<br>Estradiol (pmol/mg creatinine, LC-MS/MS),<br>Estrone (pmol/mg creatinine, LC-MS/MS),<br>Estriol (pmol/mg creatinine, LC-MS/MS),<br>16-OHE-1 (pmol/mg creatinine, LC-MS/MS),<br>2-OHE-1 (pmol/mg creatinine, LC-MS/MS) | Linear regression, percentage change for one unit increase of BMI (categorised as 20,22.5, 25,27.5, 30) when treating it as a continuous variable.<br>Current BMI:<br>Estradiol: 0.1, ptrend=0.80<br>Estrone: -0.80, ptrend=0.11<br>Estriol: 0.6, ptrend=0.29<br>16-OHE1: -1.7, ptrend=0.01<br>2-OHE1: -3.4, ptrend<0.001<br><br>Percentage change for one unit increase of BMI age 18 (categorised as 20,22.5, 25) when treating it as a continuous variable.<br>BMI age 18:<br>Estradiol: -1, ptrend=0.26<br>Estrone, -1.3, ptrend=0.19<br>Estriol: 0, ptrend=0.97<br>16-OHE1: -1.2, ptrend=0.33<br>2-OHE1: -3.4, ptrend=0.01<br><br>Adjusted for age at urine collection, time of day of urine collection, month of collection, history of benign breast disease, parity, duration of past oral contraceptive use, smoking, physical activity, menstrual cycle irregularity and ovulatory status of menstrual cycle which urine was collected |
| Ziomkiewicz 2008<br>(113)<br>Poland | Sample size n= 141<br><br>Premenopausal (all phases but | Exposures<br>% Body fat (%, BIA) | <br>% Body fat and Estradiol over full menstrual cycle (NS)<br>% Body fat and Day -1 Estradiol (p<0.05) |

| | | | |
|---|---|---|---|
| cross-sectional<br>Follow-up: months | stratified)<br><br>Normal weight (BMI: 22.6 (SD=2.84)) premenopausal women with regular menstural cycles<br>Age: 29.8 (SD=3.34)<br><br>Pre-menopause (all but stratified) | Outcomes<br>Estradiol (pmol/L ,RIA) | % Body fat and Day 0 Estradiol (p<0.05)<br>% Body fat and Mean follicular phase Estradiol (p<0.05)<br>% Body fat and Mid-cycle Estradiol (p<0.05)<br>% Body fat and Mean luteal phase Estradiol (NS)<br><br>Table II is not very practicle to read, we understood that where there is letteres the anova is significant. Hope that's correct. But the table III showed not the same results for all women.<br><br>The study also stratify by positive/negative energy balance (change in body fat % between 1st and 2nd measurment -1% is threshold).<br><br>The text says:<br>women with very low and high body fat<br>percentage had significantly lower levels of estradiol during<br>the follicular phase (21.1 versus 17.0 and 15.9 pmol/l,<br>F3,124 ¼ 3.22, P ¼ 0.025), mid-cycle (25.4 pmol/l versus<br>19.7 and 18.2 pmol/l, F3,126 ¼ 4.03, P ¼ 0.009) and on Day<br>21 (38.8 pmol/l versus 29.1 and 26.6 pmol/l, F3,123 ¼ 4.60,<br>P ¼ 0.004) and Day 0 (20.4 pmol/l versus 15.2 and<br>13.4 pmol/l, F3,122 ¼ 3.35, P ¼ 0.021). Similar differences<br>were also observed in comparisons between women with low<br>body fat and women with very low and high body fat.<br><br>ANOVA between quartiles of body fat percentage and estradiol stratified by menstrual cycle phase |
| Napoli 2012<br>(114)<br>USA<br>cross-sectional<br>Follow-up: months | Sample size n= 97<br><br>Postmenopausal women; BMI: 28.91 (SD=6.23); ≥ 1 year from the last normal menstrual period or those who have had oophorectomy;<br>Age: 63.73 (SD=7.52)<br><br>Post-menopause | Exposures<br>% total fat (%, dual-energy X-ray absorptiometry)<br>%truncal fat (%, dual-energy X-ray absorptiometry)<br>BMI (kg/m2)<br><br>Outcomes<br>2OHE1 (ng/mg creatinine, monoclonal antibody-based competitive enzyme immunoassays for estrogen metabolites), 16α-OHE (ng/mg creatinine, monoclonal antibody-based competitive enzyme immunoassays for estrogen metabolites), 2OHE1/16α-OHE (ratio, | Total fat percentage, Pearson correlation with<br>2OHE1, r=-0.27, p<0.01<br>16α-OHE, r=-0.14, p=0.19<br>2OHE1/16α-OHE, r=-0.22, p=0.03<br>Free estradiol index, r=0.24, p=0.07<br><br>Truncal fat, Pearson correlation with<br>2OHE1, r=-0.32, p<0.01<br>16α-OHE, r=-0.19, p=0.06<br>2OHE1/16α-OHE, r=-0.10, p=0.31<br>Free estradiol index, r=0.35, p<0.01<br><br>BMI, Spearman correlation with<br>2OHE1, r=-0.36, p<0.01<br>16α-OHE, r=-0.12, p=0.25<br>2OHE1/16α-OHE, r=-0.29, p<0.01<br>Free estradiol index, r=0.50, p<0.01 |

| | | | |
|---|---|---|---|
| | | caluclated from 2OHE1 and 16α-OHE) | |
| Atkinson 2004<br>(115)<br>USA<br>Intervention (lifestyle)<br>Follow-up: 12 months | Sample size n= 173<br><br>Overweight and obese, previously sedentary, postmenopausal women, BMI: 25–40 kg/m2 or BMI between 24-25 kg/m2 and % body fat > 33%;<br>Age: 50-75 years<br><br>Post-menopause | Exposures<br>Intraabdominal fat (cm2, computed tomography)<br><br>Outcomes<br>2-OHE1 (ng/mg Cr, Estramet 2/16 enzyme immunoassay (EIA) kits), 16a-OHE1 (ng/mg Cr, Estramet 2/16 enzyme immunoassay (EIA) kits), 2:16 ratio (ratio, calculated) | Exercise group lost an average of 1.29 kg (SD 3.8 kg),<br>Stretching control group gained an average of 0.1 kg (SD 3.0 kg) (P < 0.01)<br><br>Table 2 and 3 show concentration of metabolites at baseline and 12 months in Exercisers and controls but no test is provided to compare baseline to FU within each group (test accross exercisers and controls only).<br><br>Correlation between baseline intraabdominal fat<br>2-OHE1, r =-0.17, p=0.03<br>16a-OHE1, r =-0.20, p=0.01<br>2:16 ratio, r = -0.02, p=0.75<br>No significant correlations with baseline BMI, percentage body fat, and subcutaneous fat.<br><br>2-OH E1 and 16a-OH E1 adjusted for creatinine excretion |
| Fowke 2001<br>(116)<br>USA<br>cross-sectional<br>Follow-up: months | Sample size n= 37<br><br>Healthy, free-living, postmenopausal women, BMI: 27.7 (SD=4.9);<br>Age: 61.7 (SD=8.2)<br><br>Post-menopause | Exposures<br>BMI (kg/m2), WHR (calculated), Body density (calculated based on weight WC, age, height, other prediction variables)<br><br>Outcomes<br>2-hydroxyestrone/16α-hydroxyestrone (ratio, calculated from 2-hydroxyestrone and 16-hydroxyestrone) | Spearman correlations with 2:16α-OHE<br>BMI, r=−0.22, p=0.20<br>Body fat, r=-0.20, p=0.23<br>WHR, r=-0.22, p=0.19<br><br>Beta estimates (adjusted for age and serum E2<br>BMI, b=−0.05, 95%CI [-0.11;0.02]<br>Body fat, b=−0.02, 95%CI [-0.05;0.01]<br>WHR, b=−2.55, 95%CI [-8.14;3.01]<br><br>Adjusted for age, E2 levels, and dietary parameters in multivariable linear regression model |
| de Waard 1982<br>(26)<br>Netherlands<br>cross-sectional + intervention (weight loss)<br>Follow-up: 6-12 months | Sample size n= 42<br><br>obese postmenopausal women; over 10 kg overweight( according to Broca's rule);<br>Age: 52-67<br><br>Post-menopause | Exposures<br>Weight change (kg), weight (kg), BMI (kg/m2)<br><br>Outcomes<br>E1 (ng/g Cr, RIA), E2 (ng/g Cr, RIA), E3 (ng/g Cr, RIA), total E (sum of E1, E2 and E3) | Correlations between weight changes and biomarker changes, after weight loss: decrease of E1 and E3.<br>E1, r=0.33, p=0.017<br>E2, r=0.15, p=0.166<br>E3, r=0.32, p=0.018<br>Total E, r=0.34, p=0.012<br><br>Not reported |
| MacDonald 1978<br>(117)<br>USA<br>cross-sectional<br>Follow-up: months | Sample size n= 25<br><br>Postmenopausal women with no endometrial disease, controls from case-control study, Weight: 194 (SE=12) pounds;<br>Age: 62 (no measure of variance | Exposures<br>Excessive weight (pounds, body weight exeeding the upper limit of ideal weight for height and medium body frame determined | Correlations<br>body weight and % A4 to E1 transformation, 0.6958, p<0.001<br>excessive weight and % A4 to E1 transformation, 0.6526, p<0.001 |

| | | | |
|---|---|---|---|
| | reported)<br><br>Post-menopause | from actuarial data from Metropolital Life Insurance company), weight (pounds)<br><br>Outcomes<br>Conversion of plasma androstenedion to urinary estrone (in %) | |
| Mole 1989<br>(118)<br>UK<br>cross-sectional<br>Follow-up: months | Sample size n= 132<br><br>"Apparently fit" active, and healthy postmenopausal women, menopause for at least 1 year, weight: 56 kg (coefficient of variation: 0.7 %);<br>Age: 47-80<br><br>Post-menopause | Exposures<br>Weight (kg)<br>BMI (kg/m2)<br><br>(Indirect body fat variables calculated not measured:<br>Body fat %<br>Body fat<br>Fat mass)<br><br>Outcomes<br>estrone-3-D-glucuronide (nmol/l, RIA), estradiol-3-D-glucuronide (nmol/l, RIA), estrone (pmol/l, RIA), estradiol (pmol/l, RIA) | BMI, correlations age-adjusted<br>E1: 0.251, p<0.01<br>E1G: 0.204, p<0.05<br>E2: 0.204, p<0.05<br>E2G: 0.251, p<0.01<br><br>correlations, not adjusted<br>weight<br>E1: 0.182, p<0.05<br>E1G: 0.245, p<0.01<br>E2: 0.174, p<0.05<br>E2G: 0.254, p<0.01<br><br>body fat% and<br>E1: 0.160, NS<br>E1G: 0.239, p<0.01<br>E2: 0.121, NS<br>E2G: 0.298, p<0.01<br><br>body fat and<br>E1: 0.137, NS<br>E1G: 0.259, p<0.01<br>E2: 0.141, NS<br>E2G: 0.309, p<0.01<br><br>fat mass and<br>E1: 0.208, p<0.05<br>E1G: 0.255, p<0.01<br>E2: 0.188, p<0.05<br>E2G: 0.266, p<0.01<br><br>BMI correlations adjusted for age |
| Carpenter 2012<br>(6)<br>USA | Sample size n= 7<br><br>Healthy obese postmenopausal | Exposures<br>BMI (kg/m2), fat mass (kg, DEXA) | Non-periodized resistence training<br>Baseline to 3 Months, mean (SD), change<br>fat mass: 41.3 (11.9) to 35.5 (10.9), -14.4 (-18.5,-10.3), S |

| | | | |
|---|---|---|---|
| weight-loss lifestyle intervention<br>Follow-up: 12 weeks months | women,<br><br>BMI: 33.6 kg/m2(SD=5.3)<br><br>Age: 55.3 (SD=5.5)<br><br>Post-menopause | Outcomes<br>Estradiol (nmol/g protein, GC-MS/MS) | BMI: 33.6 (5.3) to 29.8 (3.8), -11.0 (-13.9,-8.1), S<br>Weight: 89.8 (15.3) to 80.3 (12.2), -10.3 (-12.6, -8.0), S<br><br>Estradiol: 10.5 (5.7) to 8.1 (4.3), -23.9 (-113.1,-10.5), S<br><br>no |
| Kirschner 1990<br>(119)<br>USA<br>cross-sectional<br>Follow-up: months | Sample size n= 29<br><br>Premenopausal (unknown)<br><br>Morbidly obese women (BMI>30)<br><br>Age: 20-40 years<br><br>Pre-menopause (unknown) | Exposures<br>% Body fat (%, electrical plethysmography)<br><br>Outcomes<br>Estrone production rates (nmol/day), peripheral aromatization (blood to urine) A4 to to E1 (%) | E1 production rate<br>mean (SD), nmol/day, p for difference<0.05<br>Upper body obesity, 356 (+/-241) nmol/d<br>Lower body obesity, 466 (+/-295) nmol/d<br><br>Peripheral aromatization, %, p<0.05<br>Upper body obesity, 1.67 (+/-0.80)<br>Lower body obestiy, 2.54 (+/-1.24)<br><br>% Body fat:<br>Upper body obesity=43.3±1.0, Lower body obestiy=38.9± 0.8<br><br>Not reported |
| Zhao 2023<br>(120)<br>China<br>cross-sectional<br>Follow-up: months | Sample size n= 135<br><br>135 premenopausal women stratified into 4 BMI groups (n=30 underweight <18.5, n=40 normal 18.5–24.0, n=35 overweight 24.0–<28.0, n=30 obese ≥28.0); ages 35–55; obese subgroups: simple obesity (n=30) and obesity with abnormal glucose metabolism (n=30)<br><br>premenopausal | Exposures<br>BMI (kg/m²)<br><br>Outcomes<br>Urinary E2, E1, 2-OHE1, 2-OHE2, 4-OHE2, 16α-OHE1, 2-MeOE2, 2-MeOE1 (ng/mg or pg/mg creatinine) via HPLC-MS | E2, E1, 16α-OHE1, 2-OHE1 increased with BMI (P<0.05); |
| Tissues | | | |
| Metwally 2007<br>(121)<br>UK<br>cross-sectional<br>Follow-up: months | Sample size n= 28 had immunocytochemistry measured for ER and PR<br><br>Premenopausal (luteal phase)<br><br>Overweight (defined as BMI: 25–29.9 kg/m2) and obese (defined as BMI ≥ 30 kg/m2) women, history of recurrent miscarriage, n=136<br><br>Pre-menopause (luteal) | Exposures<br>BMI; Low BMI Group: 22.2 (0.24), n=70; High BMI Group: 30.4 (0.71), n=66<br>Bu the analysis is done for all.<br><br>Outcomes<br>ER and PR expression; median H-score | Correlations between BMI and<br>ER stromal epithelium, r=0.2, NS;<br>PR stromal epithelium, r=0.2, NS;<br>ER glandular epithelium, r=-0.01, NS;<br>PR glandular epithelium,r=-0.15, NS;<br>ER luminal epithelium, r=-0.01, NS;<br>PR luminal epithelium, r = −0.04, NS |

| | | | |
|---|---|---|---|
| Dottino 2021<br>(122)<br>USA<br>cross-sectional<br>Follow-up: months | Sample size n= 130<br>obese: 97<br>non-obese: 33<br><br>Premenopausal (follicular phase)<br><br>Women with BMI ≥ 30 or BMI ≤25;<br>Age: 30-55<br><br>Pre-menopause (follicular) | Exposures<br>BMI (kg/m2)<br><br>Outcomes<br>RNA extraction<br><br>IHC -- antibodies in glands and stroma for PR protein level , read by one pathologist<br><br>PR gene expression (RT-PCR)<br>Biopsy between days 6 – 11 of the menstrual cycle | t-test<br>PR protein level H-score (include women with Lynch syndrom)<br>Glands: p=0.08 NS<br>Stroma: PR lower in obese vs. non-obese, p=0.045<br><br>Women without inherited cancer risk:<br>PR lower in obese vs. non-obese p=0.0027<br>Women with Lynch syndrom p=0.58 NS<br><br>linear model (include women with Lynch syndrom)<br>PR*BMI B=-0.35 [-0.56;-0.14] p=0.002<br>Adjusted B=-0.49 [-0.81;-0.16] p=0.004<br><br>linear regression adjusted by race/ethnicity, education, age at menarche, number of live births, history of oral contraceptive use, daily anti-inflammatory, medication use; |
| Hatok 2011<br>(123)<br>Slovakia<br>case-control<br>(cases=endometriosis)<br>Follow-up: months | Sample size n= 101<br>Healthy controls: 46<br>cases: 55<br><br>Premenopausal (follicular phase)<br><br>Women undergoing laparoscopic surgery for pelvic pain, regular mestrual periods, BMI: cases = 26.3 (SD=4.2) and controls = 26.2 (SD=4.7);<br>Age: 25-55<br><br>Pre-menopause (Follicular) | Exposures<br>BMI (kg/m2)<br><br>Outcomes<br>Gene expression CYP19 mRNA level (PCR) | BMI and CYP19 (aromatase) expression, correlations<br>controls, r=0.28, p=0.06<br><br>Not reported |
| Quinkler 2004<br>(124)<br>UK<br>cross-sectional<br>Follow-up: months | Sample size n= 19<br><br>Premenopausal (menstrual cycle unknown) but androgen focus<br><br>Healthy women, BMI: 19.7–39.2;<br>Age: 30-50<br><br>Pre-menopause (Unknown) | Exposures<br>BMI (kg/m2)<br><br>Outcomes<br>mRNA expression of 17βHSD5 (RT-PCR) | BMI and 17-βHSD5 mRNA expression in SAT adipocytes, r=0.506, p=0.027<br><br>BMI and 17-βHSD5 mRNA expression in VAT adipocytes, and VAT and SAT pre-adipocytes r≤0.105, p>0.05)<br><br>Not reported |
| Nilsson 2007<br>(125)<br>Sweden<br>cross-sectional<br>Follow-up: months | Sample size n= non-obese: 16, obese: 17<br><br>SNP n=998 women<br><br>Healthy premenopausal (menstrual cycle unknown)<br><br>Non-obese and obese women, | Exposures<br>BMI (kg/m2)<br><br>Outcomes<br>ERalpha mRNA expression (Real-time quantitative PCR) | ERα mRNA expression (measurement 1) in SAT lower in obese vs. normal weight, p<0.001<br>ERα mRNA expression (measurement 2) in SAT lower in obese vs. normal weight, p<0.00003<br>ERα mRNA is lower in isolated adipocytes from obese women (n=9) compared to non-obese women (n=5), p=0.037;<br>ERα gene SNPs (rs2234693; rs9340799) and obesity -- no significant association |

| | | | |
|---|---|---|---|
| | BMI: < 30 (non-obese), ≥ 30 (obese);<br>Age (measure of variance not mentioned): 38±8 (non-obese), 36±6 (obese)<br><br>Pre-menopause (Unknown) | | |
| Gavin 2013<br>(126)<br>USA<br>cross-sectional<br>Follow-up: months | Sample size n= 15<br><br>Premenopausal (follicular phase) Overweight and obese women, BMI: 29.5 ± 0.5;<br>Age: 18-39<br><br>Pre-menopause (Follicular) | Exposures<br>WHR (caluclated)<br><br>Outcomes<br>gluteal SAT ERα (Westernplot), ERß protein (Westernplot), ratio | WHR and gluteal SAT<br>ERβ protein, r=-0.315, p=0.03<br>ERα/Erβ ratio, r= 0.406, p=0.01<br>Erα protein,NS<br><br>Not reported |
| Yamatani 2013<br>(127)<br>Japan<br>cross-sectional<br>Follow-up: months | Sample size n= 8<br><br>Results only on postmenopausal women, normal weight, nondiabetic, healthy except for gynecological disease for which they were undergoing surgery (n=8 with relevant measures);<br>Age: 22-79<br><br>Post-menopause | Exposures<br>BMI, mean (SD) premenopausal=21.9 (0.6);<br>postmenopausal=22.8 (0.6)<br><br>Outcomes<br>Ratio of E1/E2 in VAT and SAT; Ratio of 17B-HSD1/17B-HSD2; aromatase mRNA | BMI x aromatase mRNA VAT, r=0.23, p<0.05.<br>BMI x aromatase mRNA SAT p>0.05. |
| O'Brien 1997<br>(128)<br>USA<br>cross-sectional<br>Follow-up: months | Sample size n= 9<br><br>postmenopausal women, undergoing either reduction mammoplasty or mastectomy;<br>Age:<br><br>Post-menopause | Exposures<br>BMI<br><br>Outcomes<br>pg/g of estrone or estradiol | BMI and estrone, r=0.48, p=0.05<br>BMI and estradiol, r=0.52, p=0.03 |
| Siesler Nóbrega Belisário 2006<br>(129)<br>Brazil<br>cross-sectional<br>Follow-up: months | Sample size n= 35<br><br>postmenopausal with endometrial polyps undergoing hysterectomy,<br>BMI: 23-41;<br>Age: 45-80<br><br>Post-menopause | Exposures<br>BMI (kg/m2)<br><br>Outcomes<br>ER expression (1+ ( up to 25% positive cell), immunohistochemical method using a semiquantitative analysis) | Degree of ER expression in glandular tissue, median BMI<br>1+, 34<br>2+, 27<br>3+, 30.5<br>4+, 27<br>(p=0.02)<br><br>No significant association BMI*PR<br><br>Not reported |
| Hetemaki 2017<br>(130)<br>Finland | Sample size n= 37<br><br>postmenopausal women | Exposures<br>BMI (kg/m2) | Correlations between BMI and<br>estrone (SAT), r= 0.32, p >= 0.05<br>estrone (VAT), r=0.46, p < 0.05 |

| | | | |
|---|---|---|---|
| cross-sectional<br>Follow-up: months | undergoing surgery for nonmalignant gynecological indication (uterine fibroids (n = 13), ovarian cysts (n = 15), uterine polyp (n = 1), partial/total prolapse of the uterus (n = 7), and diagnostic laparoscopy (n = 1)),<br>BMI: 26 (range=21-37);<br>Age: 61 (range=46-84)<br><br>Post-menopause | Outcomes<br>estrone (pmol/kg, LC-MS/MS), estradiol (pmol/kg, LC-MS/MS) | estradiol (SAT), r=0.04, p >= 0.05<br>estradiol (VAT), r=0.13, p >= 0.05<br><br>Not reported |
| Campbell 2013<br>(131)<br>Canada/USA?<br>RCT; weight loss intervention<br>Follow-up: 6 months | Sample size n= 45<br>Diet Intervention: 8<br>Exercise Intervention: 14<br>Diet + Exercise Intervention: 16<br>Controls: 7<br><br>Female, overweight or obese postmenopausal women, BMI: >= 25.0 (>= 23.0 if AsianAmerican], engaging in <100 min/wk of moderate or vigorous activity;<br>Age: 50–75 years<br><br>Post-menopause | Exposures<br>Weight loss (kg), % Body fat (%, DEXA)<br><br>Outcomes<br>gene expression: ESR1 (BeadChip microarray assay), HSD17B1 (BeadChip microarray assay) | Diet intervention vs. control group<br>HSD17B1 decreased, p=0.01 (multiple comparisons adjusted, p=0.29)<br>ESR1 increased, p=0.03 (p=0.29).<br>COX2 increased (p=0.004/0.29)<br>IL1B increased (p=0.03/0.29)<br>LEP decrease (p=0.03/0.29)<br>insulin receptor INSR deacrease (p=0.02/0.29)<br><br>Diet and exercise intervention vs. control group<br>HSD17B1 decreased, p=0.02 (p=0.45)<br>ESR1 increased, p=0.02 (p=0.45).<br>LEP decrease (p=0.04/0.60)<br><br>No different changes in gene expression in the exercise group only, but lower levels of weight loss were observed and no different in % body fatness was observed.<br><br>pre- vs. post intervention; weight loss,<br>Diet, -11.3% (P < 0.001);<br>Exercise, -3.0% (P =0.03);<br>Diet and exercise, -9.4% (P < 0.0001);<br>controls, +1.0%.<br><br>Mean body fat loss :<br>diet, -12.6% (P = 0.02);<br>exercise, -3.1% (P=0.56);<br>diet and exercise, -13.2% (P < 0.01)<br><br>adjusted for multiple comparaison |
| Zhao 2018<br>(132)<br>USA<br>cross-sectional<br>Follow-up: months | Sample size n= 112<br>Premenopausal: 57<br>Postmenopausal: 55<br><br>Healthy women, BMI: 18.5-24.9 (normal), 25-29.9 (overweight), ≥ 30 (obese);<br>Age: 35-60 | Exposures<br>BMI (kg/m2)<br><br>Outcomes<br>Gene expression: ESR1 (Quantitative RT-PCR Using TaqMan Low-Density Assay (TLDA) Microfluidic Cards), | ANOVA by BMI category:<br><br>ESR1 mRNA:<br>Premenopausal p = 0.071, NS<br>Postmenopausal p=0.357, NS<br><br>CYP19A1 mRNA:<br>Premenopausal p = 0.511, NS<br>Postmenopausal p=0.585, NS |

| | | | |
|---|---|---|---|
| | STRAT PRE (all) POST | CYP19A1 (Quantitative RT-PCR Using TaqMan Low-Density Assay (TLDA) Microfluidic Cards)<br><br>E2 (LC-MS) breast tissue aspiration | Pearson correlation:<br>CYP19A1 mRNA:<br>Pre+postmenopausal , r=0.189, p=0.0463<br><br>Breast tissue E2 levels were higher in overweight women compared to normal-weight women in postmenopausal women (P< 0.05), but did not differ among all three groups of premenopausal women.<br><br>adjusted for age |
| Meza-Munoz 2006<br>(133)<br>Mexico<br>cross-sectional<br>Follow-up: months | Sample size n= 113<br>Postmenopausal: 54,<br>Premenopausal: 59<br><br>Volunteers pre- and postmenopausal healthy women,<br>BMI: < 27 (non-obese), >=30 (obese);<br>Age: premenopausal = 25–45, postmenopausal > 48<br><br>STRAT PRE (follicular) POST | Exposures<br>BMI (kg/m2)<br><br>Outcomes<br>Immunohistochemical (IHC) quantification:<br>ER-alpha<br>ER-beta<br>PR | Postmenopausal, BMI x PR, Beta=0.5, p=0.0003 (Bonferroni corrected=0.002)<br><br>Premenopausal women (follicular), p for difference between obese vs. non-obese<br>ER-a, NS<br>ER-b, NS<br>PR, higher non-obese, p < 0.006<br><br>Postmenopausal women, p for difference between obese vs. non-obese<br>ER-a, higher obese, <0.03<br>ER-b, higher obese, <0.02<br>PR, NS |
| Morris 2011<br>(134)<br>USA<br>cross-sectional<br>Follow-up: months | Sample size n= 30,<br>Premenopausal: 16,<br>Postmenopausal: 14<br><br>Pre- and postmenopausal women undergoing mastectomy for breast cancer treatment,<br>BMI: 24.5±4.7 (CLS-B negative), 31.6±6.4 (CLS-B positive);<br>Age: 48.6±11.8 (CLS-B negative), 51.4±8.7 (CLS-B positive)<br><br>PRE + POST Mix | Exposures<br>BMI (unit not reported), breast adipocyte size (size determined by Linear Dimensional Tool in Canvas 11, photographed at 20x with Olympus BX50 microscope and MicroFire digital camera)<br><br>Outcomes<br>Aromatase mRNA expression (Real-time PCR),<br><br>Aromatase activity (quantified by measurement of the tritiated water released from 1beta-[3H]androstenedione, femtomoles per microgram of protein per hour, ) | Weak to moderate Spearman rank correlation between<br>BMI*aromatase mRNA expression, rho=0.42, p=0.02;<br>BMI* aromatase activity, rho=0.5, p=0.02;<br><br>Aromatase activity: normalized to protein concentration |

| | | | |
|---|---|---|---|
| Daraei 2017<br>(135)<br>Iran<br>cross-sectional<br>Follow-up: months | Sample size n= 120,<br>normal weight: 43,<br>overweight: 51,<br>obese: 26<br><br>Healthy pre- (n=100) and postmenopausal (n=20) women with cosmetic mammoplasty, BMI: < 25 (normal), 25-29 (overweight), ≥ 30 (obese);<br>Age: not reported<br><br>PRE + POST Mix | Exposures<br>BMI (kg/m2)<br><br>Outcomes<br>ESR1 methylation (methylated DNAimmunoprecipitation-quantitative PCR) | Significant differences,<br>Higher methylation levels with higher BMI in promoter region, p<0.001 and Exon 1 region, p=0.023;<br><br>Beta -0.28 (SE=0.089) for association between BMI and -log2 transformed ESR1 methylation in promoter region<br><br>Not reported |
| Park 2017<br>(136)<br>USA<br>cross-sectional<br>Follow-up: months | Sample size n= 45<br><br>Pre- (n=23) and postmenopausal (n=22), normal weight women, BMI: 23.7±2.5;<br>Age: 35-65<br><br>PRE + POST Mix | Exposures<br>% body fat (%, not reported), Total Fat Mass (kg, dual-energy X-ray absorptiometry), Trunk Fat Mass (kg, dual-energy X-ray absorptiometry), Leg Fat Mass (kg, dual-energy X-ray absorptiometry), VFA (not reported, computed tomography), Abdominal SFA (cm2, computed tomography), Femoral SFA (cm2, computed tomography)<br><br>Outcomes<br>Estrogen receptor alpha - ESR1 (qPCR), estrogen receptor beta - ESR2 (qPCR) mRNA expression | Summary, correlation only significant for Total % fat and ERSR1:ESR2 measured in abdominal fat, r=−0.359, p<0.05;<br><br>%fat<br>ESR1 abdominal (a) r=-0.291, NS<br>ESR1 femoral (f) r=−0.219, NS<br>ESR2 a r=−0.028,p NS<br>ESR2 f r=−0.053, NS<br>ESR1:ESR2 a r=−0.359, p<=0.05<br>ESR1:ESR2 f r−0.211, NS<br><br>Total fat mass<br>ESR1 a r=−0.178 , NS<br>ESR1 f r=−0.160, NS<br>ESR2 a r=−0.139, NS<br>ESR2 f r=−0.061, NS<br>ESR1:ESR2 a r=−0.127, NS<br>ESR1:ESR2 f r=−0.156, NS<br><br>visceral fat area<br>ESR1 a r=−0.233 , NS<br>ESR1 f r=−0.248, NS<br>ESR2 a r=−0.129, NS<br>ESR2 f r=−0.055, NS<br>ESR1:ESR2 a r=−0.224, NS<br>ESR1:ESR2 f r=−0.229, NS<br><br>Abdominal subcutaneous fat area<br>ESR1 a r=−0.155 , NS<br>ESR1 f r=−0.179, NS<br>ESR2 a r=−0.038, NS<br>ESR2 f r=−0.032, NS<br>ESR1:ESR2 a r=−0.296, NS<br>ESR1:ESR2 f r=−0.250, NS<br><br>Femoral subcutaneous fat area |

| | | | |
|---|---|---|---|
| | | | ESR1 a r=0.022 , NS<br>ESR1 f r=0.030, NS<br>ESR2 a r=0.130, NS<br>ESR2 f r=−0.038, NS<br>ESR1:ESR2 a r=−0.071, NS<br>ESR1:ESR2 f r=0.042, NS<br><br>Trunk and Leg fat mass NS correlation as well<br><br>Analysis for age and ESR1 and ESR1:ESR2 were adjusted for either %fat, E2 or leptin |
| Shin 2007<br>(137)<br>Republic of Korea<br>cross-sectional<br>Follow-up: months | Sample size n= 43<br><br>Pre- (n=31) and postmenopausal (n=12) women scheduled for gynecologic surgery (total abdominal hysterectomy or myomectomy for non-inflammatory uterine leiomyoma or adenomyosis), BMI: premenopausal = 25.86±3.76, postmenopausal = 25.61±5.34; Age: premenopausal = 43.74±5.52, postmenopausal = 54.92±7.43<br><br>PRE + POST Mix | Exposures<br>BMI (kg/m2), waist circumference (cm), hip circumference (cm), thigh circumference (cm), arm circumference (cm), Waist-hip ratio<br><br>Outcomes<br>ER-a to ER-b semi-quantification (Real-time PCR)<br><br>Sc–Om ratio of the ER subtypes= ERa/ERB in abdominal subcutaneous adipose tissue by ERa/ERb in omental adipose tissue | Pearson correlation NS between ERa or ERb or ERa/ERb ratio in SAAT or VAT and anthropometric indices (data not shown).<br><br>However, when ERa/ERb ratio of SAAT compared to VAT (=Sc–Om ratio of the ER subtypes), the significant correlation with anthropometric indices:<br><br>BMI, r=0.801, p<0.05<br>WC, r=0.696, p<0.05<br>Hip circ, r=0.716, p<0.05<br>WHR, r=0.347, p<0.05<br>Thigh circ, r=0.661, p<0.05<br>Arm circ, r=0.409, p<0.05<br><br>Not reported |
| Iyengar 2021<br>(138)<br><br>cross-sectional<br>Follow-up: months | Sample size n= 100<br><br>women undergoing mastectomy as cancer treatment or risk reduction, median age 49 (range 29-82); 54% premenopausal; 46% postmenopausal (n=100)<br><br>PRE + POST Mix | Exposures<br>BMI, median (range) 26.1 (17.5–42.0)<br>Total Fat % 33.5 (21.7–43.8)<br>Trunk fat % 30.3 (18.2–48.1)<br>measured with DXA<br><br>Outcomes<br>Aromatase expression | Aromatase mRNA<br>BMI, r= 0.50, p<0.001<br>%total fat, r= 0.51,p<0.001<br>%trunk fat, r= 0.58, p<0.001 |
| Iyengar 2021<br>(139) | Sample size n= 141 (74 BRAC1 and 67 BRAC2 mutation)<br><br>BRCA1 and BRCA2 mutation | Exposures<br>BMI | Relative aromatase expression by BMI category<br>BRCA1: Higher expression with higher BMI, p<0.001<br>BRCA2: Higher expression with higher BMI, p<0.001 |

| | | | |
|---|---|---|---|
| cross-sectional<br>Follow-up: months | carriers, pre (61%) and postmenopausal (39%) women ages median 43 (range 37–50), 64% had invasive breast cancer, 46% had noninvasive/benign tumors<br><br>STRAT PRE POST | Outcomes<br>Aromatase expression | Premenopausal (BRAC1 and 2 ): Higher expression with higher BMI, p<0.001<br>Postmenopausal (BRAC1 and 2 ): Higher expression with higher BMI, p<0.001<br><br>Correlations between aromatase and adipocyte size<br>BRCA1, r=0.62, p<0.001<br>BRCA2, r=0.49, p<0.001 |
| Iyengar 2017<br>(140)<br>USA<br>cross-sectional<br>Follow-up: months | Sample size n= 72<br><br>normal weight, pre (n=75%) and postmenopausal (n=25%) women undergoing mastectomy for breast cancer risk reduction or treatment, BMI: < 25;<br>Age: 31-64 years (median age aprox. 48)<br><br>PRE + POST Mix | Exposures<br>BMI (kg/m2)<br><br>Outcomes<br>Aromatase expression (Real-time PCR) | Average adipocyte diameter positively correlates with breast aromatase mRNA level (r= 0.33; P < 0.01)<br><br>Higher BMI correlates with elevated aromatase mRNA levels in breast tissue (r = 0.28, P =0.02).<br><br>Multiple linear regression analysis were adjusted for potential batch effects |
| Argenta 2014<br>(141)<br>USA<br>Non-intervnetional trial<br>Follow-up: 12 months | Sample size n= No use of hormonal therapy: 43<br><br>Morbidly obese women who underwent Roux-en-Y gastric bypass surgery, We only extract data on women not on anti-estrogen therapy, BMI: 46 (36–64);<br>Age: 39 (20–60)<br><br>PRE + POST Mix<br>(Unknown menstrual cycle) | Exposures<br>Weight loss (kg)<br><br>Outcomes<br>ER - SP1- (H-scores, IHC - Antibody staining), PR -1E2- (H-scores, IHC - Antibody staining)<br><br>--> H-score (Histocores had a potential range of 0 (100% unstained) to 300 (100% intensely staining)) were assesed by 2 independent pathologists | Weight loss = 41kg (19 to 67)<br>t-test<br>ER protein level H-score<br>195 (80-295) to 173 (30-275), p=0.42<br><br>PR protein level H-score<br>223 (105-300) to 160 (50-284), p=0.01<br><br>Not reported |
| Morán 1996<br>(142)<br>Mexico<br>Cross-sectional<br>Follow-up: months | Sample size n= 30 women<br>(10 nonobese, 10 upper body segment obesity, 10 lower body obesity)<br><br>No information on women selection<br><br>BMI: nonobese: 23.1 (20.7-23.9)<br>Lower body obesity 27.8 (26.2-32.4) | Exposures<br>Upper body obesity WHR>0.85<br>Lower body obesity WHR<=0.85<br><br>Outcomes<br>Protein levels (fmol/mg protein) (method Lowry et al) | ER:<br>Upper body segment obesity had significantly greater ER than nonobese women ((49.3 (15.8-109.0) vs. 12.9 (2.2-68.8), p<0.05).<br>Upper vs lower ((49.3 (15.8-109.0) vs.19.9 (1.6-75.7), NS)<br><br>PR<br>NS between the 3 groups (nonobese 21.2 (11.8-46.7) vs. upper 37.7 (3.6-103.0) vs. lower 16.7 (4.1-92.7), NS) |

| | | | |
|---|---|---|---|
| | Upper body obesity 30.2 (25.2-34.8)<br>Age: 20-35y<br><br>No information (Assumed premenopausal women but no menstrual cycle phase reported) | | |
| Laforest 2019<br>(143)<br>Canada<br>Cross-sectional<br>Follow-up: months | Sample size n= 23<br><br>23 women total: 17 breast cancer cases (partial/total mastectomy) and 6 controls (reduction mammoplasty);<br>median age 55 years (IQR 50.1–62.9);<br>pre- and postmenopausal women included (35% premenopausal, 65% postmenopausal);<br>median BMI 25.6 kg/m² (overweight);<br><br>PRE + POST Mix | Exposures<br>BMI (kg/m²)<br><br>Outcomes<br>Relative adipose tissue steroid amounts (pmol/kg, LC-MS/MS): cortisone, cortisol, estrone (E1), estradiol (E2); calculated ratios (E2:E1 as 17β-HSD activity marker)<br>gene expression (qPCR, relative mRNA) of CYP19A1, ESR1, ESR2 | E2:E1 ratio higher in lean vs. BMI≥25 women (P=.0335)<br>Higher CYP19A1 expression in higher-BMI women;<br>E2 amount higher in lean vs. overweight/obese cancer cases (P=.0494); |
| Angela Cho 2022<br>(144)<br>USA<br>Cross-sectional<br>Follow-up: months | Sample size n= 32 normal BMI women<br><br>high vs. low BMI (median split, ≥21.8 vs. <21.8 kg/m²)<br><br>32 normal-BMI (18.5–<25 kg/m²) women who underwent mastectomy for breast cancer treatment or risk reduction; median age 44.5 years; 67.7% White, 19.4% Asian, 6.5% Black, 6.5% other; 75% had invasive disease; 20% postmenopausal<br><br>PRE + POST Mix | Exposures<br>Trunk fat percentage by DXA (dual-energy x-ray absorptiometry)<br><br>Outcomes<br>RNA-seq gene expression<br><br>A total of 226 upregulated and 137 downregulated differentially expressed genes (DEGs) (P.adj <0.05, \|Log2FoldChange\| > 0.6 ) were found in high compared to low trunk fat women using DESeq2 | Spearman's method<br>Nearly significant positive correlations between trunk fat % and levels of expression of CYP19A1/aromatase gene (log2FC=0.95, rho=0.33, P=0.07).<br><br>Wilcoxon:<br>Levels of CYP19A1 which encodes aromatase were increased in association with high vs. low levels of trunk fat (log2FoldChange = 0.95; P = 0.01).<br><br>unadjusted |
| Wagner 2022<br>(145)<br>Germany<br>cross-sectional | Sample size n= 20<br><br>n=10, BMI 22.3±2.3 kg/m², body weight 57.3±6.1 kg) vs. obese women (n=10, BMI 50.5±7.7 | Exposures<br>Unknown how leana dn obses are defined<br><br>Outcomes | t-tests/Mann-Whitney<br>In adipose tissue: testosterone was 7.9-fold higher in visceral AT of obese vs. lean women (p=0.032, only significant AT androgen difference (NS -- DHT, androstenedione, progesterone, DHEA-S); CYP19/aromatase expression 4.1-fold higher in visceral AT of obese vs. lean (p=0.012); |

| | | |
|---|---|---|
| bariatric surgery patients, baseline vs. 12 months post-surgery Follow-up: Lean and obese months | kg/m², body weight 136.4±14.2 kg);<br><br>PRE + POST Mix | AT androgens (testosterone, DHT, androstenedione, progesterone, DHEA-S) via LC-MS/MS (pmol/mg AT or pmol/L in supernatant);<br><br>mRNA expression CYP19/Aromatase |

### Figures

| | CUP level of evidence | Description | Positive association | Inverse association |
|---|---|---|---|---|
| | Convincing | **Convincing causal**<br>-Significant association from more than one study type (RCT, cohort) or pooled estimates from RCT or cohort<br>-At least two independent cohorts with same results or their significant meta-analysis<br>-No unexplained heterogeneity<br>-No substantial publication biases<br>-Good quality studies<br>-Plausible gradient dose-response<br>-Experimental studies human or animal | *Estrone (Postmenopausal women) (lifestyle intervention, I2=20%; observational I2=20%)* | |
| **Strong evidence** | Probable | **Probable causal**<br>-At least two independent cohorts both same direction or their significant meta-analysis<br>-No unexplained heterogeneity<br>-No substantial publication biases<br>-Good quality study to exclude bias<br>-Biological plausibility | Estradiol (Postmenopausal women) (lifestyle intervention, I2=84%)<br><br>Free estradiol (Postmenopausal women) (lifestyle intervention, I2=84%)<br><br>Testosterone (pre and postmenopausal) (surgery intervention, I2=51%; lifestyle intervention, I2=45%; observational, I2=0 100%)<br><br>Free testosterone (postmenopausal?) (surgery intervention, I2=27%; lifestyle intervention, I2=69%; observational, I2=0 29%) | SHBG (pre and postmenopausal, longer FU, higher loss fat, VAAT) (surgery intervention, I2=51%; lifestyle intervention, I2=74%; observational, I2=0%) |
| | Substantial effect unlikely | **Causal unlikely**<br>-Evidence from more than 1 study type (RCT, cohort, pooled)<br>-At least two independent cohort studies<br>-Summary estimate close to 1.0 for comparison of high vs. low exposure<br>-No unexplained heterogeneity<br>-Good quality study to exclude bias<br>-Absence of biological gradient (dose-response) and absence of plausible experimental evidence | Androstenedione (significant in surgerical intervention I2=59%; lifestyle intervention, I2=0% borderline significant (postmenopausal), observational not significant)<br><br>DHEAS<br><br>Estradiol (Premenopausal women) | |
| | Limited-Suggestive | **Limited suggesting causal**<br>(Borderline or not significant but toward one direction)<br>-At least two independent cohorts both same direction<br>-The direction of effect is generally consistent though some unexplained heterogeneity may be present.<br>-There may be moderate or high risk of bias in some studies<br>-There may be moderate or high risk of publication bias<br>-Evidence of biological plausibility. | Aromatase (Tissue-based)<br><br>PR (postmenopausal women) | ERα (Tissue-based)<br><br>PR (Tissue-based, premenopausal women follicular) |
| **Limited evidence** | Limited- No conclusion | No firm conclusion can be made because of limited evidence | Progesterone or pregnanediol glucuronide<br><br>Free estradiol (premenopausal women)<br><br>Estrone (Premenopausal women)<br><br>Estrone glucuronide (pre and postnmenopausal women)<br><br>Estrogen metabolite (pre and postnmenopausal women)<br><br>Free testosterone (premenopausal)<br><br>DHEA<br><br>Androstenediol , androsterone glucuronide , androsterone<br><br>Group of estrogen, estriol, dihydrotestosterone, free androgen index, progesterone | |

Fig. S1: Strength of evidence for the associations between body fatness and sex steroid hormones (adapted from CUP program (146)

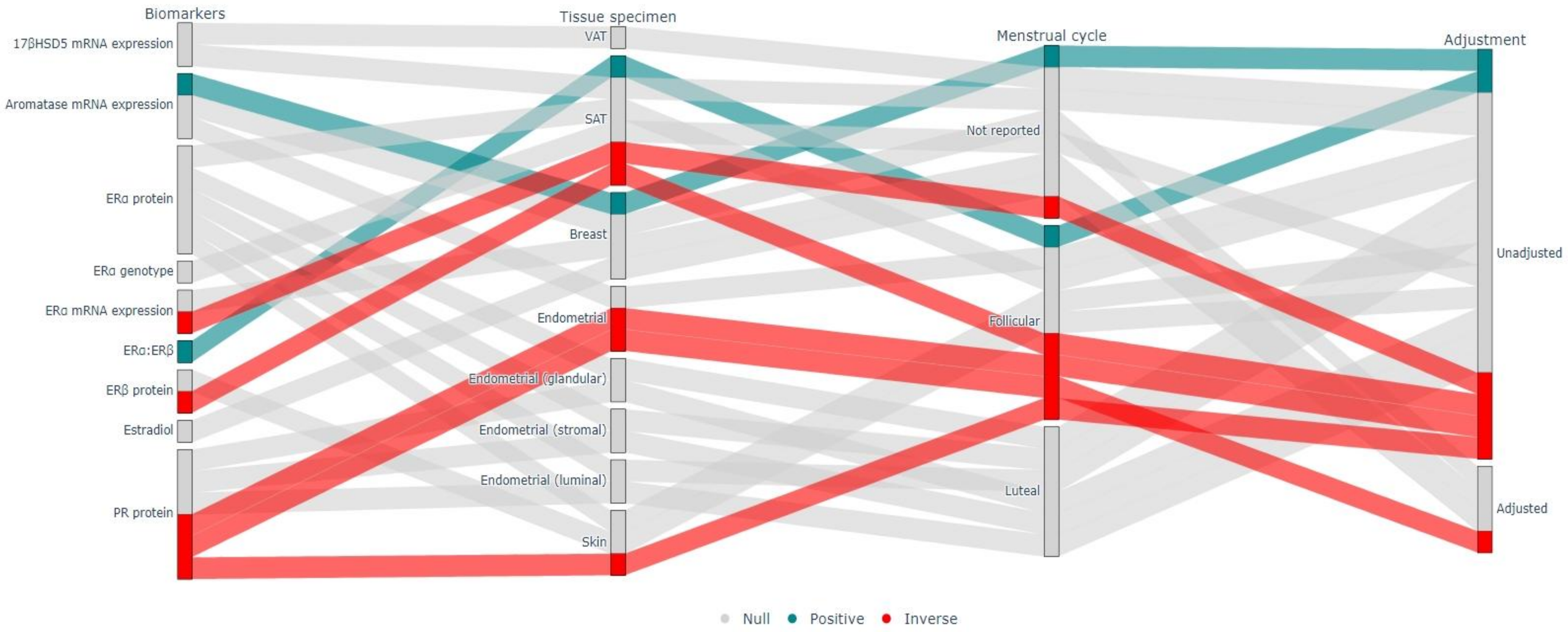

Fig. S2: Sanky plot on associations between body fatness and sex steroids measured in tissue, premenopausal women.

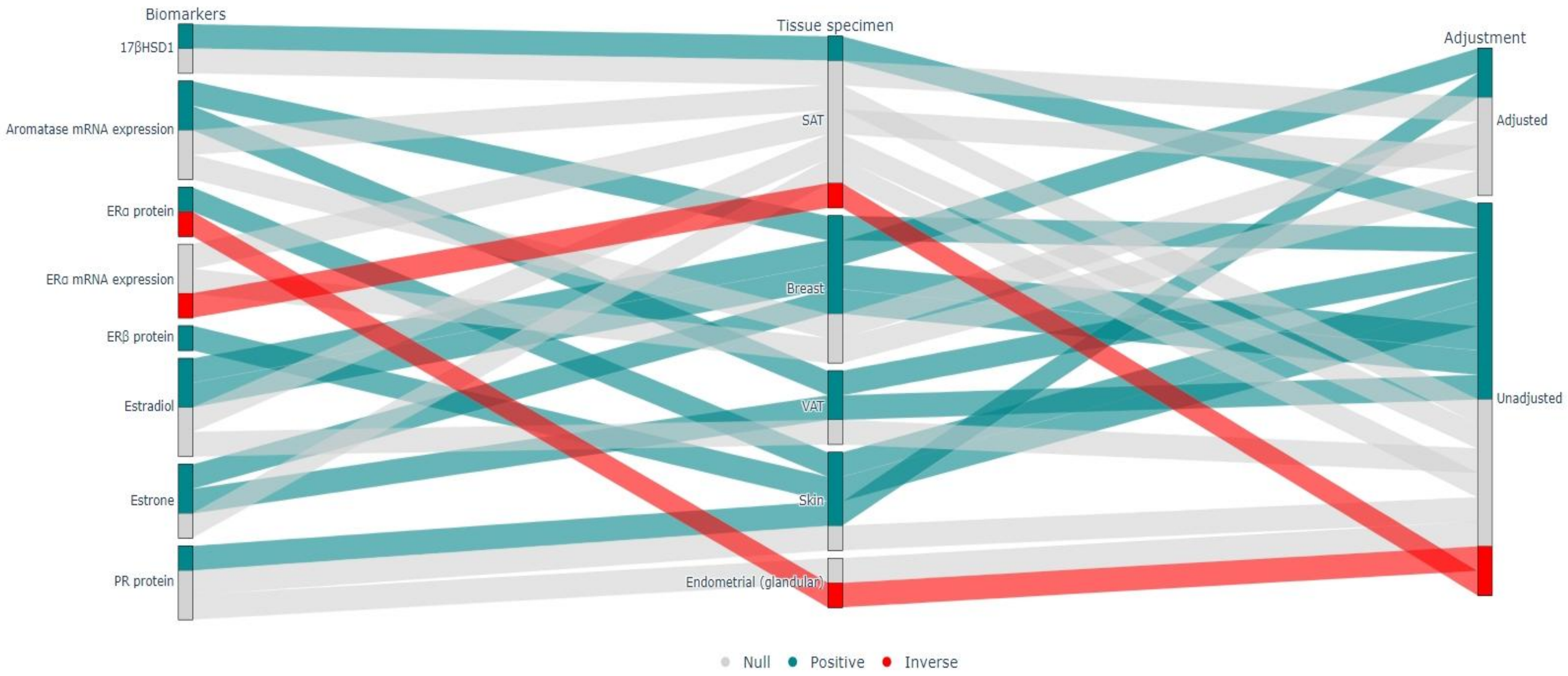

Fig. S3: Sanky plot on associations between body fatness and sex steroids measured in tissue, postmenopausal women.

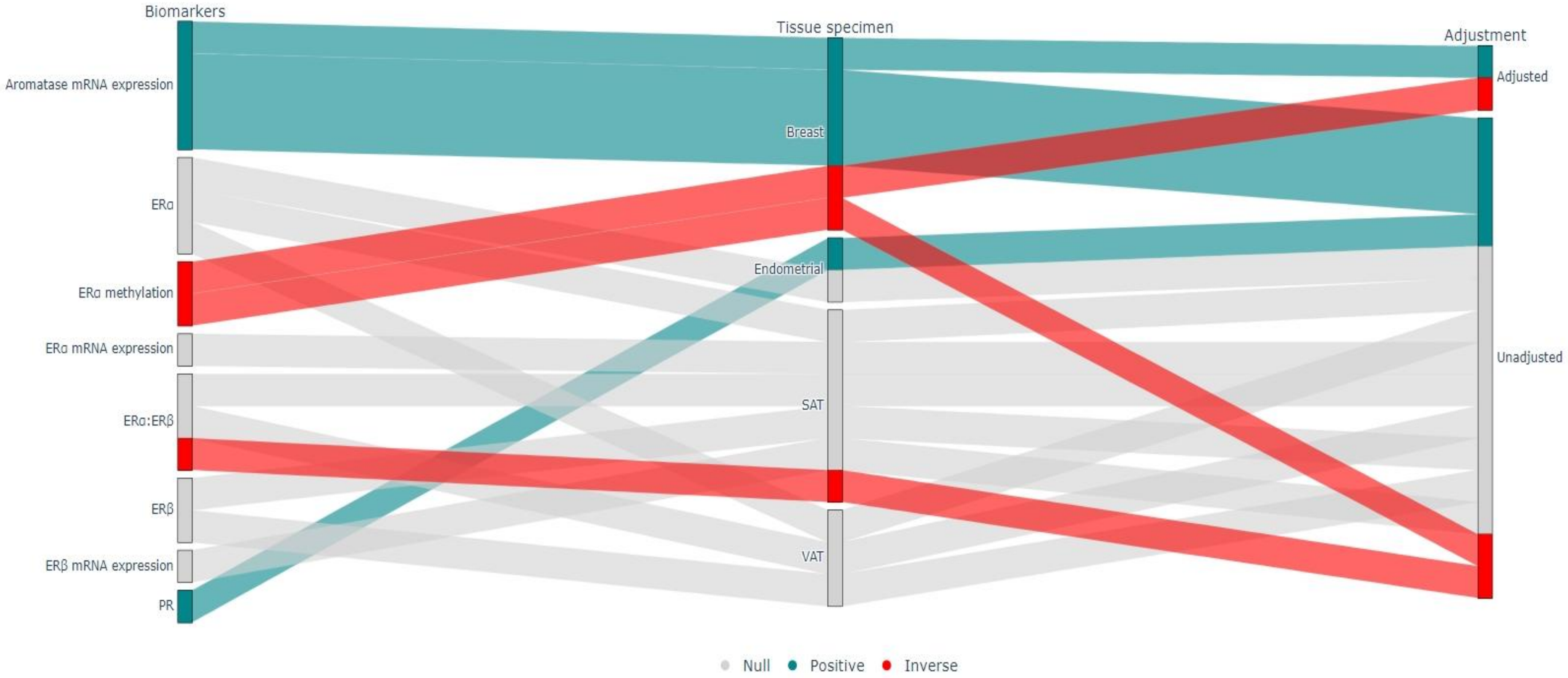

Fig. S4: Sanky plot on associations between body fatness and sex steroids measured in tissue, pre- and postmenopausal women.

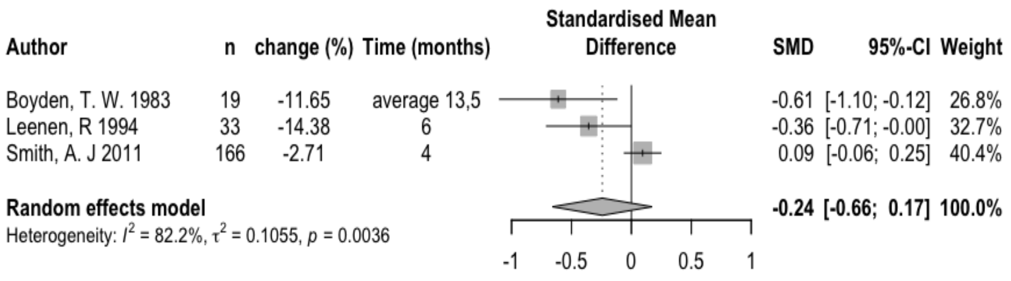

Fig. S5. Premenopausal estradiol - meta-analysis of lifestyle intervention studies

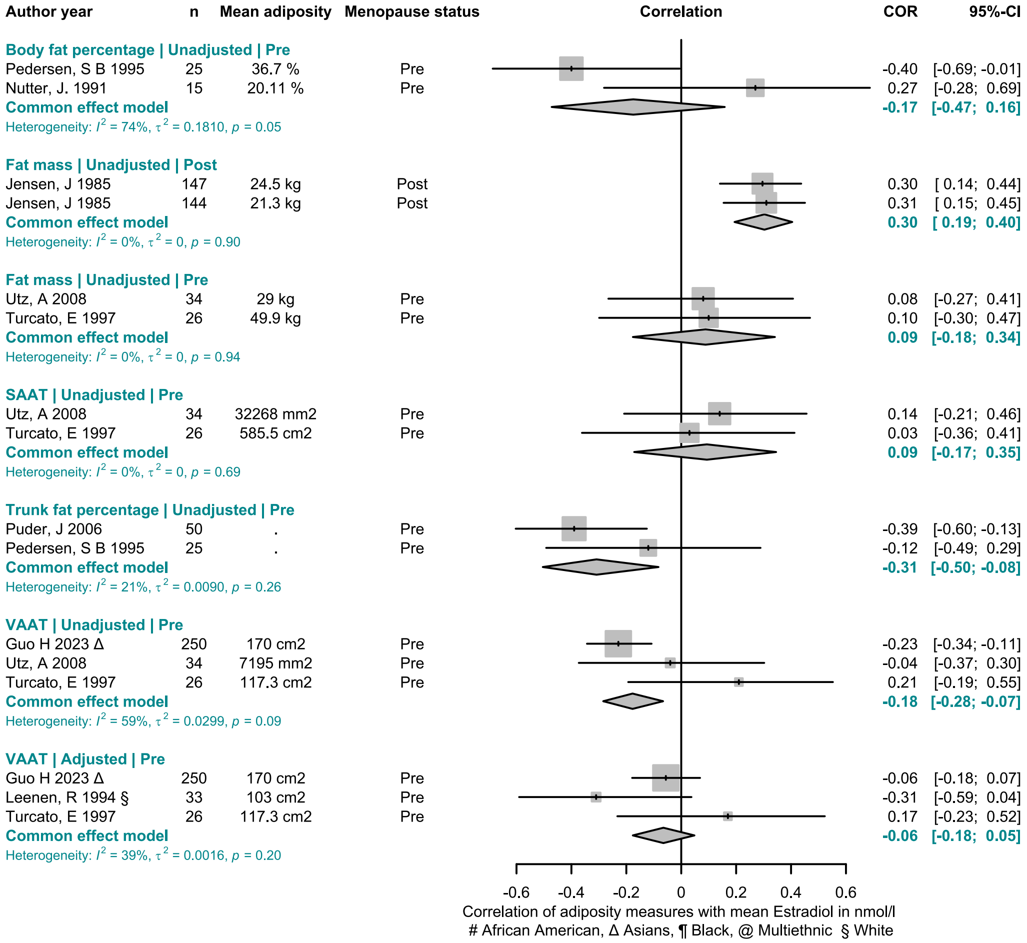

Fig. S6. Premenopausal estradiol - meta-analysis of correlations with different adiposity measures.

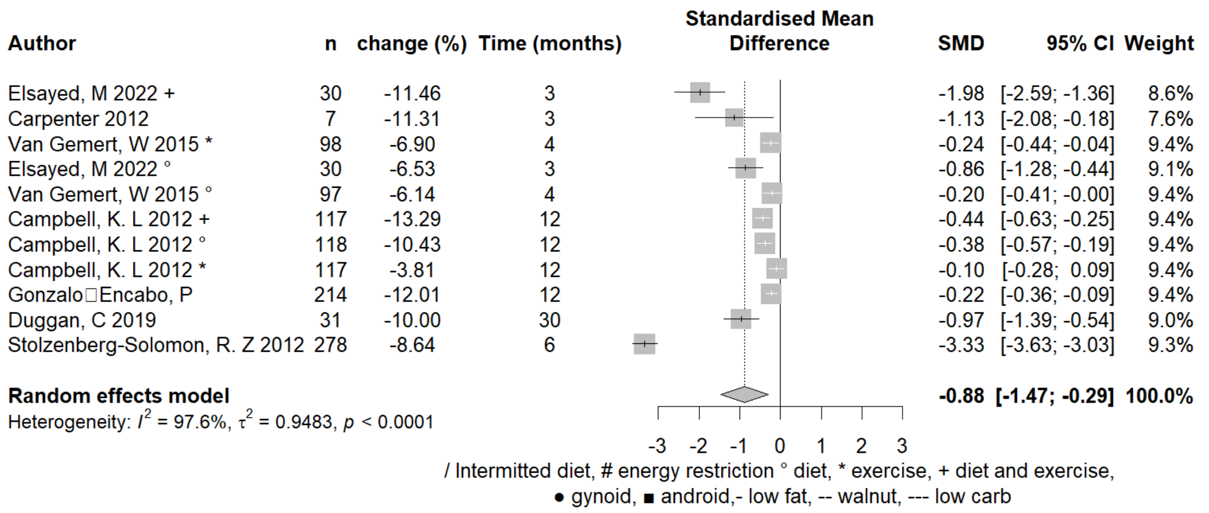

Fig. S7. Postmenopausal estradiol - meta-analysis of lifestyle intervention studies

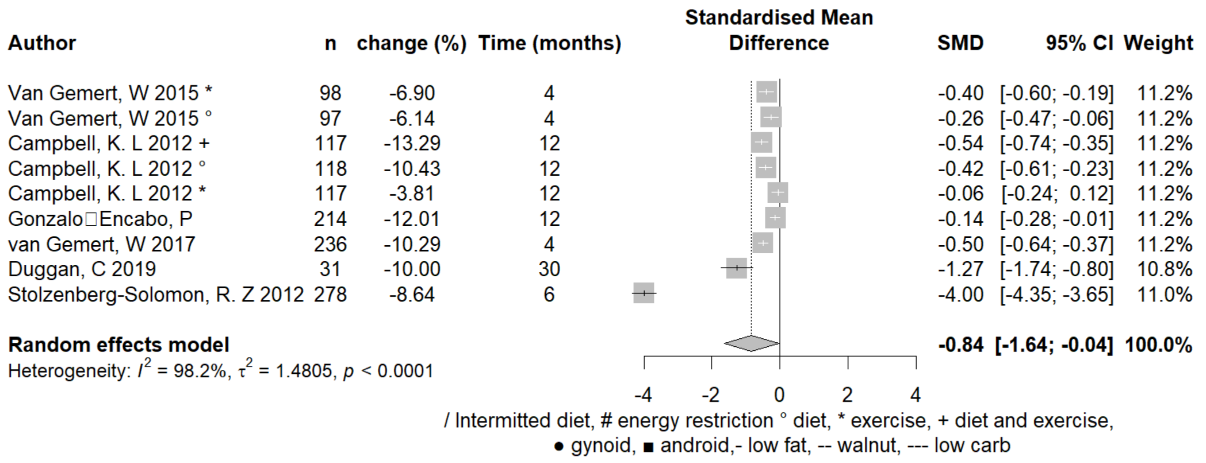

Fig. S8 Postmenopausal free estradiol - meta-analysis of lifestyle intervention studies.

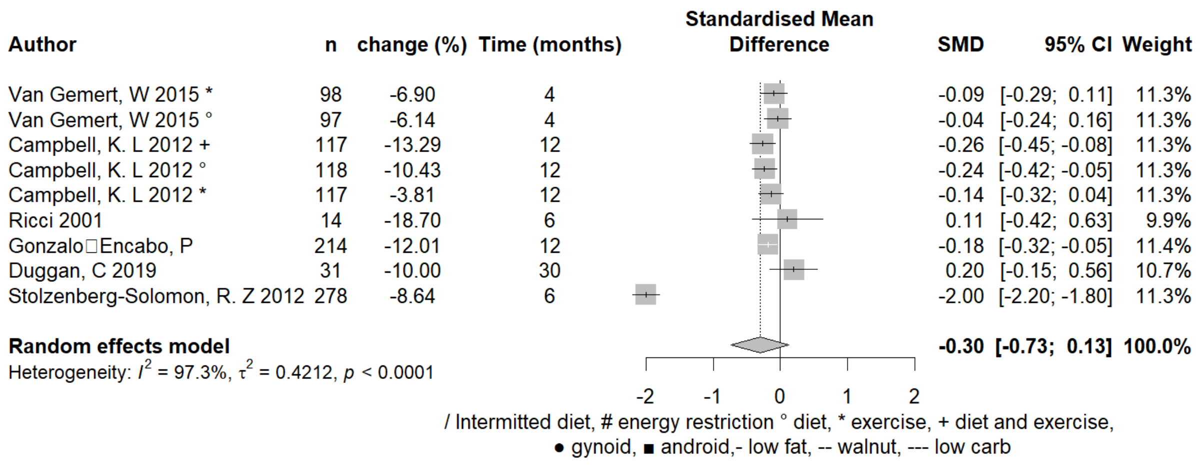

Fig. S9. Postmenopausal estrone - meta-analysis of lifestyle intervention studies.

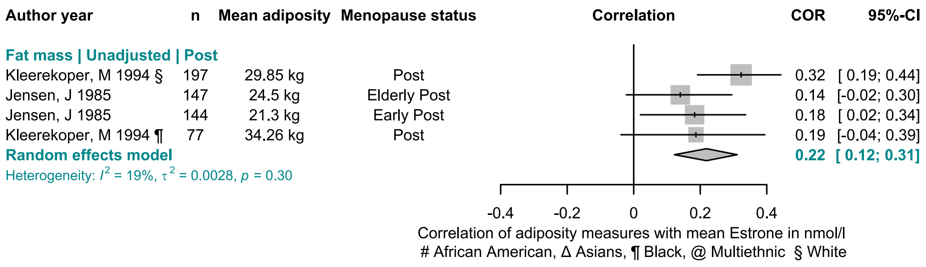

Fig. S10. Postmenopausal estrone - meta-analysis of unadjusted correlations with fat mass.

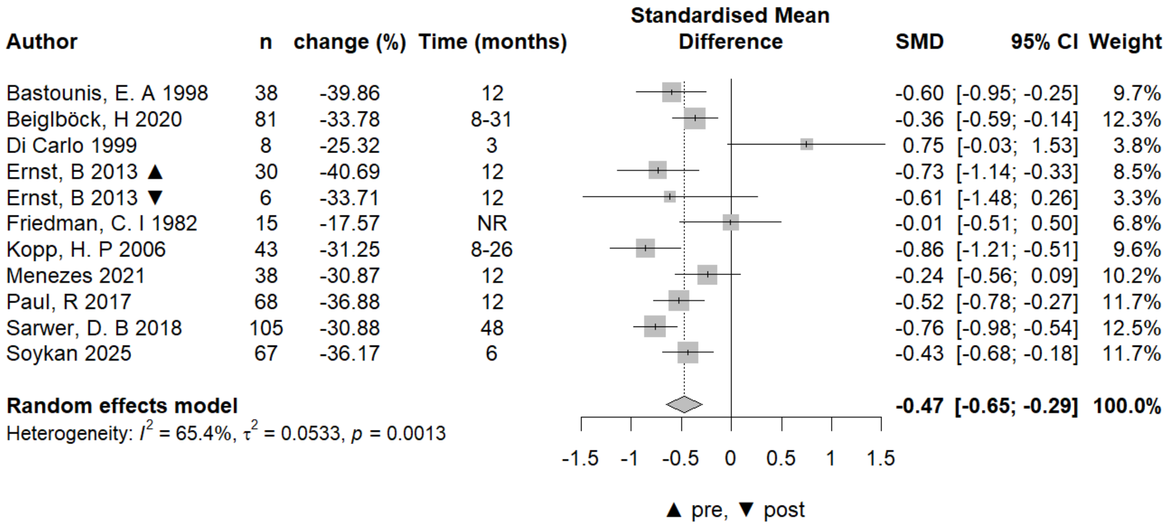

Fig. S11. Circulating testosterone - meta-analysis of surgical interventions.

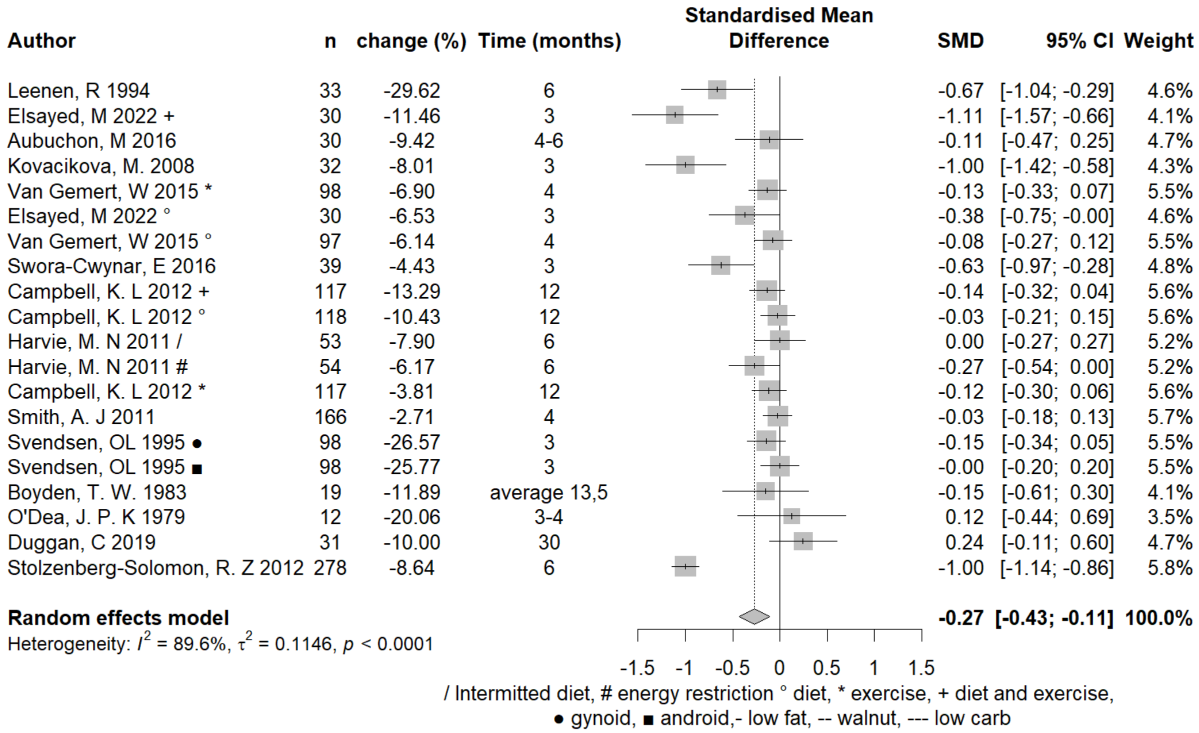

Fig. S12. Circulating testosterone - meta-analysis of lifestyle interventions.

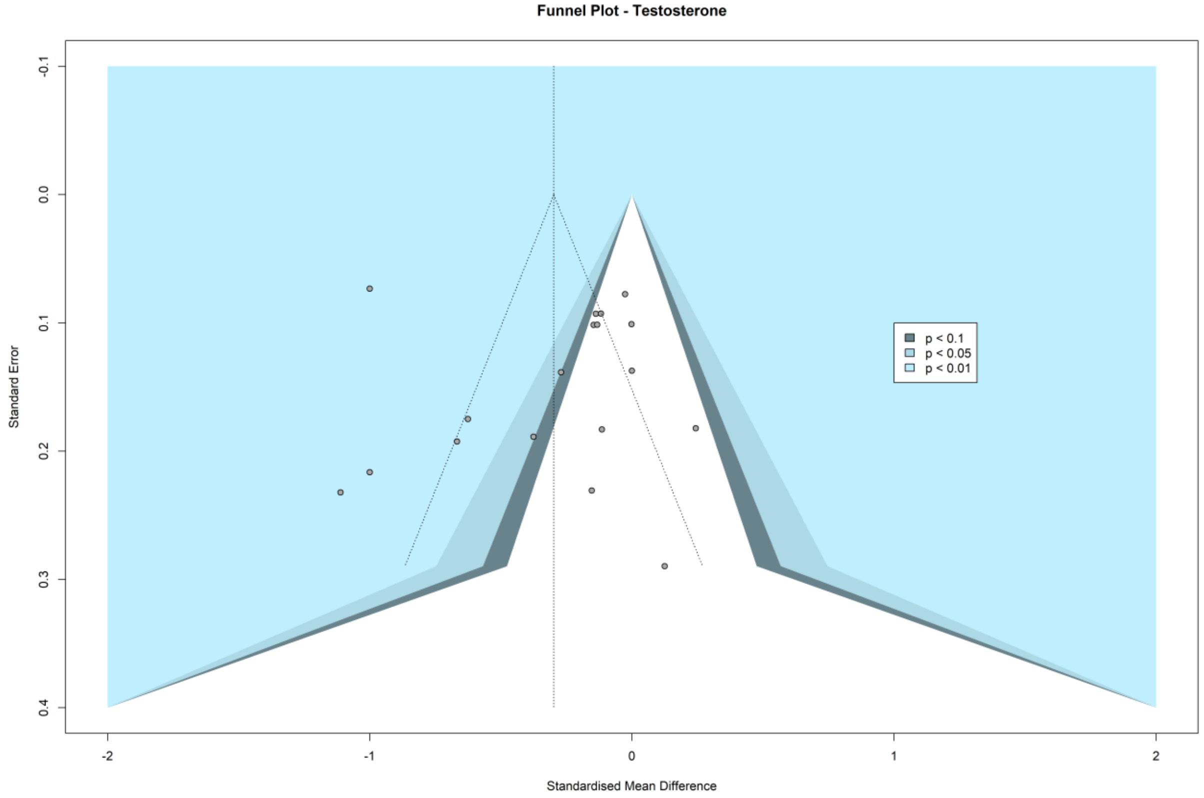

Fig. S13. Circulating testosterone -. Funnel plot of lifestyle intervention studies.

| Author year | Adiposity measurement | Mean adiposity | Menopausal status | N | COR | 95%-CI |
|---|---|---|---|---|---|---|
| **Body fat percentage \| Unadjusted** | | | | | | |
| Lee, C 2004 | DEXA | 39 % | Post | 34 | -0.03 | [-0.36; 0.31] |
| Casson, P 2010 | DEXA | 37.9 % | Post | 29 | -0.38 | [-0.65; -0.01] |
| Marchand, G 2018 | DEXA | 35.3 % | Pre | 42 | -0.06 | [-0.36; 0.25] |
| Ofori, E 2019 | DEXA | 33.53 % | Post | 35 | 0.38 | [ 0.05; 0.63] |
| Pedersen, S B 1995 | DEXA | 36.7 % | Pre | 25 | -0.09 | [-0.47; 0.32] |
| **Random effects model** | | | | | **-0.03** | **[-0.27; 0.22]** |
| Heterogeneity: $I^2$ = 57.7%, $\tau^2$ = 0.0465, $p$ = 0.0505 | | | | | | |
| **Body fat percentage \| Adjusted** | | | | | | |
| Keller, J 2011 | DEXA | 31.6 % | Pre | 30 | 0.35 | [-0.01; 0.63] |
| **Central fat \| Unadjusted** | | | | | | |
| Lee, C 2004 | DEXA | 2545 g | Post | 34 | -0.02 | [-0.36; 0.32] |
| **Fat mass \| Unadjusted** | | | | | | |
| Utz, A 2008 | DEXA | 29 kg | Pre | 34 | 0.20 | [-0.15; 0.50] |
| Casson, P 2010 | DEXA | 26.3 kg | Post | 29 | -0.33 | [-0.62; 0.04] |
| Peiris, A N 1989 | Hydrostatic weighing | 35.9 kg | Pre | 31 | -0.26 | [-0.56; 0.10] |
| Ofori, E 2019 | DEXA | 24.36 kg | Post | 35 | 0.42 | [ 0.10; 0.66] |
| Armellini, F 1994 | CT | 52 kg | Pre | 36 | -0.23 | [-0.52; 0.11] |
| Turcato, E 1997 | CT | 49.9 kg | Pre | 26 | -0.01 | [-0.40; 0.38] |
| Turcato, E 1997 | CT | 44 kg | Post | 15 | -0.10 | [-0.58; 0.43] |
| **Random effects model** | | | | | **-0.04** | **[-0.26; 0.19]** |
| Heterogeneity: $I^2$ = 58.7%, $\tau^2$ = 0.0533, $p$ = 0.0243 | | | | | | |
| **Fat mass \| Adjusted** | | | | | | |
| Phillips, G 2008 | MRI | 22.3 kg | Pre | 58 | 0.22 | [-0.04; 0.45] |
| Phillips, G 2008 | MRI | 32 kg | Post | 20 | 0.43 | [-0.02; 0.73] |
| Keller, J 2011 | DEXA | 20.1 kg | Pre | 30 | 0.38 | [ 0.02; 0.65] |
| **Random effects model** | | | | | **0.30** | **[ 0.11; 0.47]** |
| Heterogeneity: $I^2$ = 0%, $\tau^2$ = 0, $p$ = 0.6034 | | | | | | |
| **IAAT \| Unadjusted** | | | | | | |
| Kunesová , M 2002 | CT | 195.9 cm2 | Both | 94 | -0.17 | [-0.36; 0.03] |
| Utz, A 2008 | CT | 48393 mm2 | Pre | 34 | 0.11 | [-0.24; 0.43] |
| **Random effects model** | | | | | **-0.07** | **[-0.32; 0.20]** |
| Heterogeneity: $I^2$ = 45.7%, $\tau^2$ = 0.0182, $p$ = 0.1749 | | | | | | |
| **IAAT thickness \| Unadjusted** | | | | | | |
| De Pergola, G 1994 | Sonography | 56.6 mm | Pre | 40 | -0.32 | [-0.58; -0.01] |
| **Percent Android fat \| Adjusted** | | | | | | |
| Cao, Y 2013 | DEXA | . | Early Post | 105 | -0.03 | [-0.22; 0.17] |
| Cao, Y 2013 | DEXA | . | Late post | 107 | 0.12 | [-0.07; 0.30] |
| **Random effects model** | | | | | **0.05** | **[-0.09; 0.19]** |
| Heterogeneity: $I^2$ = 8.4%, $\tau^2$ = 0.0009, $p$ = 0.2962 | | | | | | |
| **Percent Gynoid fat \| Adjusted** | | | | | | |
| Cao, Y 2013 | DEXA | . | Early Post | 105 | 0.02 | [-0.18; 0.21] |
| Cao, Y 2013 | DEXA | . | Late post | 107 | -0.11 | [-0.29; 0.08] |
| **Random effects model** | | | | | **-0.05** | **[-0.18; 0.09]** |
| Heterogeneity: $I^2$ = 0%, $\tau^2$ = 0, $p$ = 0.3642 | | | | | | |
| **SAAT \| Unadjusted** | | | | | | |
| Kunesová , M 2002 | CT | 471.8 cm2 | Both | 94 | 0.27 | [ 0.07; 0.45] |
| Utz, A 2008 | CT | 32268 mm2 | Pre | 34 | 0.20 | [-0.15; 0.50] |
| Casson, P 2010 | CT | 291.1 cm2 | Post | 29 | -0.06 | [-0.42; 0.31] |
| Marchand, G 2018 | CT | . | Pre | 41 | -0.08 | [-0.38; 0.23] |
| Ofori, E 2019 | MRI | 9.03 L | Post | 35 | 0.37 | [ 0.04; 0.63] |
| Zamboni, M 1994 | CT | 527 cm2 | Pre | 19 | 0.48 | [ 0.03; 0.77] |
| Cigolini, M 1996 | CT | 220 cm2 | Pre | 18 | 0.20 | [-0.29; 0.61] |
| Turcato, E 1997 | CT | 585.5 cm2 | Pre | 26 | 0.14 | [-0.26; 0.50] |
| Turcato, E 1997 | CT | 436.1 cm2 | Post | 15 | -0.13 | [-0.60; 0.41] |
| **Random effects model** | | | | | **0.17** | **[ 0.04; 0.30]** |
| Heterogeneity: $I^2$ = 15.6%, $\tau^2$ = 0.0069, $p$ = 0.3040 | | | | | | |

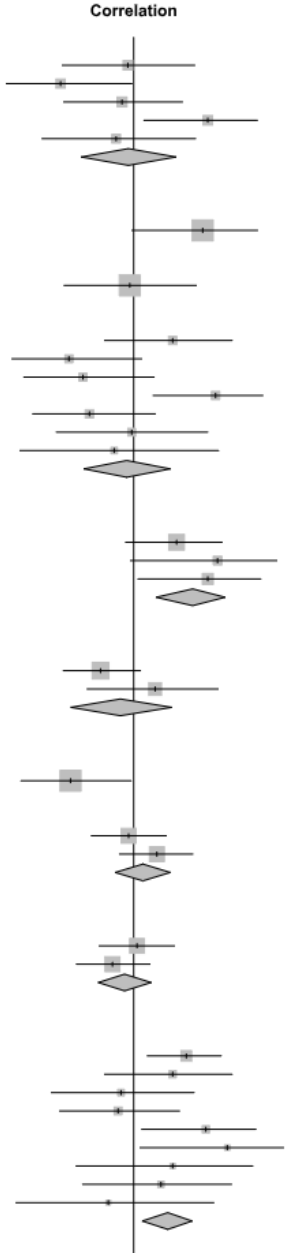

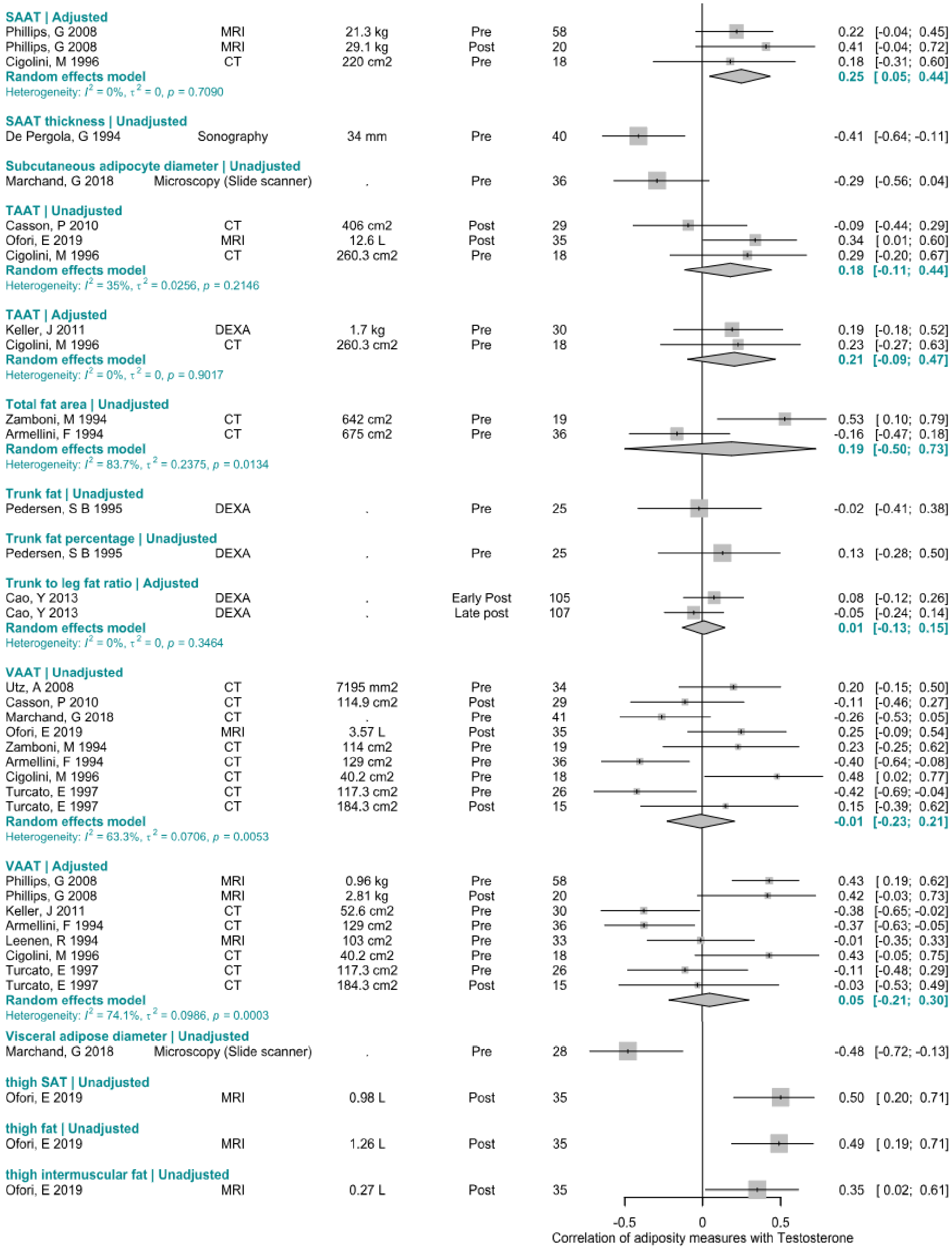

Fig. S14. Circulating testosterone - Forest plot for the correlation with direct adiposity measures

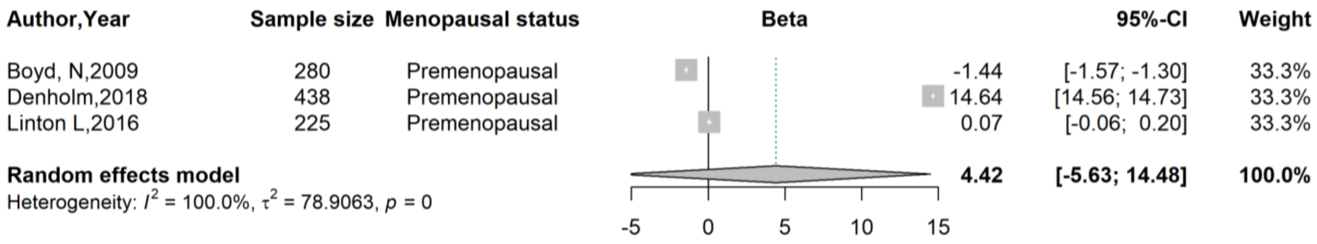

Fig. S15. Circulating testosterone - Meta-analysis of standardized beta coefficients from adjusted linear regression for the association with breast fat volume.

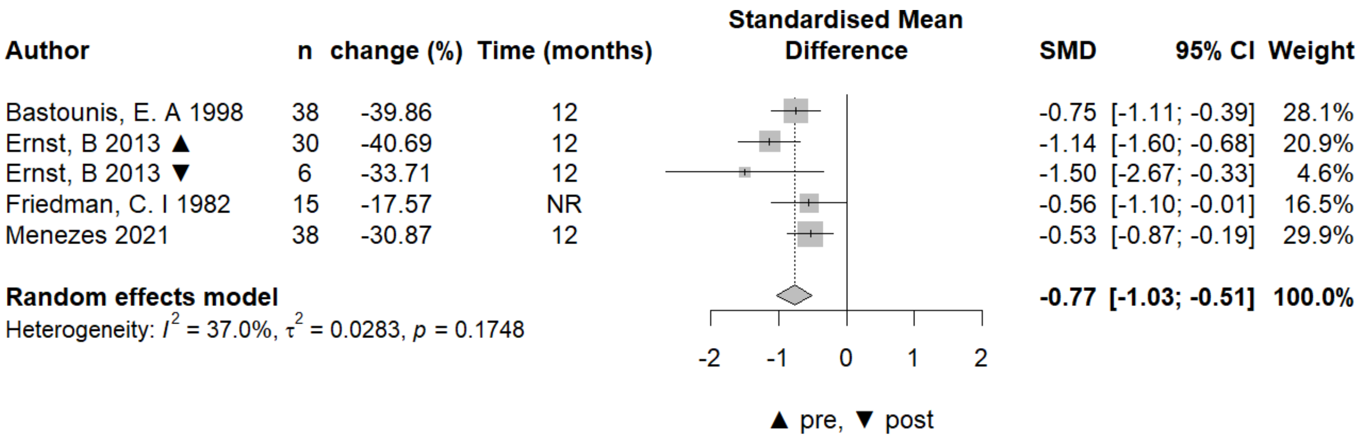

Fig. S16. Circulating free testosterone - Meta-analysis of -surgical interventions.

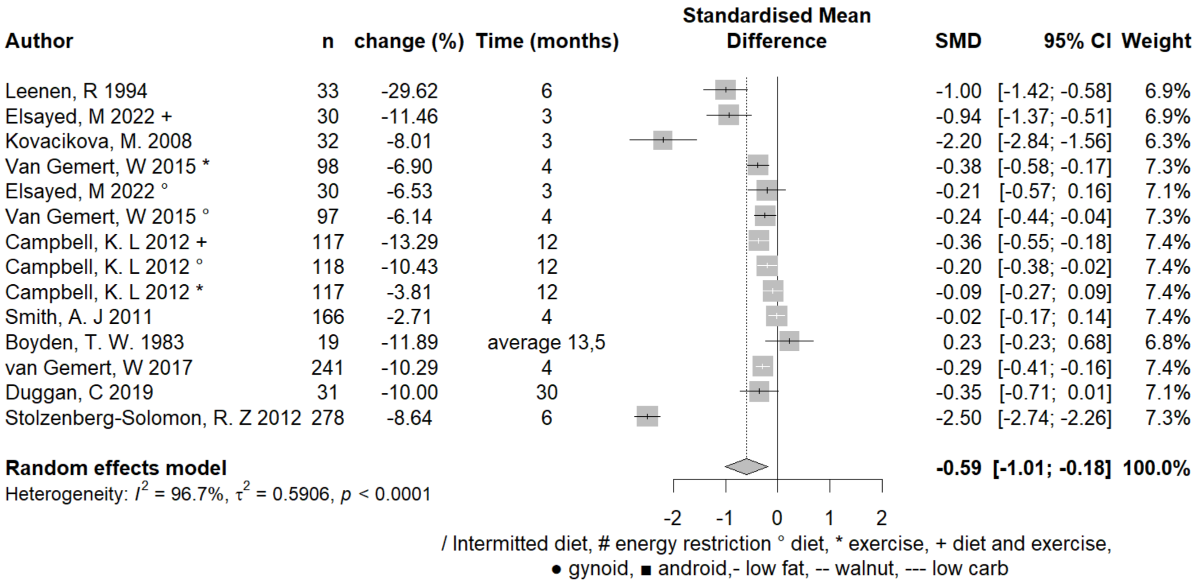

Fig. S17. Circulating free testosterone - Meta-analysis of - lifestyle interventions.

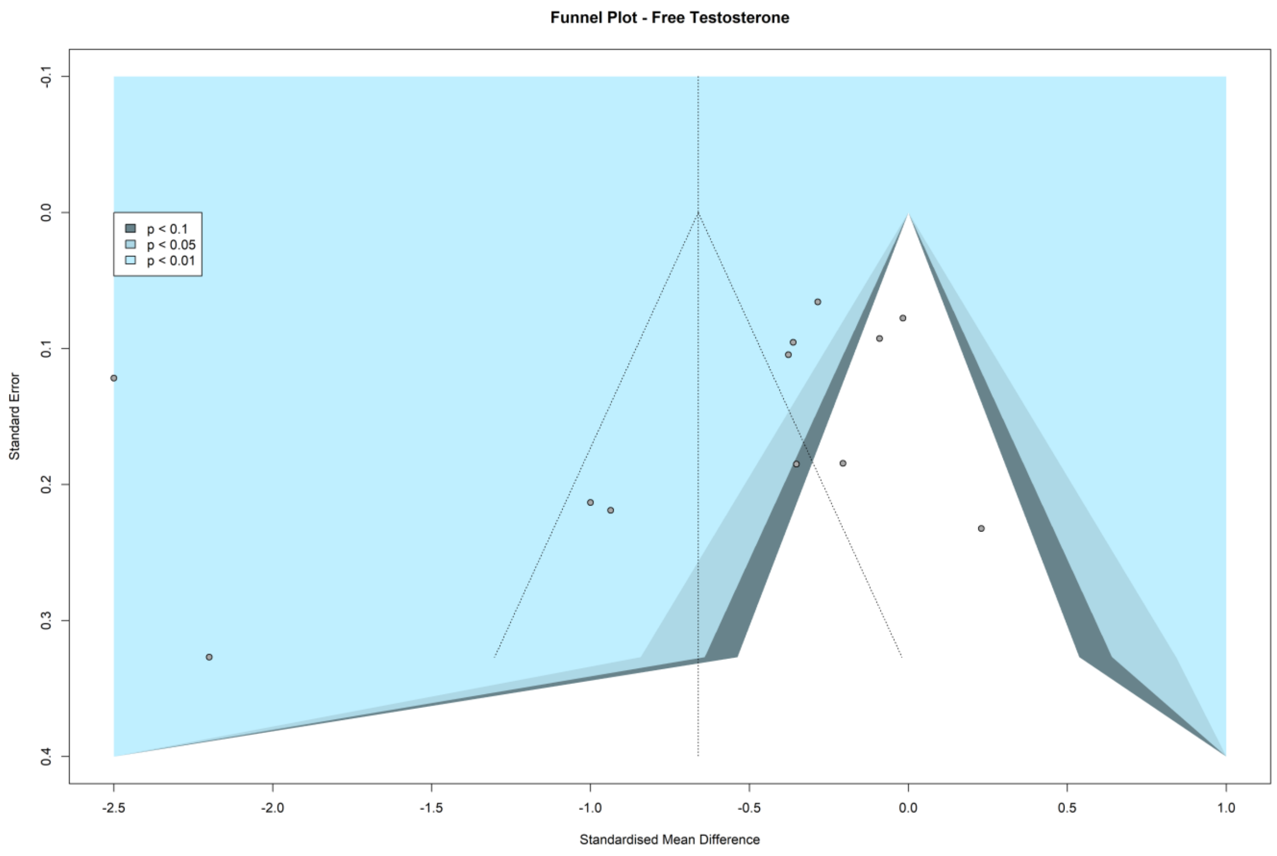

Fig. S18. *C*irculating free testosterone - Funnel plot of lifestyle intervention studies.

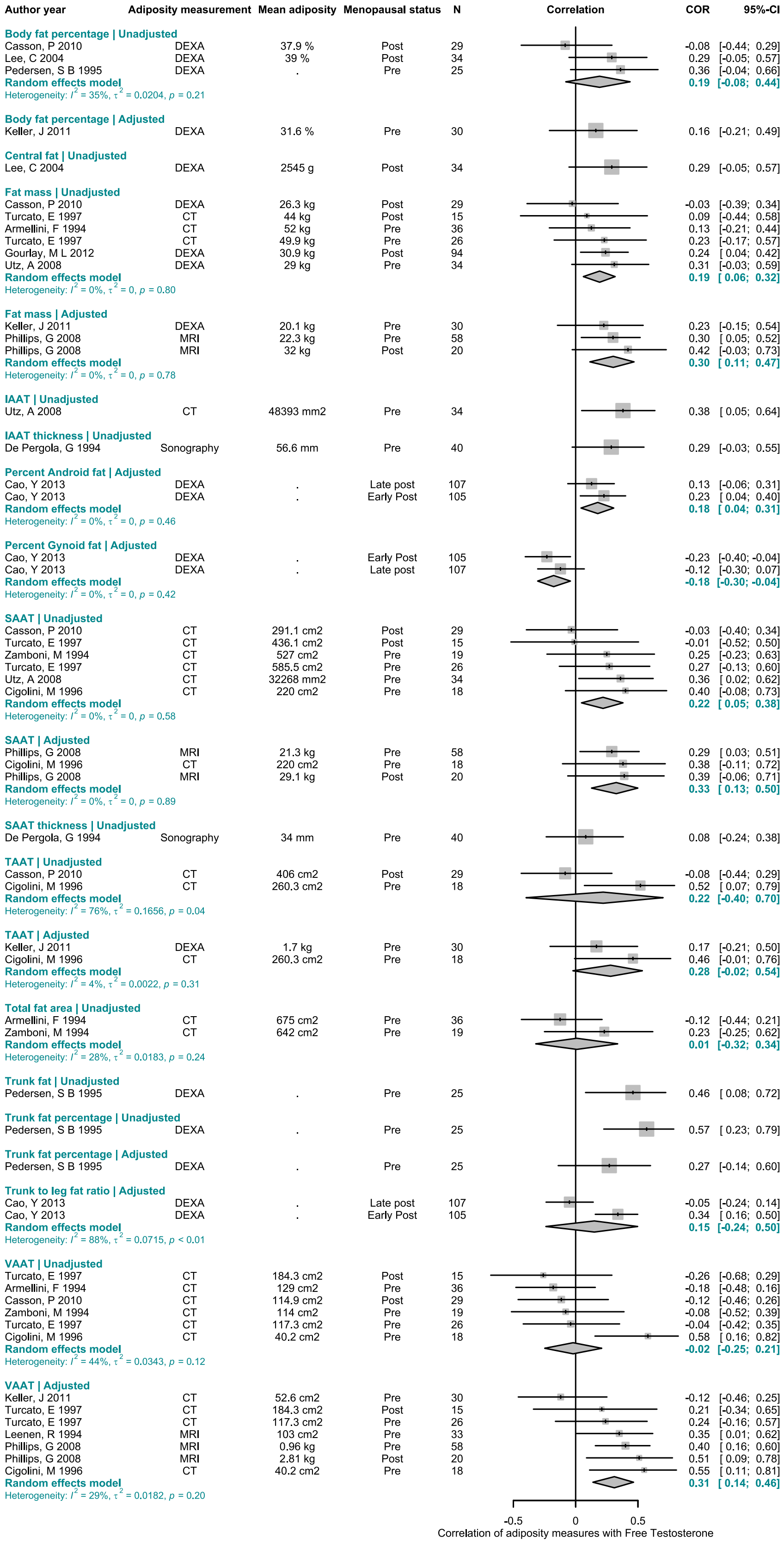

Fig. S19. Circulating free testosterone – *Meta-analysis of* correlations with direct adiposity measures.

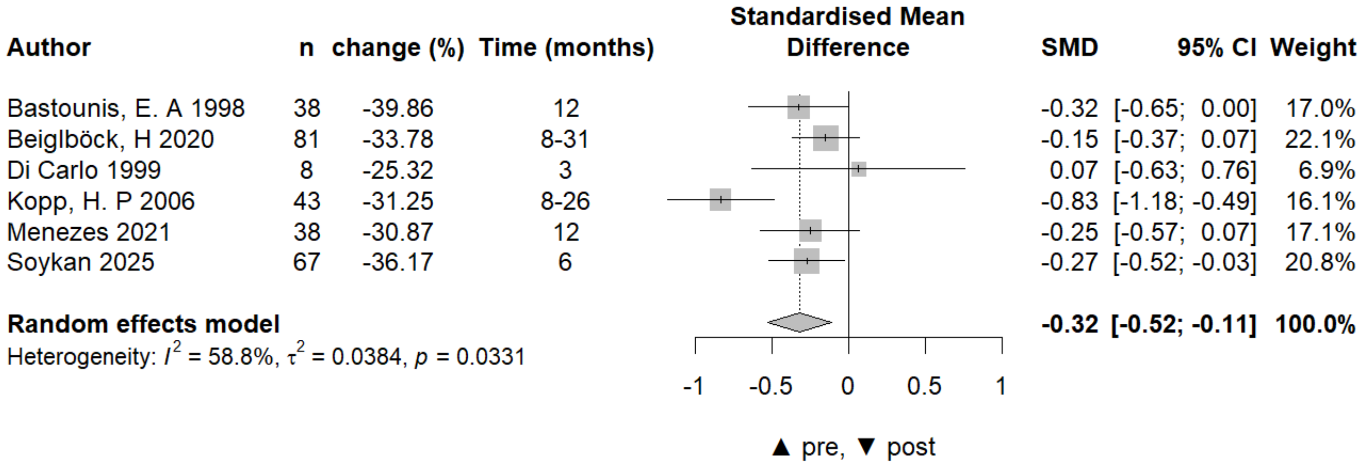

Fig. S20. Circulating androstenedione – Meta-analysis of surgical intervention studies.

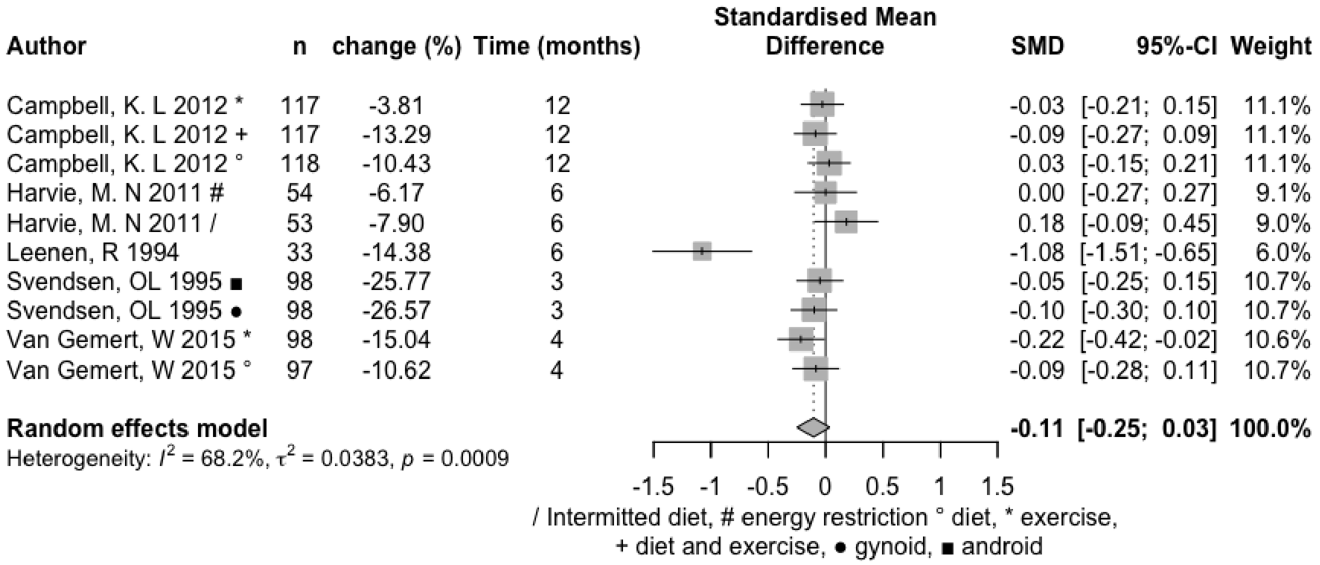

Fig. S21. Circulating androstenedione – Meta-analysis of lifestyle intervention studies.

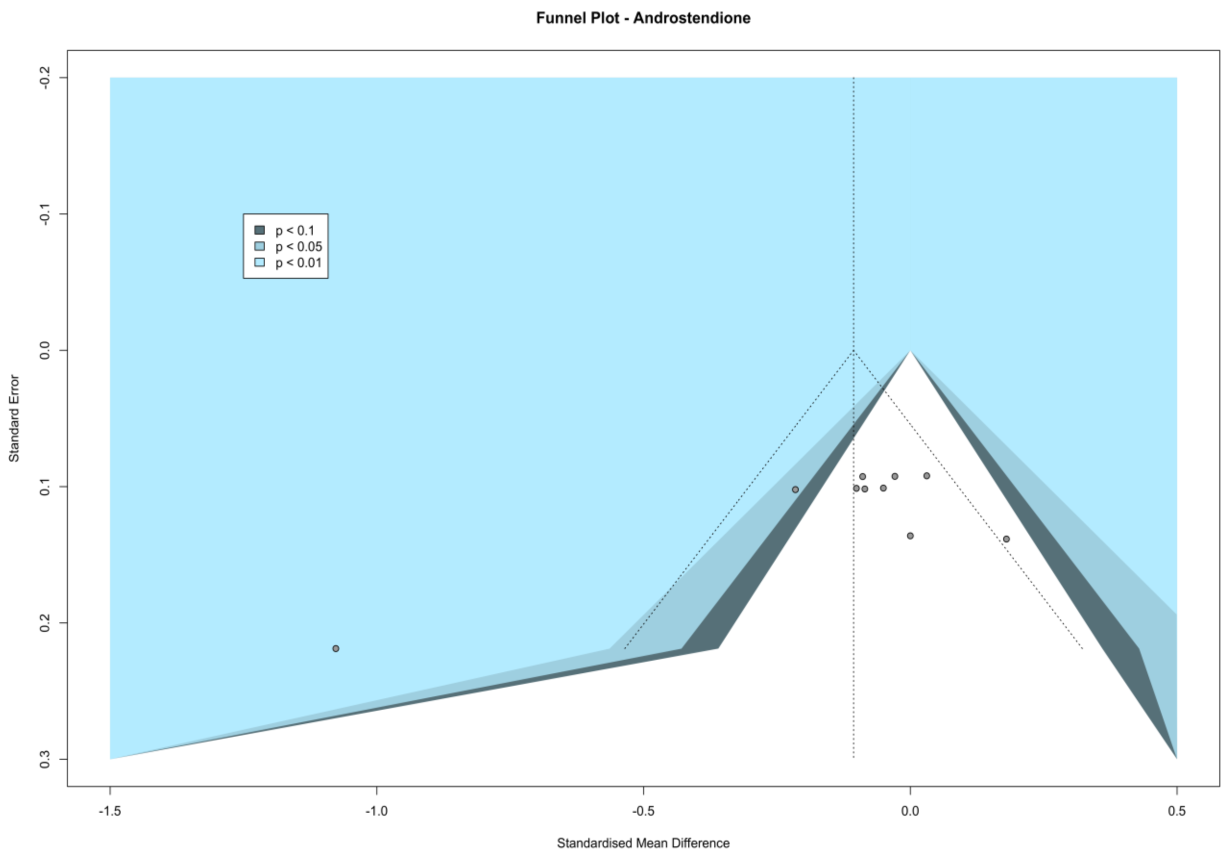

Fig. S22. Circulating androstenedione – Funnel plot of lifestyle intervention studies.

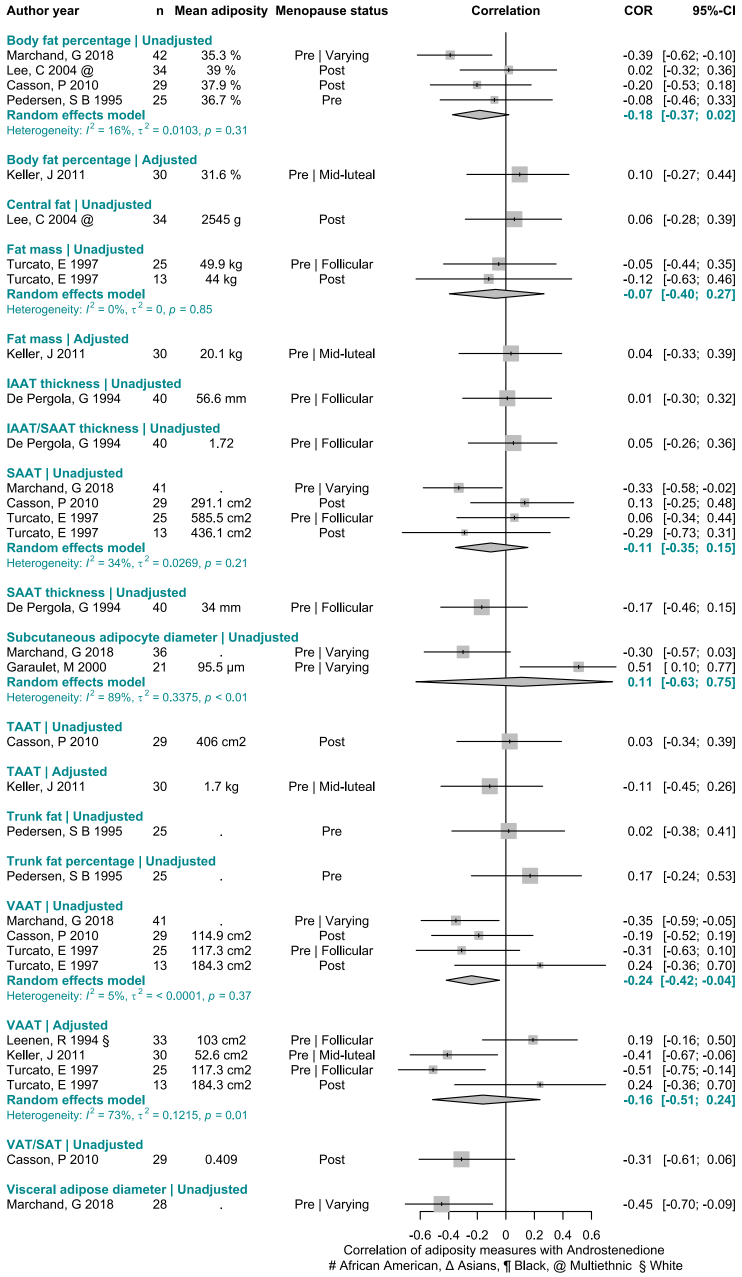

Fig. S23. Circulating androstenedione – Meta-analysis of correlations with direct adiposity measures.

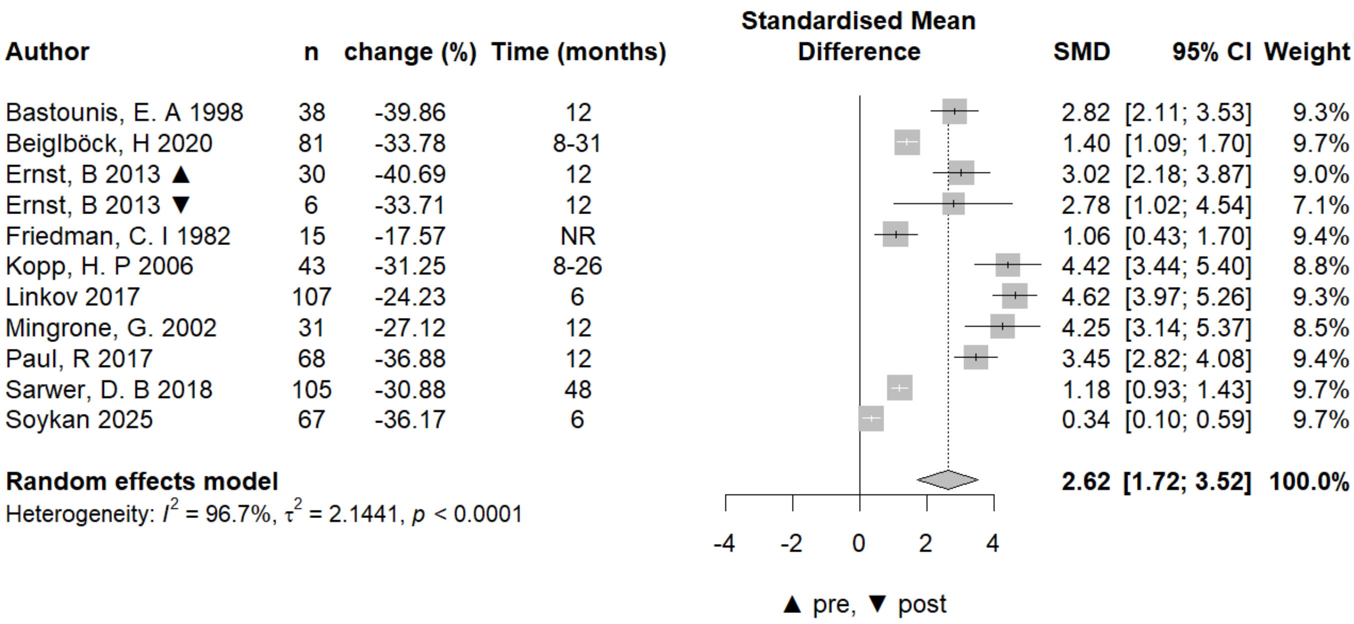

Fig. S24. Circulating SHBG – Meta-analysis of of-surgical intervention studies.

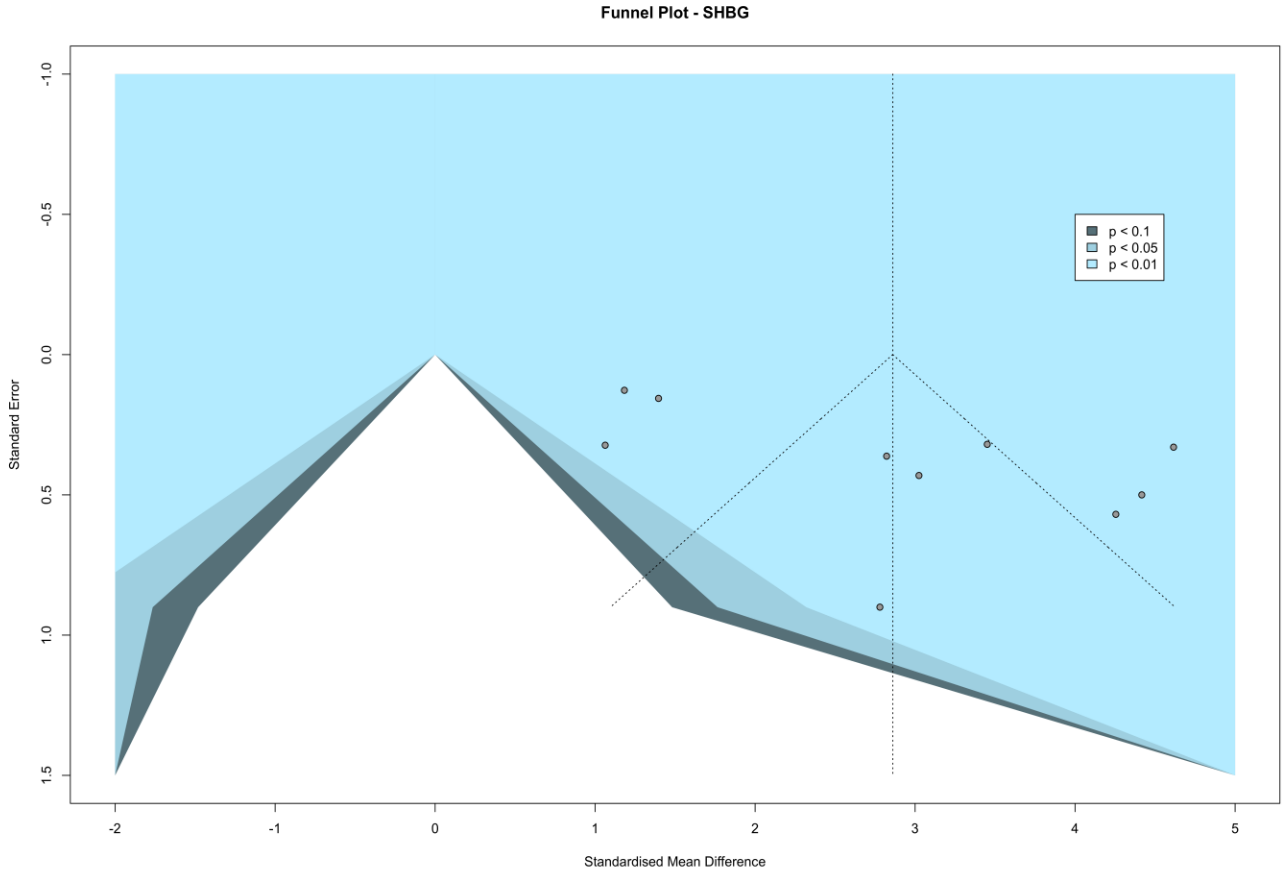

Fig. S25. Circulating SHBG - Funnel plot of surgical intervention studies.

| Author | n | change (%) | Time (months) | SMD | 95% CI | Weight |
|---|---|---|---|---|---|---|
| Leenen, R 1994 | 33 | -29.62 | 6 | 0.50 | [ 0.14; 0.86] | 3.2% |
| Elsayed, M 2022 + | 30 | -11.46 | 3 | 1.18 | [ 0.71; 1.64] | 3.1% |
| Aubuchon, M 2016 | 30 | -9.42 | 4-6 | 0.16 | [-0.20; 0.52] | 3.2% |
| Kovacikova, M. 2008 | 32 | -8.01 | 3 | 2.44 | [ 1.75; 3.14] | 2.9% |
| Van Gemert, W 2015 * | 98 | -6.90 | 4 | 0.45 | [ 0.24; 0.66] | 3.2% |
| Van Rossum, EF 2000 | 42 | -6.87 | 6 | 0.19 | [-0.12; 0.49] | 3.2% |
| Elsayed, M 2022 ° | 30 | -6.53 | 3 | 0.44 | [ 0.06; 0.81] | 3.2% |
| Van Gemert, W 2015 ° | 97 | -6.14 | 4 | 0.30 | [ 0.10; 0.50] | 3.2% |
| Kim, J 2012 | 15 | -3.28 | 4 | 0.29 | [-0.22; 0.81] | 3.1% |
| Bhargava, A. 2006 | 615 | -2.75 | 12 | 0.25 | [ 0.17; 0.33] | 3.3% |
| Kraemer, W. J 2003 | 9 | -16.59 | 9 | 0.00 | [-0.65; 0.66] | 3.0% |
| Campbell, K. L 2012 + | 117 | -13.29 | 12 | 0.71 | [ 0.51; 0.91] | 3.2% |
| Caballero MJ 1992 | 18 | -11.73 | 4 | -0.33 | [-0.81; 0.14] | 3.1% |
| Campbell, K. L 2012 ° | 118 | -10.43 | 12 | 0.50 | [ 0.31; 0.69] | 3.2% |
| Kraemer, W. J 2003 | 10 | -8.86 | 9 | 0.13 | [-0.49; 0.75] | 3.0% |
| Harvie, M. N 2011 / | 53 | -7.90 | 6 | 0.30 | [ 0.02; 0.57] | 3.2% |
| Harvie, M. N 2011 # | 54 | -6.17 | 6 | 0.15 | [-0.12; 0.42] | 3.2% |
| Campbell, K. L 2012 * | 117 | -3.81 | 12 | -0.02 | [-0.20; 0.16] | 3.2% |
| Smith, A. J 2011 | 166 | -2.71 | 4 | -0.06 | [-0.21; 0.09] | 3.3% |
| Svendsen, OL 1995 ● | 98 | -26.57 | 3 | 0.70 | [ 0.48; 0.92] | 3.2% |
| Svendsen, OL 1995 ■ | 98 | -25.77 | 3 | 0.91 | [ 0.67; 1.14] | 3.2% |
| Ricci 2001 | 14 | -18.70 | 6 | 0.78 | [ 0.18; 1.37] | 3.0% |
| Gonzalo□Encabo, P | 214 | -12.01 | 12 | 0.25 | [ 0.11; 0.38] | 3.3% |
| van Gemert, W 2017 | 242 | -10.29 | 4 | 0.31 | [ 0.18; 0.44] | 3.3% |
| Van Rossum, EF 2000 | 42 | -16.10 | 6 | 0.19 | [-0.12; 0.49] | 3.2% |
| O'Dea, J. P. K 1979 | 12 | -20.06 | 3-4 | 4.19 | [ 2.42; 5.96] | 1.9% |
| Duggan, C 2019 | 31 | -10.00 | 30 | 2.54 | [ 1.81; 3.26] | 2.9% |
| Rock, C. L. 2016 -- | 119 | -9.28 | 12 | 0.52 | [ 0.33; 0.71] | 3.2% |
| Rock, C. L. 2016 - | 119 | -8.65 | 12 | 0.76 | [ 0.56; 0.97] | 3.2% |
| Stolzenberg-Solomon, R. Z 2012 | 278 | -8.64 | 6 | 5.00 | [ 4.57; 5.43] | 3.1% |
| Mingrone, G. 2002 | 21 | -5.84 | 12 | 0.54 | [ 0.08; 1.00] | 3.1% |
| Rock, C. L. 2016 --- | 119 | -4.92 | 12 | 0.24 | [ 0.06; 0.43] | 3.2% |
| **Random effects model** | | | | **0.71** | **[ 0.34; 1.07]** | **100.0%** |

Heterogeneity: $I^2 = 95.5\%$, $\tau^2 = 1.0628$, $p < 0.0001$

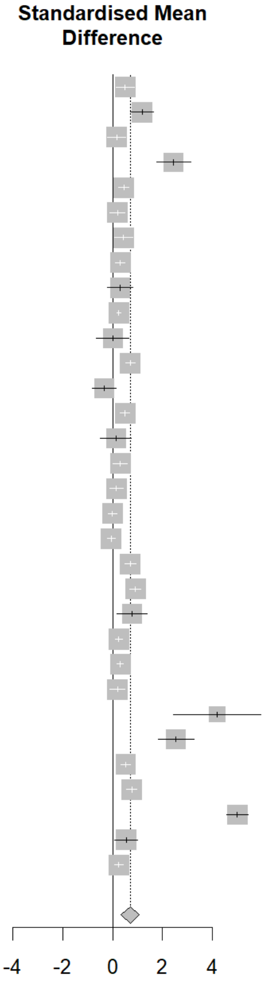

/ Intermitted diet, # energy restriction ° diet, * exercise, + diet and exercise,
● gynoid, ■ android,- low fat, -- walnut, --- low carb

Fig. S26. Circulating SHBG – Meta-analysis of lifestyle intervention studies.

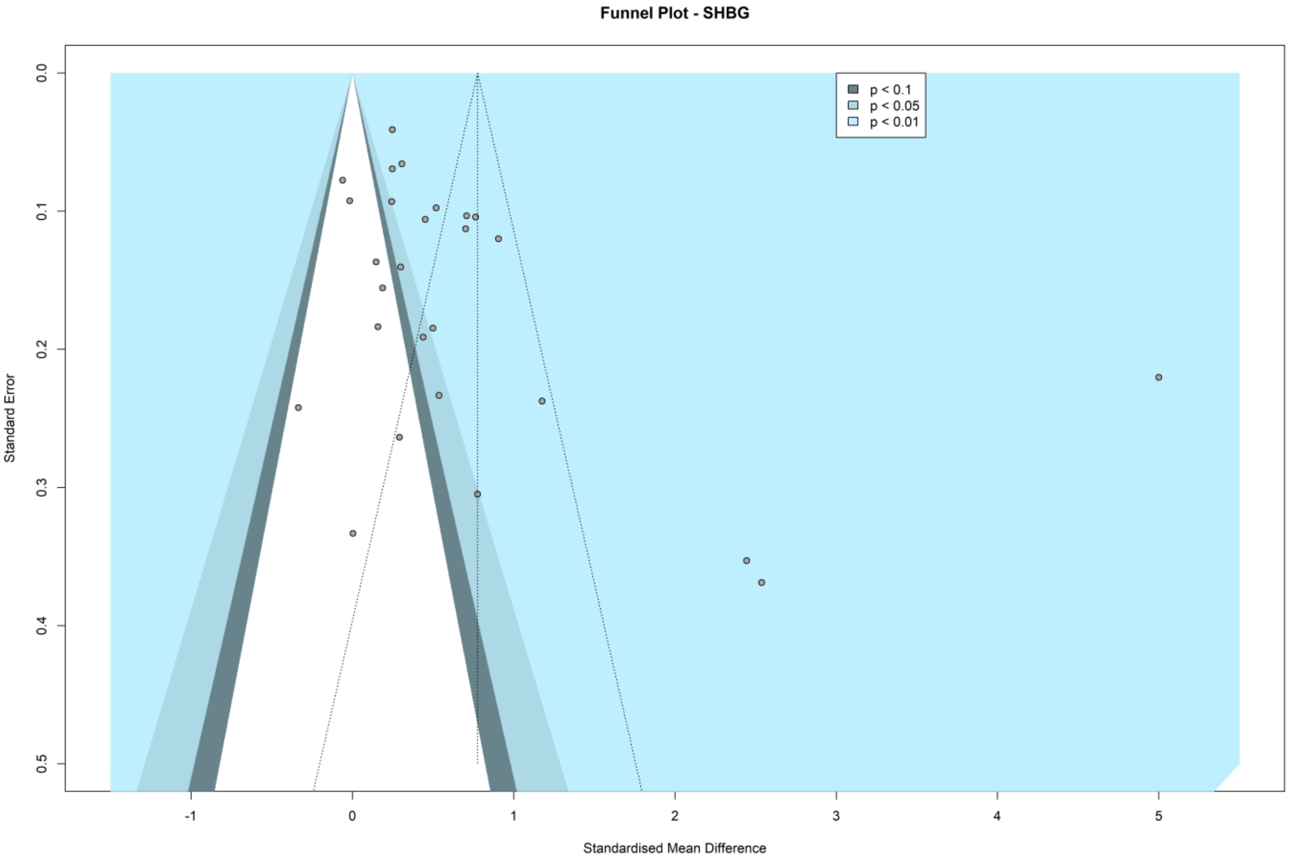

Fig. S27. Circulating SHBG - Funnel plot of lifestyle intervention studies.

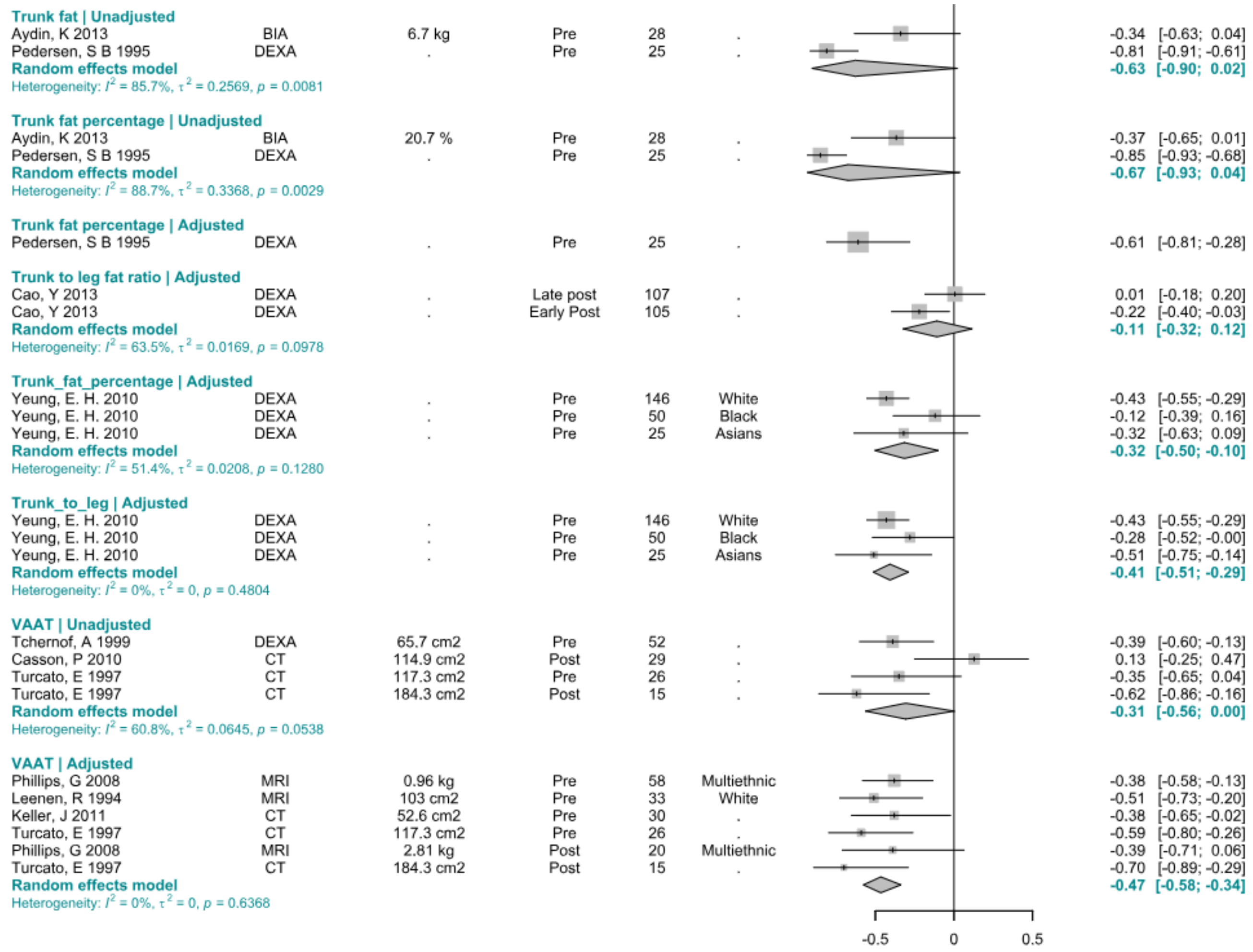

Fig. S28. Circulating SHBG – Meta-analysis of correlations with adiposity measures.

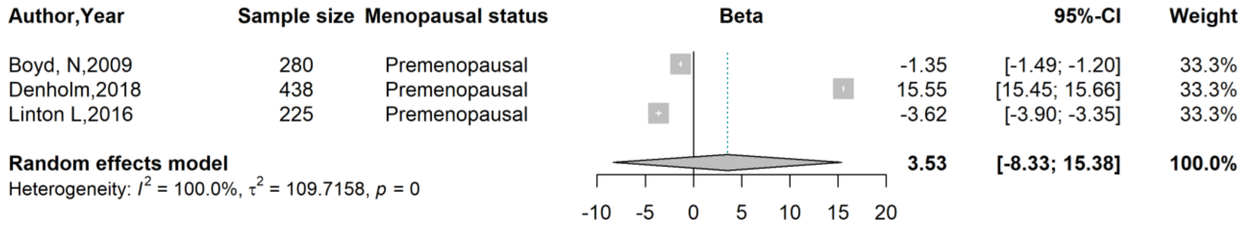

Fig. S29. Circulating SHBG - Meta-analysis of standardized beta coefficient from adjusted linear regression for the association between SHBG and breast fat volume.

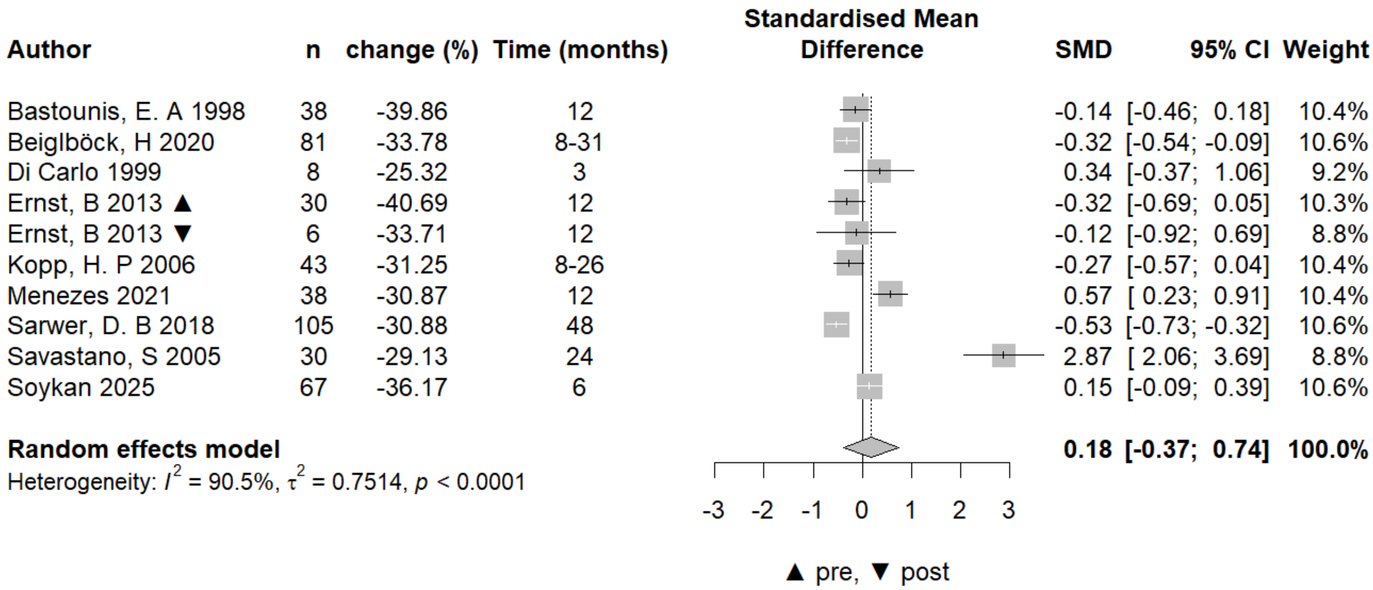

Fig. S30. Circulating DHEAS – Meta-analysis of surgical intervention studies.

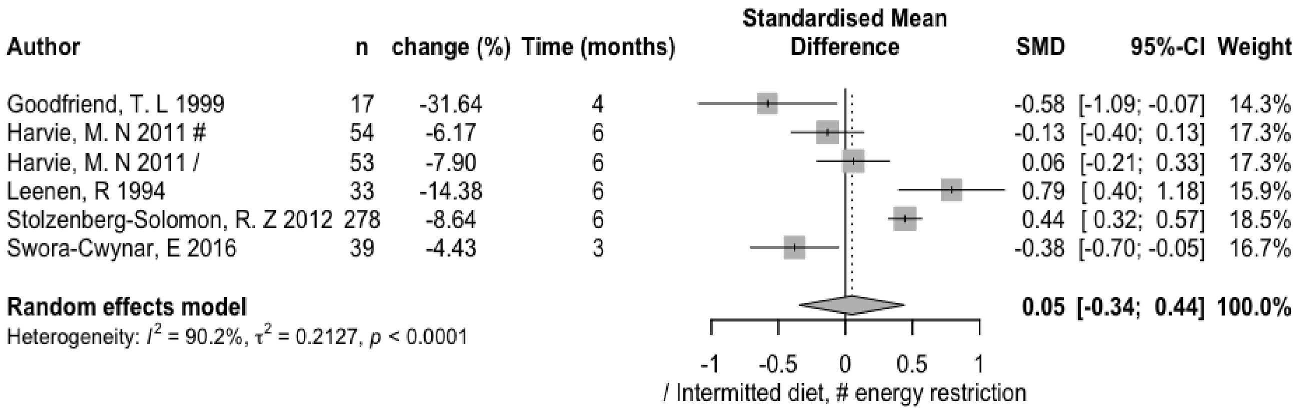

Fig. S31. Circulating DHEAS – Meta-analysis of lifestyle intervention studies.

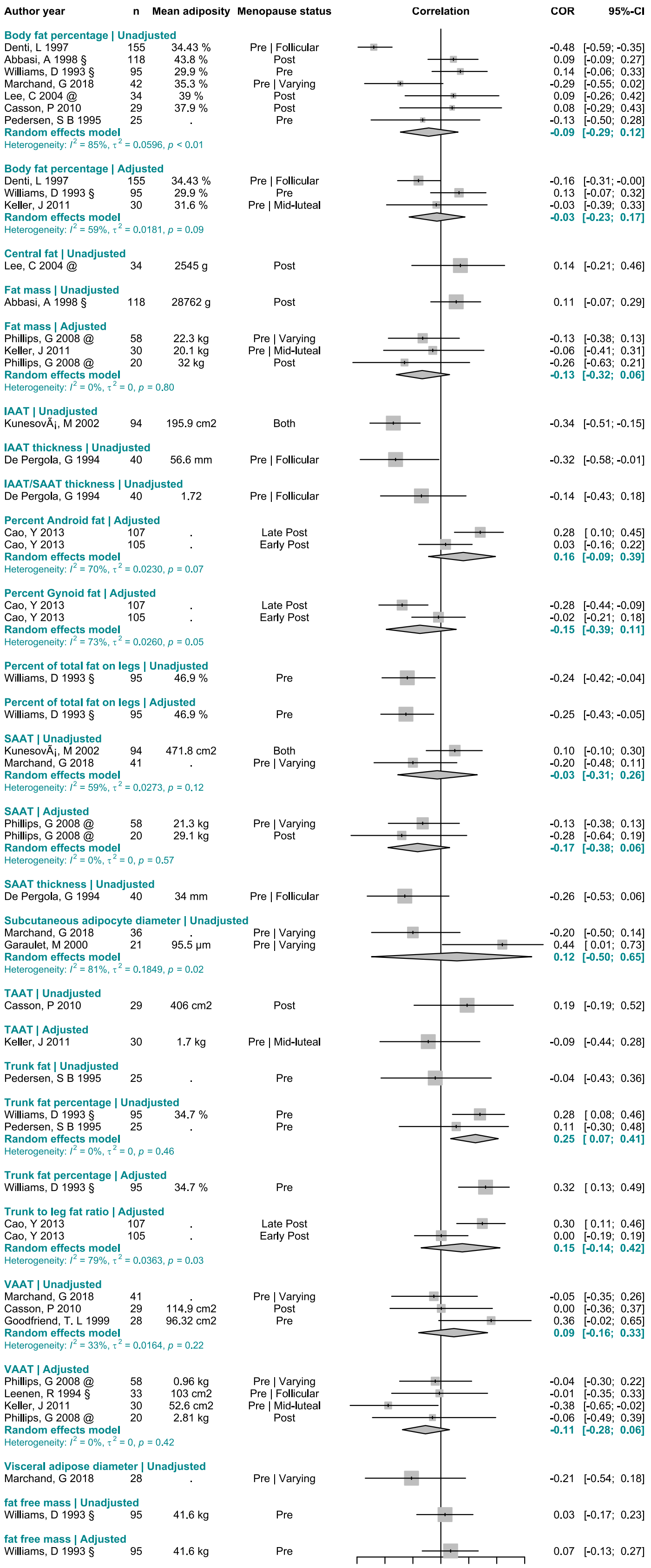

Fig. S32. Circulating DHEAS – Meta-analysis of correlations with direct adiposity measures.
